# Time-restricted eating and exercise training before and during pregnancy for people with increased risk of gestational diabetes: the BEFORE THE BEGINNING randomised controlled trial

**DOI:** 10.1101/2024.11.08.24316895

**Authors:** MAJ Sujan, HMS Skarstad, G Rosvold, SL Fougner, T Follestad, KÅ Salvesen, T Moholdt

## Abstract

**Objective:** To determine the effect of a pre-pregnancy lifestyle intervention on glucose tolerance in people at higher risk of gestational diabetes mellitus (GDM).

**Design:** Randomised controlled trial.

**Setting:** University hospital in Trondheim, Norway.

**Participants:** 167 people with at least one risk factor for GDM who contemplated pregnancy.

**Intervention:** The participants were randomly allocated (1:1) to a lifestyle intervention or a standard care control group. The intervention consisted of exercise training and time-restricted eating, started pre-pregnancy and continued throughout pregnancy. Exercise volume was set using a physical activity metric that translates heart rate into a score (Personal Activity Intelligence, PAI), with the goal of ≥ 100 weekly PAI-points. Time-restricted eating involved consuming all energy within ≤ 10 hours/day, ≥ 5 days per week.

**Main outcome measures:** 2-hour plasma glucose level in an oral glucose tolerance test (OGTT) in gestational week 28. The primary analysis used an intention-to-treat principle.

**Results:** From 02.10.2020 to 12.05.2023, we included 167 participants: 84 in intervention and 83 in control, out of whom 111 became pregnant (56 in intervention and 55 in control). One participant in the intervention group was excluded from the analysis because of pre-pregnancy diabetes. Pregnancy data from one participant in the control group were excluded from the analysis because of twin pregnancy. Time to pregnancy was 112 days (SD 105) in the intervention (INT) group and 83 days (SD 69) in the control (CON) group (p = .087). The intervention had no significant effect on 2-hour plasma glucose level in an OGTT in gestational week 28 (mean difference, 0.48 mmol/L, 95% confidence interval [CI], -0.05 to 1.01, p = .077). There was no evidence of between-group differences in other measures of glycaemic control before or during pregnancy. The intervention did not significantly influence GDM prevalence rates in gestational week 12 (INT 5.5%, CON 5.6%, p = 1.000) or gestational week 28 (INT 14.5%, CON 11.1%, p = .592). In gestational week 28, the intervention group had gained less weight (2.0 kg, 95% CI, -3.3 to -0.8, p = .002) and fat mass (-1.5 kg, 95% CI, -2.5 to -0.4, p = .008) than the control group. Participants could adhere to the ≤ 10-hour eating window and maintain ≥ 100 PAI per rolling week pre-pregnancy, but adherence to both intervention components decreased during pregnancy.

**Conclusions:** A combination of time-restricted eating and exercise training started before and continued throughout pregnancy had no significant effect on glycaemic control in late pregnancy, but our findings suggest that the intervention lowered gestational weight and fat mass gain in people with increased risk of GDM.

**Trial registration:** ClinicalTrials.gov NCT04585581

## Introduction

Gestational diabetes mellitus (GDM) is defined as hyperglycaemia first diagnosed during pregnancy and affects approximately one out of seven live births globally.(1) Risk factors for GDM include having a high body mass index (BMI) and excessive gestational weight gain, older age, GDM in a previous pregnancy, a family history of diabetes, and non-European ethnicity.(1) Chronic insulin resistance as a result of a complex interplay of these genetic, environmental, and behavioural risk factors aggravates physiological insulin resistance in the second half of pregnancy leading to pancreatic β-cell dysfunction, elevated glucose levels, and eventually GDM.(2) Although the hyperglycaemia usually resolves after delivery, GDM is associated with an increased risk for several adverse consequences later in life, including recurrence of GDM in later pregnancies, type 2 diabetes, metabolic syndrome, and cardiovascular diseases.(3–6) Children of mothers with GDM are at increased risk of cardiac dysfunction at birth, childhood obesity, and future diabetes mellitus, thus continuing the intergenerational cycle of obesity and diabetes.(7–11)

Conventional lifestyle recommendations in pregnancy include moderate-intensity exercise for at least 150 minutes per week and a healthy diet.(12) However, many pregnant individuals fail to meet these recommendations and it may be difficult to change lifestyle because of biological changes in pregnancy and due to concerns about the effects on the growing fetus.(13–15) Most clinical trials of lifestyle interventions in pregnancy have initiated the intervention around 16-20 weeks of gestation, leaving a missed window of opportunity to implement lifestyle changes and improve glycaemic control.(16) Since pre-pregnancy patterns of physical activity are associated with exercise during pregnancy(17) and pre-pregnancy healthy dietary habits are associated with a lower risk of GDM,(18) the pre-pregnancy period can be a “teachable moment” to make favourable changes that can improve maternal and fetal outcomes. Over the last years, several systematic reviews and meta-analyses have concluded that pre-pregnancy lifestyle interventions are necessary to improve adherence and reduce the risk of GDM and related short- and longer-term adverse outcomes.(19–21) However, the evidence on the specific components of pre-pregnancy interventions and their effectiveness remains scarce. Alternative diet-exercise intervention strategies, such as time-restricted eating (TRE) and high-intensity interval training (HIIT) improve glycaemic control, and cardiometabolic outcomes in non-pregnant people with cardiometabolic disorders.(22–26) The data on the effects of TRE in pregnancy are scarce. We recently showed that it was feasible to consume all energy within maximum 10 h/day for 5 weeks in the second and third trimesters of pregnancy, albeit without effect on the measured cardiometabolic outcomes.(27) However, observational data indicate that longer night-fasting duration is associated with improved fasting glucose in pregnant individuals.(28) Recent publications show that HIIT is safe and enjoyable, as well as relatively easy to adhere to, during pregnancy.(29–31) In the BEFORE THE BEGINNING trial, we hypothesised that combined TRE and exercise training commenced pre-pregnancy and continued throughout pregnancy, would improve maternal glucose tolerance in gestational week 28 in people at increased risk of GDM.

## Methods

### Study design

BEFORE THE BEGINNING was a single-centre randomised controlled trial with two parallel groups: an intervention group and a control group (Figure 1). The trial was undertaken at the Norwegian University of Science and Technology (NTNU) in Trondheim, Norway, in collaboration with the St. Olav’s Hospital, Trondheim, Norway. The trial was registered in ClinicalTrials.gov (NCT04585581) on 25.09.2020. A detailed study protocol has been published previously.(32)

**Figure 1.**
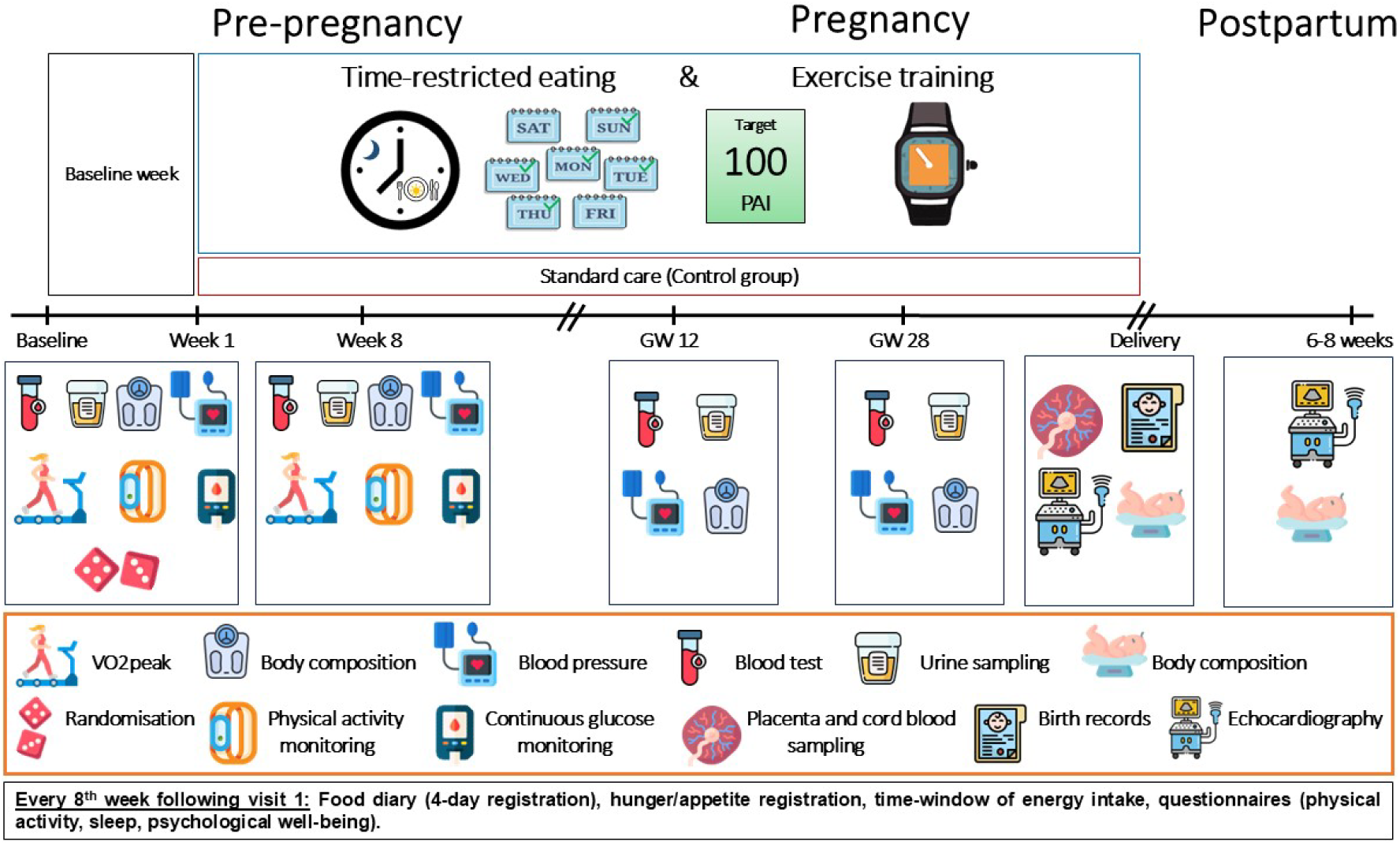
Study design. After baseline assessments, the participants were randomly allocated (1:1) to a lifestyle intervention or a standard care control group. The intervention consisted of exercise training and time-restricted eating, started pre-pregnancy, and continued throughout pregnancy. The amount of exercise was set using a heart rate-based physical activity metric (Personal Activity Intelligence, PAI), with the goal of ≥ 100 weekly PAI-points. Time-restricted eating involved consuming all energy within ≤ 10 hours/day, ≥ 5 days per week. Further assessments were performed 8 weeks after baseline in the pre-pregnancy period, and in gestational weeks 12 and 28. Pregnancy and birth outcomes were collected from hospital records after delivery. Neonatal outcomes were assessed within 72 hours after birth and 6-8 weeks after birth. GW = gestational week

### Recruitment and participants

We advertised the trial on social media, hospital and university websites, local stores, and public places. Moreover, we sent electronic invitations to the trial to all females aged 20-35 years in Trondheim and the surrounding area using information obtained from the national population register. We included females who were 18-39 years old and contemplating pregnancy within the next 6 months, who understood oral and written Norwegian or English and who had at least one of the following risk factors: BMI ≥ 25 kg/m^2^, GDM in a previous pregnancy, close relative with diabetes (parents, siblings, or children with diabetes), fasting plasma glucose > 5.3 mmol/L, previous newborn > 4.5 kg, or non-European ethnicity (one or both parents originating from an area outside Europe). Exclusion criteria were ongoing pregnancy, trying to conceive ≥ 6 cycles at study entry, known diabetes (type 1 or 2), shift work that included night shifts > 2 days/week, previous hyperemesis, known cardiovascular diseases, high-intensity exercise > 2 times/week in the last 3 months, habitual eating window ≤ 12 hours/day, bariatric surgery, or any other reason which according to the researchers made the potential participant ineligible.

### Randomisation and blinding

Baseline assessments were performed before we randomly allocated the participants (1:1) to the intervention or a standard care control group, stratified by GDM in a previous pregnancy (yes/no). The study personnel used WebCRF3, a computer random number generator developed and administered at The Clinical Research Unit (Klinforsk), NTNU/St. Olav’s Hospital, Trondheim, Norway to randomly allocate participants using various block sizes. The randomisation sequence was concealed until interventions were assigned. Neither participants nor study personnel were blinded.

### Intervention and adherence

The intervention consisted of TRE and exercise training and spanned from inclusion pre-pregnancy and throughout pregnancy. We counselled the participants in the intervention group to restrict their daily time-window of energy intake to ≤ 10 hours, ending no later than 19:00 hours, for minimum 5 days per week throughout the study period. On the “off” days, the participants could choose their time-window for energy intake. We gave no advice about dietary composition or the amount of energy the participants should consume, and they could consume non-energy drinks outside the time-window.

The exercise programme was based on Personal Activity Intelligence (PAI), which is a physical activity metric that translates heart rate during physical activity into a score.(33) We instructed the participants to obtain ≥ 100 weekly PAI-points throughout the study period since this amount of physical activity is associated with higher cardiorespiratory fitness and a decreased cardiovascular disease risk.(33,34) High-intensity exercise and thus higher heart rates give substantially more PAI-points than low-to-moderate-intensity exercise. The participants in the intervention group wore smartwatches (Amazfit GTS and/or Polar Ignite 2) throughout the study period. These watches shared PAI data with the research team.

The exercise training consisted of endurance exercise with the aim of high intensity and was mainly unsupervised. We provided participants in the intervention group with a brochure suggesting various HIIT sessions to complete at home (Supplementary file 1). The participants were free to choose whether to exercise indoors or outdoors, and if they wanted to use any cardio machines (e.g., treadmill, stationary bike). Once pregnant, we advised the participants to choose between short work-bouts (30 seconds) at high intensity with low-to-moderate intensity periods in-between, or longer work periods with an intensity up to 85% of heart rate maximum. The participants were invited for supervised exercise sessions twice after baseline: after 2 and 4 weeks. Additionally, participants could ask for extra support and more supervised sessions at any time during the intervention period. The supervised sessions were undertaken on a treadmill or a stationary bike in our training facility. Participants in the control group received standard care and were asked to continue with their habitual physical activity and dietary intake.

We asked all participants to report their daily time-window of energy intake for 4 days (three weekdays and one weekend day) every 8^th^ week in a printed study handbook. We categorised participants in the intervention group as adherent to TRE if they reported a ≤ 10-hour time window for energy intake on ≥ 2 of these 4 days and as adherent to exercise training if they earned and maintained ≥ 75 PAI per rolling week. We sent reminders to all participants via text messages to complete dietary reporting and contacted the participants in the intervention group to offer additional supervised exercise sessions if they were not reaching the PAI target.

### Experimental procedures and outcome measures

Assessments of the participants were undertaken twice during pre-pregnancy and twice during pregnancy. The first week after allocation to groups consisted of baseline recordings, followed by visits in the laboratory after 8 weeks, and in gestational weeks 12 and 28. Participants who did not become pregnant within 6 months of inclusion were excluded from the study, but their pre-pregnancy data were included in the intention-to-treat analysis (changed from 12 months, see modifications to the protocol after trial commencement). In cases of spontaneous abortions, we adjusted the period to allow the participants to continue in the study by a 4-weeks extension added to the number of weeks the participants were pregnant before abortion.

#### Primary outcome measure

The primary outcome measure was 2-hour plasma glucose concentration in a 75 g oral glucose tolerance test (OGTT) in gestational week 28. After an overnight fast (≥ 10 hours) and no exercise for ≥ 24 hours, the participants consumed a premade drink of 75 g glucose diluted in 250 mL water (Glucosepro, Finnamedical, Finland) within 5 minutes. Using an indwelling catheter, we collected venous blood before the OGTT, with subsequent collections at 30, 60, 90, and 120 minutes after ingestion of glucose.

#### Secondary outcome measures

The trial has several secondary outcomes, as specified in the study protocol.(32) Here we report the main secondary maternal cardiometabolic outcomes and will report the remaining secondary outcomes in separate publications. Secondary outcome measures were assessed twice during pre-pregnancy (at baseline and after 8 weeks) and twice during pregnancy (in gestational weeks 12 and 28), if not otherwise specified. Fasting plasma glucose, blood lipids, and glycated haemoglobin (HbA1c) were analysed immediately after blood sampling at St. Olavs Hospital laboratory, following local standardised procedures. GDM was recorded in gestational weeks 12 and 28, and diagnosed according to the WHO 2013 criteria: fasting plasma glucose 5.1-6.9 mmol/L and/or 2-hour plasma glucose 8.5-11.0 mmol/L after a 75-g glucose load.(35) We measured fasting, 30-minute, and 120-minute insulin concentrations in thawed serum samples with enzyme-linked immunosorbent assay (ELISA, IBL-International, Hamburg, Germany) according to the manufacturer’s protocol using a DS2 ELISA processing system (Dynex technologies, Virginia, USA) at the research laboratories at the Department of Circulation and Medical Imaging, NTNU. The area under the curve (AUC) and incremental AUC (iAUC) from glucose concentrations were calculated from venous blood sampling obtained every 30 minutes during the OGTT.(36) We calculated homeostasis model assessment of insulin resistance (HOMA2-IR) and pancreatic beta cell function (HOMA2-β) using the online HOMA2 calculator: https://www.dtu.ox.ac.uk/homacalculator/index.php (37) We also estimated the Insulin Sensitivity Index (ISI_0,120_)(38), insulinogenic index during the first 30 minutes of the OGTT(39), and beta cell function (AUC_ins_/AUC_glu_)(40).

Weight and body composition were estimated in the morning after overnight fasting using bioelectrical impedance analysis (Inbody 720, Biospace CO, Korea), with participants wearing light clothes and standing barefoot. Additionally, waist circumference was measured using a measuring tape at the level of the navel, in a standing position. We used an automatic blood pressure device (Welch Allyn, Germany) to measure blood pressure and resting heart rate on the participants’ left arm after they had rested in a seated position for 15 minutes. The average of three measurements taken 1 minute apart is reported. We asked the participants to register their diet in an online food diary (Fatsecret app) intake for 4 days (three weekdays and one weekend day) every 8^th^ week. They also completed the International Physical Activity Questionnaire(41) every 8^th^ week throughout the study period.

#### Modifications to the protocol after trial commencement

From November 2022, we started identifying eligible participants using population data from the Norwegian Tax Administration and sending out electronic participant invitations to reach a broader target population. Concurrently, we added ‘bariatric surgery’ and ‘any other reason which according to the researchers makes the potential participant ineligible’ to undergo either or both interventions (e.g., traumatic foot injury, anorexia/bulimia) as exclusion criteria. From December 2022, we removed ‘planned assisted fertilisation with female factor reason’ from the exclusion criteria. The changes in the inclusion criteria were made to account for challenges that arose during the screening process. Also in December 2022, we changed the maximum time before pregnancy from 12 months to 6 months to allow for the trial to be terminated in time for us to analyse the data within the project period. In March 2023, the required number of total participants was reduced from 260 to 200 based on the revised sample size calculation described below. In June 2023, we changed from Amazfit GTS (Huami, China) to Polar Ignite 2 (Polar, Finland) smartwatch and from Zepp and Memento to Polar Flow and Mia app to improve recording of PAI-points.

#### Sample size

The primary outcome measure was 2-hour plasma glucose level in a 75 g OGTT in gestational week 28. We considered a difference between the intervention group and the control group of 1.0 mmol/L as the minimally clinically relevant difference, based on findings from the HAPO study,(7) and have used the observed standard deviation (SD) in 2-hour plasma glucose concentrations during the OGTT from HAPO in the calculations. Sample size calculation for a two-sided t-test to detect a 1.0 mmol/L difference between the groups, using an SD of 1.3, a power of .90, and a significance level of .05, yields 37 participants in each group in gestational week 28. To allow for an expected exclusion from the study due to not conceiving within the study period (∼50%)(42) yielding 74 per group, further drop-out during the study period (10-20%), yielding 93 per group, and to increase statistical power for secondary analyses, we initially wanted to include 260 participants in the trial. However, we concluded participant recruitment after reaching 167 participants, because we at that point had more than 45 participants per group in gestational week 12, which allowed for an anticipated 20% dropout during pregnancy.

#### Statistical analysis

We analysed data from 166 participants in the intention-to-treat analyses (excluding one participant in the intervention group because of pre-pregnancy diabetes). For the analysis of pregnancy-specific data, we used data obtained from 109 pregnant participants (data obtained during pregnancy from one participant in the control group were excluded from the analysis because of twin pregnancy). We used linear mixed models to estimate differences in primary and secondary continuous outcomes between groups, with time and the interaction between time and group as fixed effects variables, and subject (participant ID) as random effect.(43) Since no systematic baseline differences between the groups are expected in randomised controlled trials, the main effect of group was not included, such that the means at baseline were constrained to be equal in these models. We report estimated effects in the intervention group compared with the control group, with corresponding 95% confidence intervals and *p*-values. For outcomes obtained only during the pregnancy period (variables using data from the OGTT at gestational week 12 and 28), we included the main effect of group in the linear mixed model.

We checked the normality of residuals by visually inspecting QQ-plots. For variables that were not normally distributed, bias-corrected and accelerated bootstrap confidence intervals (CIs) based on 3000 bootstrap samples were calculated. Additionally, we used Fisher’s exact test to compare GDM prevalence in gestational week 12 and chi-square test in gestational week 28, and Student’s t-test to compare time to pregnancy between groups. For our primary outcome measure, we considered a *p* -value < .05 to indicate statistically significant results. For the secondary outcome measures, we used a significance level of .01 to give some protection against fall positives due to multiple comparisons. We also performed pre-specified per-protocol analyses, in which we included only the participants in the intervention group who obtained ≥ 75 weekly PAI-points and reported a ≤ 10-hour time window for energy intake on ≥ 2 out of 4 days in the handbook pre-pregnancy. The statistical analyses were performed using IBM SPSS Statistics 29.0 and STATA MP version 18.

### Patient and public involvement

In the planning phase, we invited users to discuss the study 1 month before applying to the regional ethical committee for approval. We arranged a 1-hour interactive digital workshop with participant representatives (reproductive-aged females with overweight/obesity) to discuss relevant topics or issues related to participation and long-term adherence. Among the topics we discussed were potential barriers to participation in diet-exercise interventions, how to engage participants and keep them engaged, how to increase motivation, use of digital methods to collect exercise- and diet data, and recruitment strategies. Four months after including the first participant, we organised another interactive workshop in which we discussed how to increase participant recruitment and adherence. Based on input from the participant representatives, we established a Facebook group for planning exercise sessions together with other participants. Additionally, we offered supervised exercise training for interested participants at our training facilities via the Facebook group. Participant representatives were not involved in the recruitment process. In February 2023, we had another interactive meeting with participants who completed the study to discuss their views on exercise training during pregnancy and follow-up before and during pregnancy.

## Results

### Participants

We randomised 167 participants (Intervention, *n* = 84 and Control, *n* = 83) between October 2^nd^ 2020 and May 12^th^ 2023 (Figure 2). We ended inclusion of new participants when 45 participants in each group reached gestational week 12, as per our sample size calculations. We excluded one participant in the intervention group because of pre-pregnancy diabetes, leaving data from 166 participants in the intention-to-treat analyses. Within the specified time, 111 participants became pregnant (Intervention, *n* = 56, Control, *n* = 55). Pregnancy data from one participant in the control group were excluded from the analysis because of twin pregnancy (Figure 2). Time to pregnancy did not differ significantly between groups, with mean of 112 days (SD 105) in the intervention group and 83 days (SD 69) in the control group (*p* = .087). Table 1 shows the baseline characteristics of the participants.

**Figure 2.**
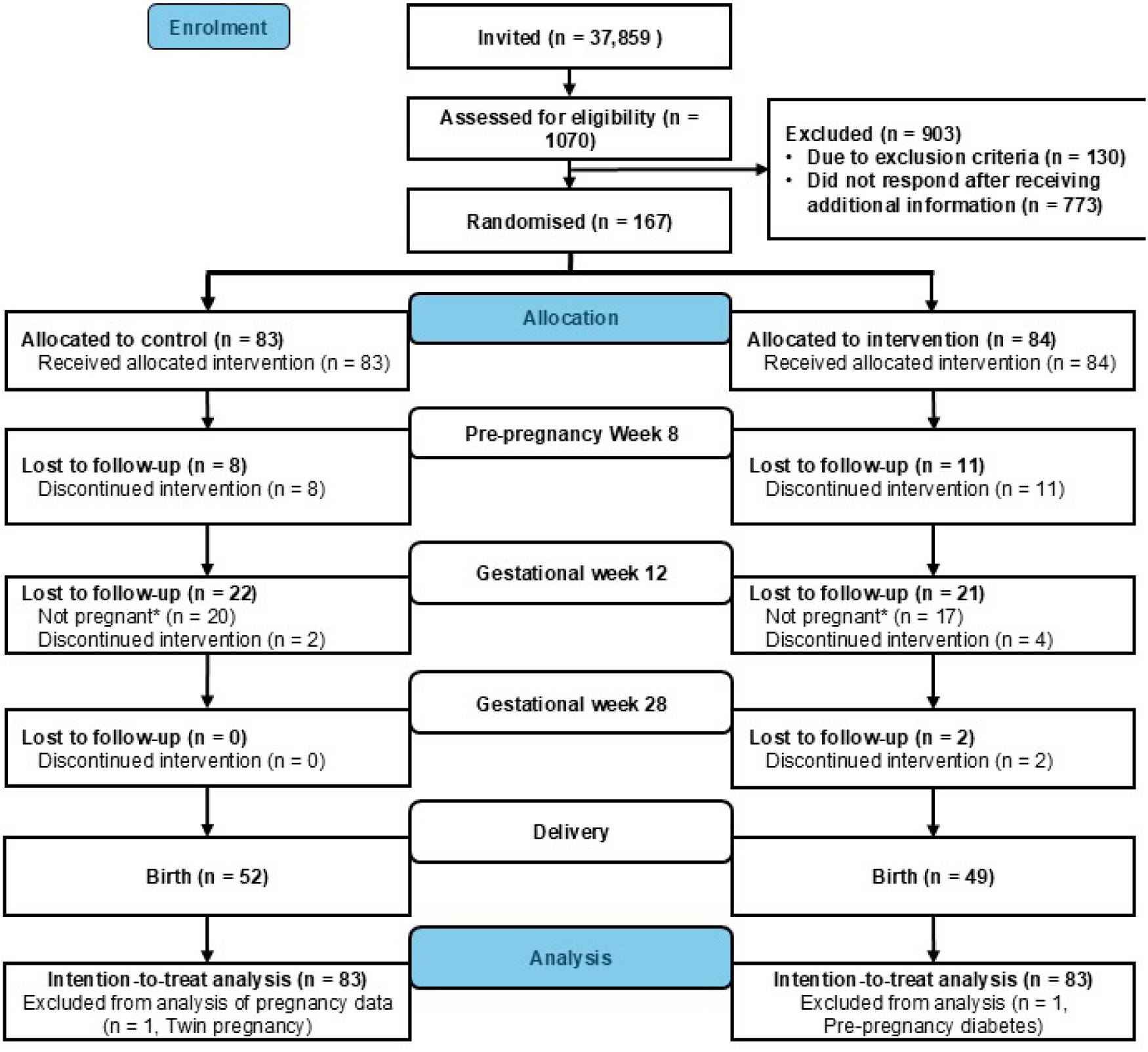
Flowchart of participants. * Participants who did not conceive within the specified time were excluded from further participation in the trial, but their pre-pregnancy data were included.

**Table 1.** Baseline characteristics of participants, according to group allocation.

|  | Control (n = 83) | Intervention (n = 83) |
| --- | --- | --- |
| Age, years | 30.3 (3.2) | 30.2 (3.1) |
| Weight, kg | 81.5 (13.2) | 81.1 (16) |
| Body mass index, kg/m<sup>2</sup> | 29.1 (4.5) | 29.2 (4.9) |
| Waist circumference, cm | 94.5 (11.8) | 93.0 (12.3) |
| Muscle mass, kg | 27.7 (3.6) | 27.9 (4.3) |
| Fat mass, kg | 31.7 (9.9) | 30.8 (11) |
| Fat percentage, % | 37.9 (7.2) | 37.0 (7.6) |
| Visceral fat area, cm<sup>2</sup> | 153 (52) | 146 (56) |
| Systolic blood pressure, mmHg | 121 (10) | 118 (8) |
| Diastolic blood pressure, mmHg | 79 (7) | 79 (6) |
| Resting heart rate, bpm | 72 (11) | 71 (11) |
| HbA1c, mmol/mol | 34.3 (3.1) | 34.1 (2.8) |
| Fasting glucose, mmol/L | 5.0 (0.4) | 4.9 (0.4) |
| Total cholesterol, mmol/L | 4.5 (0.7) | 4.6 (0.8) |
| HDL cholesterol, mmol/L | 1.4 (0.3) | 1.4 (0.3) |
| LDL cholesterol, mmol/L | 3.0 (0.9) | 3.1 (0.8) |
| Triglycerides, mmol/L | 1.0 (0.5) | 1.0 (0.5) |
| Fasting insulin, µIU/mL | 19.1 (11.7) | 17.1 (10.4) |
| HOMA2-IR | 2.4 (1.4) | 2.2 (1.2) |
| HOMA2-B | 168.2 (59.2) | 164.9 (69.2) |
| **Parity, total number of births, n (%)** |  |  |
| 0 | 45 (55) | 47 (58) |
| 1 | 30 (37) | 30 (37) |
| 2 | 7 (9) | 4 (5) |
| **Education level, n (%)** |  |  |
| Compulsory schooling | 0 (0) | 1 (1) |
| Completed upper secondary school | 8 (10) | 10 (12) |
| Completed university education, less than 4 years | 28 (34) | 23 (28) |
| Completed university education, 4 years or more | 46 (56) | 47 (58) |
| **Ethnic origin, n (%)** |  |  |
| Europe | 74 (89) | 72 (87) |
| Africa and Middle East | 3 (4) | 1 (1) |
| Asia | 4 (5) | 6 (7) |
| North America | 1 (1) | 1 (1) |
| Latin America | 1 (1) | 3 (4) |
| **Reason for inclusion\*, n (%)** |  |  |
| Body mass index ≥ 25 kg/m<sup>2</sup> | 74 (89) | 68 (82) |
| GDM in a previous pregnancy | 1 (1) | 2 (2) |
| Family history of diabetes, *n* | 20 (24) | 24 (29) |
| Previous newborn > 4.5 kg, *n* | 1 (1) | 1 (1) |

Data are presented as means of observed values with standard deviation (SD) or number (n) and percentage (%) of participants. Missing: number of participants with missing data in each group is 0 to 3 for all variables except for waist circumference, where 5 are missing in the intervention group and 8 in the control group.
HbA1c = Glycated haemoglobin, HDL = High-density lipoprotein cholesterol, LDL = Low-density lipoprotein cholesterol, HOMA2-IR = Homeostatic model assessment of insulin resistance, HOMA2-B = Homeostatic model assessment of beta cell function, GDM = Gestational diabetes mellitus.

#### Primary outcome and secondary glycaemic outcomes in pregnancy

There was no statistically significant difference between the intervention and control groups in 2-hour plasma glucose concentrations during a 75 g OGTT (mean difference 0.48 mmol/L, 95% CI -0.05 to 1.01, *p* = .077) in gestational week 28 (Table 2, Figure 3). The intervention did not significantly improve secondary glycaemic outcomes (fasting glucose, fasting insulin, HbA1c, HOMA2-IR, and HOMA2-B) compared with control. No statistically significant between-group differences were found in glycaemic outcomes obtained only during pregnancy (AUC, iAUC, insulinogenic index during the first 30 minutes of OGTT and ISI_0,120_) (Supplementary file 2). In gestational week 12, three participants in each group fulfilled the criteria for GDM diagnosis (INT 5.5%, CON 5.6%, *p* = 1.000). The corresponding numbers in gestational week 28 were eight participants (14.5%) in the intervention group and six participants (11.1%) in the control (*p* = .776) (Supplementary file 2).

**Figure 3.**
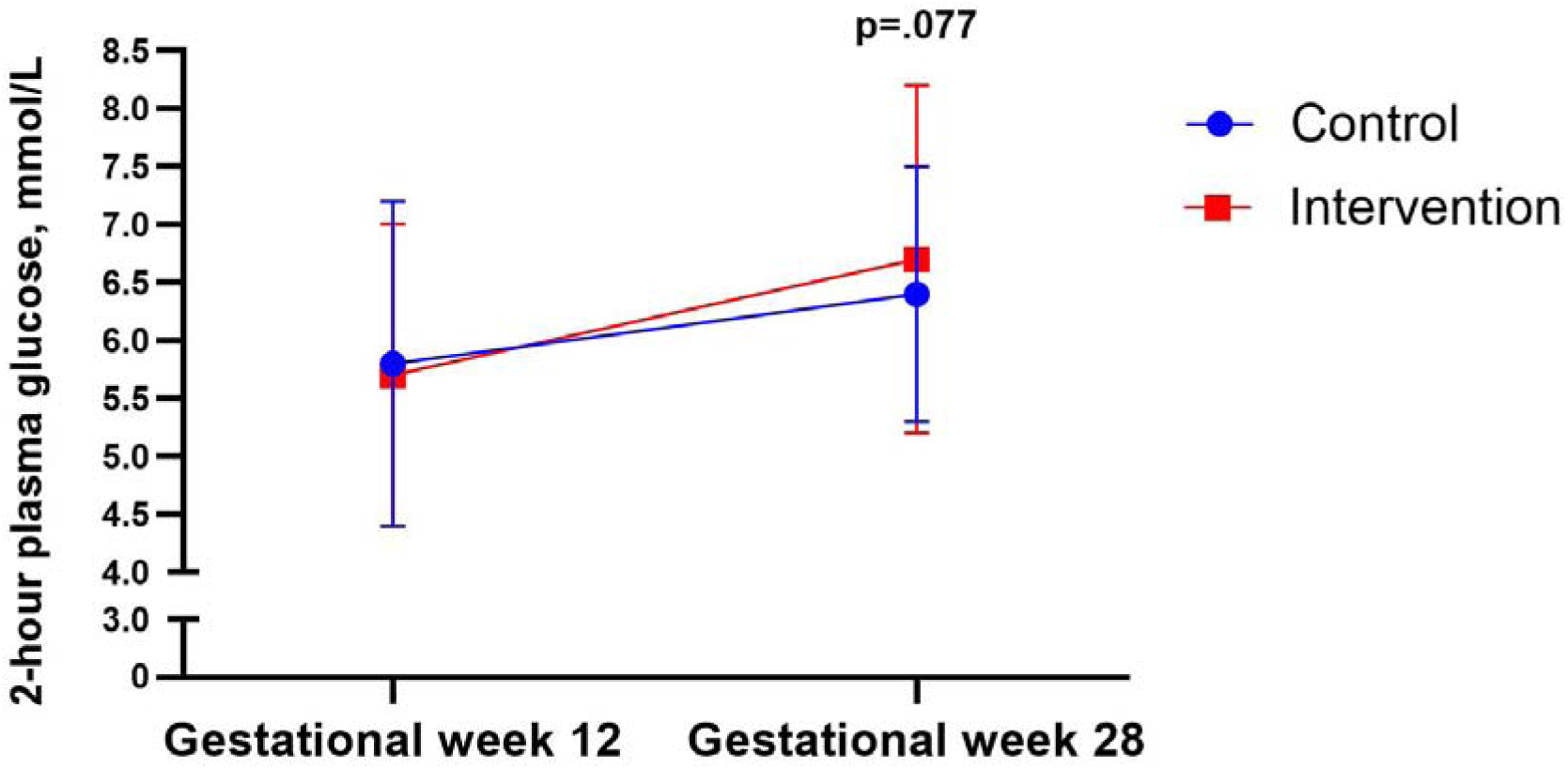
Plasma glucose 2 hours after a 75-g oral glucose tolerance test in gestational weeks 12 and 28, according to group. Data are observed means and standard deviations for the intention-to-treat population. The p-value was calculated for a test of between-group differences, using a linear mixed model.

**Table 2.** Intention-to-treat analyses. Data are observed means and standard deviations (SD) at baseline, in pre-pregnancy week 8, gestational week 12, and gestational week 28 for n participants in each group. Results from linear mixed model analyses are presented as estimated mean difference (Est. effect) in the intervention group compared with the control group, with corresponding 95% confidence interval (CI) and p-values. *95% CI and p-values are from bias-corrected and accelerated confidence intervals based on bootstrap with 3000 samples, due to non-normally distributed residuals.

|  |  | Control<br>(*n* = 54) | Intervention<br>(*n* = 55) | Difference (intervention - control) |  |  |
| --- | --- | --- | --- | --- | --- | --- |
| Outcome | Time | Mean (SD) | Mean (SD) | Est.<br>effect | 95% CI | *p* |
| **Primary outcome** |  |  |  |  |  |  |
| 2-hour plasma glucose during OGTT, mmol/L | Gestational week 12 | 5.8 (1.4) | 5.7 (1.3) |  |  |  |
|  | Gestational week 28 | 6.4 (1.1) | 6.7 (1.5) | 0.48 | -0.05 to 1.01 | .077 |
| **Secondary outcomes** |  |  |  |  |  |  |
|  |  | Control<br>(*n* = 83) | Intervention<br>(*n* = 83) | Difference (intervention – control) |  |  |
| Outcome | Time | Mean (SD) | Mean (SD) | Est.<br>effect | 95% CI | *p* |
| Fasting blood glucose, mmol/L | Baseline | 5.0 (0.4) | 4.9 (0.4) |  |  |  |
|  | Pre-pregnancy week 8 | 5.1 (0.4) | 5.0 (0.3) | 0.0 | -0.2 to 0.1 | .392 |
|  | Gestational week 12 | 4.5 (0.3) | 4.5 (0.3) | 0.0 | -0.1 to 0.1 | .640 |
|  | Gestational week 28 | 4.5 (0.4) | 4.5 (0.5) | 0.0 | -0.1 to 0.2 | .427 |
| HbA1c, mmol/mol | Baseline | 34.3 (3.1) | 34.1 (2.8) |  |  |  |
|  | Pre-pregnancy week 8 | 34.5 (3.2) | 33.9 (2.5) | -0.4 | -1.1 to 0.2 | .205 |
|  | Gestational week 12 | 32.7 (2.3) | 33.3 (2.4) | 0.5 | -0.2 to 1.2 | .140 |
|  | Gestational week 28 | 32.3 (2.8) | 32.0 (3.19) | -0.2 | -0.9 to 0.5 | .516 |
| Fasting insulin, µU/mL* | Baseline | 19.1 (11.7) | 17.1 (10.4) |  |  |  |
|  | Pre-pregnancy week 8 | 19.3 (9.7) | 16.3 (8.6) | -2.0 | -4.4 to 0.5 | .123 |
|  | Gestational week 12 | 12.8 (10.7) | 12.3 (9.1) | 0.1 | -3.8 to 4.0 | .960 |
|  | Gestational week 28 | 19.4 (12.2) | 18.6 (11.0) | -0.8 | -5.0 to 3.4 | .716 |
| HOMA2-B* | Baseline | 167.8 (58.9) | 164.9 (69.2) |  |  |  |
|  | Pre-pregnancy week 8 | 171.2 (59.2) | 155.0 (51.6) | -13.8 | -30.1 to 2.5 | .097 |
|  | Gestational week 12 | 153.8 (81.4) | 155.8 (68.7) | 4.6 | -24.1 to 33.2 | .755 |
|  | Gestational week 28 | 212.4 (84.9) | 205.2 (70.0) | -7.7 | -35.1 to 19.6 | .579 |
| HOMA2-IR* | Baseline | 2.4 (1.4) | 2.2 (1.2) |  |  |  |
|  | Pre-pregnancy week 8 | 2.4 (1.2) | 2.1 (1.1) | -0.2 | -0.5 to 0.1 | .213 |
|  | Gestational week 12 | 1.6 (1.2) | 1.5 (1.1) | 0.1 | -0.4 to 0.5 | .787 |
|  | Gestational week 28 | 2.3 (1.4) | 2.3 (1.3) | 0.0 | -0.5 to 0.5 | .878 |
| Total Cholesterol, mmol/L | Baseline | 4.6 (0.7) | 4.6 (0.8) |  |  |  |
|  | Pre-pregnancy week 8 | 4.5 (0.7) | 4.7 (0.8) | 0.3 | 0.0 to 0.5 | .027 |
|  | Gestational week 12 | 4.4 (0.7) | 4.4 (0.8) | 0.0 | -0.3 to 0.2 | .811 |
|  | Gestational week 28 | 5.9 (1.2) | 5.8 (1.1) | -0.1 | -0.3 to 0.2 | .574 |
| LDL cholesterol, mmol/L | Baseline | 3.0 (0.9) | 3.1 (0.8) |  |  |  |
|  | Pre-pregnancy week 8 | 2.9 (0.8) | 3.1 (0.8) | 0.2 | 0.0 to 0.4 | .089 |
|  | Gestational week 12 | 2.8 (0.7) | 2.8 (0.8) | 0.0 | -0.3 to 0.2 | .844 |
|  | Gestational week 28 | 3.9 (1.2) | 3.9 (1.1) | -0.1 | -0.4 to 0.1 | .398 |
| HDL cholesterol, mmol/L | Baseline | 1.4 (0.3) | 1.4 (0.3) |  |  |  |
|  | Pre-pregnancy week 8 | 1.4 (0.3) | 1.5 (0.3) | 0.0 | 0.0 to 0.1 | .376 |
|  | Gestational week 12 | 1.6 (0.4) | 1.6 (0.2) | 0.0 | -0.1 to 0.1 | .494 |
|  | Gestational week 28 | 1.8 (0.4) | 1.8 (0.3) | 0.0 | -0.1 to 0.1 | .816 |
| Triglycerides, mmol/L | Baseline | 1.0 (0.5) | 0.9 (0.5) |  |  |  |
|  | Pre-pregnancy week 8 | 1.0 (0.5) | 0.9 (0.4) | 0.0 | -0.2 to 0.1 | .826 |
|  | Gestational week 12 | 1.1 (0.4) | 1.1 (0.4) | 0.0 | -0.1 to 0.2 | .545 |
|  | Gestational week 28 | 2.0 (0.7) | 1.9 (0.6) | 0.0 | -0.2 to 0.1 | .738 |
| Weight, kg | Baseline | 81.5 (13.2) | 81.1 (16.0) |  |  |  |
|  | Pre-pregnancy week 8 | 81.6 (12.5) | 78.9 (14.8) | -0.9 | -2.0 to 0.3 | .145 |
|  | Gestational week 12 | 79.5 (12.9) | 78.8 (15.2) | -0.9 | -2.2 to 0.3 | .152 |
|  | Gestational week 28 | 86.8 (12.3) | 85.1 (15.4) | -2.0 | -3.3 to -0.8 | .002 |
| Fat mass, kg | Baseline | 31.6 (9.8) | 30.8 (11.0) |  |  |  |
|  | Pre-pregnancy week 8 | 31.2 (9.5) | 28.8 (9.7) | -0.8 | -1.8 to 0.2 | .103 |
|  | Gestational week 12 | 30.6 (9.5) | 29.5 (10.7) | -0.5 | -1.5 to 0.6 | .410 |
|  | Gestational week 28 | 34.2 (9.0) | 32.5 (10.8) | -1.5 | -2.5 to -0.4 | .008 |
| Fat percentage, % | Baseline | 37.9 (7.2) | 37.0 (7.6) |  |  |  |
|  | Pre-pregnancy week 8 | 37.6 (7.0) | 35.7 (6.6) | -0.6 | -1.4 to 0.2 | .144 |
|  | Gestational week 12 | 37.7 (7.0) | 36.5 (7.4) | -0.2 | -1.0 to 0.7 | .734 |
|  | Gestational week 28 | 38.8 (6.0) | 37.4 (7.2) | -0.7 | -1.5 to 0.2 | .139 |
| Visceral fat area, cm<sup>2</sup> | Baseline | 153.6 (52) | 146 (57) |  |  |  |
|  | Pre-pregnancy week 8 | 151 (50) | 135 (50) | -5.1 | -10.9 to 0.7 | .086 |
|  | Gestational week 12 | 148 (51) | 136 (54) | -2.3 | -8.6 to 4.1 | .483 |
|  | Gestational week 28 | 165 (47) | 153 (54) | -6.2 | -12.5 to 0.1 | .054 |
| Muscle mass, kg | Baseline | 27.7 (3.6) | 27.9 (4.3) |  |  |  |
|  | Pre-pregnancy week 8 | 28.0 (3.6) | 27.8 (4.2) | 0.0 | -0.3 to 0.3 | .855 |
|  | Gestational week 12 | 27.0 (3.2) | 27.2 (4.1) | -0.2 | -0.5 to 0.2 | .316 |
|  | Gestational week 28 | 29.1 (3.1) | 29.1 (4.2) | -0.3 | -0.7 to 0.0 | .061 |
| Waist circumference, cm | Baseline | 94.5 (11.8) | 93.0 (12.3) |  |  |  |
|  | Pre-pregnancy week 8 | 93.9 (11.2) | 92.0 (11.5) | -0.5 | -2.9 to 1.8 | .663 |
|  | Gestational week 12 | 95.1 (11.5) | 94.9 (12.1) | 0.6 | -1.8 to 3.1 | .610 |
|  | Gestational week 28 | 107.3 (8.4) | 105.9 (10.6) | -1.5 | -4.0 to 1.0 | .243 |
| Systolic blood pressure, mmHg | Baseline | 121 (10) | 118 (8) |  |  |  |
|  | Pre-pregnancy week 8 | 119 (8) | 117 (8) | 0.2 | -2.4 to 2.7 | .901 |
|  | Gestational week 12 | 114 (10) | 110 (8) | -2.2 | -5.0 to 0.6 | .122 |
|  | Gestational week 28 | 110 (9) | 110 (11) | 0.6 | -2.3 to 3.4 | .688 |
| Diastolic blood pressure, mmHg | Baseline | 79 (7) | 79 (6) |  |  |  |
|  | Pre-pregnancy week 8 | 77 (6) | 77 (6) | 0.0 | -2.1 to 2.0 | .973 |
|  | Gestational week 12 | 71 (8) | 70 (6) | -0.9 | -3.1 to 1.3 | .414 |
|  | Gestational week 28 | 69 (8) | 70 (8) | 0.5 | -1.7 to 2.8 | .647 |
| Resting heart rate, beats per minute | Baseline | 72 (11) | 71 (11) |  |  |  |
|  | Pre-pregnancy week 8 | 69 (9) | 67 (10) | -1.7 | -4.8 to 1.4 | .288 |
|  | Gestational week 12 | 71 (11) | 69 (15) | -1.9 | -5.4 to 1.5 | .275 |
|  | Gestational week 28 | 78 (12) | 77 (13) | -0.9 | -4.3 to 2.6 | .629 |

OGTT = Oral glucose tolerance test, HbA1c = Glycated haemoglobin, HOMA2-B = Homeostatic model assessment of beta cell function, HOMA2-IR = Homeostatic model assessment of insulin resistance, HDL = High-density lipoprotein cholesterol, LDL = Low-density lipoprotein cholesterol.

#### Secondary outcomes

The estimated mean weight gain in the intervention group in gestational week 28 was 2.0 kg lower (95% CI -3.3 to -0.8, *p* = .002), and fat mass gain was 1.5 kg lower (95% CI to -2.5 to - 0.4, *p* = .008), than in the control group (Table 2). There was little or no evidence of other between-group differences during pregnancy.

#### Adherence

At baseline, the average daily eating window for all participants was 11.9 (SD 1.7) hours. The eating window was shorter in the intervention group during the rest of the study period, compared with the control group (Figure 4). In the pre-pregnancy period, the intervention group had an average eating window of 9.9 (SD 1.2) hours, which increased during pregnancy (Figure 4 and Supplementary file 2). The intervention group significantly increased their activity levels during the pre-pregnancy phase compared with baseline. The average weekly PAI-points decreased throughout pregnancy and their physical activity levels during pregnancy did not differ significantly from baseline (Figure 4 and Supplementary file 2).

**Figure 4.**
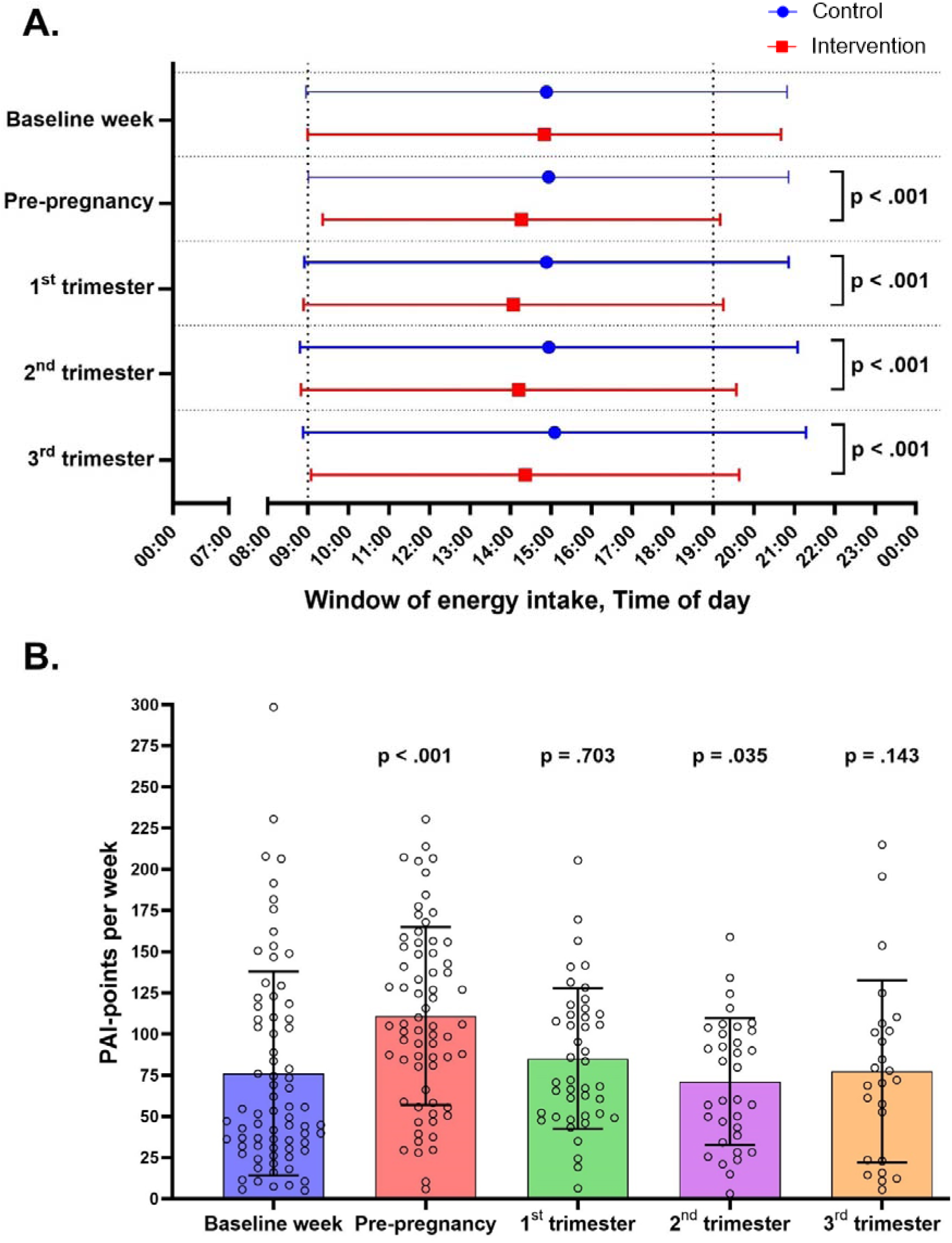
Adherence to the intervention. (A) Time-window of energy intake during the baseline week, pre-pregnancy period, and 1^st^, 2^nd^, and 3^rd^trimesters of pregnancy, according to group. The horizontal lines represent the daily duration of energy intake, with the left and right ends indicating means of the time of first and last energy intake for the intention-to-treat population. The p-values were calculated for a test of between-group differences in the window of energy intake, using linear mixed model. (B) Personal Activity Intelligence (PAI)-points earned per rolling week during baseline week, pre-pregnancy period, and first, second and third trimesters of pregnancy, in the intervention group. The bars show mean PAI-points per week, error bars show standard deviation, and symbols show individual data. The p-values were calculated for within-group differences from baseline, estimated by a linear mixed model.

#### Per protocol analyses

In the per-protocol analyses, we included only the participants in the intervention group who obtained ≥ 75 weekly PAI-points and reported a ≤ 10-hour time window for energy intake on ≥ 2 out of 4 days in the handbook in the pre-pregnancy period. Thirty-one (37%) participants satisfied both criteria and were included in the analyses. The results from the per-protocol analyses were similar to those from the intention-to-treat analyses, except for time to pregnancy which was significantly longer in the intervention group (49 days, 95% CI 16 to 82, *p* = .004) (Supplementary File 3).

#### Dietary intake and physical activity

There was no statistically significant difference in self-reported total energy intake in the intervention group compared with the control group pre-pregnancy (-57.0 kcal/day, 95% CI - 183.9 to 69.9, *p* = .378) or during the first trimester (41.2 kcal/day, 95% CI -92.6 to 175.1, *p* = .545), second trimester (39.7 kcal/day, 95% CI –101.4 to 180.9, *p* = .580), or third trimester (-110.3 kcal/day, 95% CI -248.0 to 27.3, *p* = .116). Nor were there any significant differences in the distribution of macronutrients in the intervention group compared with the control group before or during pregnancy (Supplementary file 2). Self-reported physical activity level did not differ significantly between groups before or during pregnancy (Supplementary file 2).

#### Adverse events

No serious adverse events were reported during the study. Minor events included some participants feeling dizzy and/or nauseous during fasting blood sampling and OGTT. Water intake and fresh air solved this problem for most participants, although some discontinued blood sampling. Some participants experienced local skin irritation from wearing a continuous glucose monitor, requiring premature removal. Others reported local skin irritation from the physical activity monitor armbands and smartwatches, which was resolved by repositioning the monitors or switching to metal wristwatch armbands.

## Discussion

### Main findings

The BEFORE THE BEGINNING study is to our knowledge the first to implement a combination of TRE and exercise training before conception and throughout pregnancy in people at increased risk of developing GDM. Contrary to our hypothesis, we found no significant between-group differences in 2-hour plasma glucose level after a 75 g glucose load in gestational week 28, or in secondary outcome measures of glycaemic control at any time-points before or during pregnancy. Despite that the intervention reduced body weight and fat mass gain in gestational week 28, there was no significant effect on GDM incidence. The participants were able to adhere to the ≤ 10-hour TRE intervention pre-pregnancy, with a slight increase in the time-window of energy intake during pregnancy. While participants in the intervention group earned more than 100 PAI during pre-pregnancy, their activity levels declined during pregnancy, and there were no significant between-group differences in self-reported energy intake or physical activity throughout the study period.

### Strengths and limitations

The main strengths of the BEFORE THE BEGINNING trial are the randomised controlled design and longitudinal measurements throughout the pre-pregnancy and pregnancy periods. The study had a high pregnancy rate (66% in both groups), and a low dropout rate during pregnancy (7% in both groups). Still, the study has several limitations. Most participants were highly educated, potentially resulting in healthy volunteer bias as they were likely knowledgeable about their health and well-being.(44) Most of our participants were also white and thus it is difficult to generalize our findings to other ethnicities. However, recruitment via social media is thought to give a better representation of the general population than other recruitment methods.(45) Since we had a low number of participants (*n* = 3) with previous GDM, adjustment for stratification variable (GDM) was not included in the analyses. Despite good adherence to TRE and exercise training in the pre-pregnancy period, we did not observe any improvements in cardiometabolic outcomes compared with the control group. One possible reason for this could be the variable length of the pre-pregnancy period, as the time to pregnancy varied greatly among participants. Additionally, the number of PAI-points earned during the baseline week also varied greatly within the participants in the intervention group and some of the participants had already a high physical activity level at baseline. Self-reported activity levels did not differ significantly between groups, suggesting that the level of physical activity in the control group was similar as in the intervention group.

### Comparisons with other studies

Most lifestyle interventions for GDM prevention begin during pregnancy, missing the crucial window of opportunity in the pre-pregnancy period to improve cardiometabolic health outcomes in at-risk individuals.(46) Systematic reviews and meta-analyses have highlighted the lack of randomised controlled trials focusing on pre-pregnancy lifestyle interventions in this population.(47,48) To date, only a handful of studies have investigated the role of pre-pregnancy lifestyle intervention continued throughout pregnancy on cardiometabolic outcomes.(49–53) We found no significant effect of the intervention on glucose tolerance in gestational week 28, or on any other glycaemic indices. In contrast to our hypothesis, we estimated a *higher* mean 2-hour plasma glucose concentration in the intervention group in gestational week 28 according to both the ITT and the per-protocol analysis (0.48 and 0.64 mmol/L, respectively), compared with the control group. However, the estimated mean differences were smaller than what was considered a clinically relevant difference (1.0 mmol/L). A slight, non-significant, increase in 2-hour plasma glucose concentration (0.2 mmol/L) was also observed after a 5-week TRE intervention during pregnancy.(27)

In contrast to our findings, Price et al.(50) reported that a 12-week pre-pregnancy very low-energy diet intervention significantly reduced 2-hour glucose during a 75 g OGTT by 0.8 mmol/L compared with a standard diet intervention group among women with BMI between 30 and 55 kg/m^2^. In that study, the participants in the intervention group consumed approximately 800 kcal/day for 12 weeks, followed by a maintenance period of energy expenditure-matched energy intake throughout the pre-pregnancy period, while also being advised to remain physically active (> 10,000 steps/day). The 12-week pre-pregnancy intervention induced a 9.2-kg weight loss compared with the standard diet, but gestational weight gain remained unaffected.(50) Based on the findings form Price et al(50) and our study, it seems likely to conclude that a pre-pregnancy intervention must induce a substantial weight loss to impact glucose tolerance in pregnancy. Our trial, along with others(49–51,53), did not find any significant effect of pre-pregnancy lifestyle interventions on GDM incidence in gestational week 28. Despite being at higher risk of GDM, with 86% of the participants having BMI above 25 kg/m^2^ and 27% having a family history of diabetes, only 12% of our participants were diagnosed with GDM in gestational week 28. The GDM incidence in our trial was substantially lower than in the RADIEL study (60% in the control group and 54% in the intervention group)(49) and in the study by Phelan et al. (40% in the control group and 25% in the intervention group).(53) The low incidence of GDM observed in our study may be attributed to the high education level (87%) and mostly white participants (89%)(54). Importantly, all our participants included in the analyses were normoglycaemic at baseline, which may help explain the lack of effect since individuals with impaired glucose metabolism benefit the most from TRE.(55)

The intervention group had significantly lower body weight gain (2.0 kg) in gestational week 28, compared with the control group, but there were no significant differences between groups in the pre-pregnancy period or the first trimester of pregnancy. In contrast, some of the previous pre-pregnancy lifestyle interventions induced pre-pregnancy weight loss, but did not affect weight gain during pregnancy.(49,50,52,53) In the PREPARE study, the intervention group lost weight before pregnancy, followed by a greater weight gain in late pregnancy.(52) The addition of exercise training to the TRE intervention in our study may have prevented lean mass loss which is common in TRE interventions.(25) Although not statistically significant, the estimated mean visceral fat area was lower in the intervention group compared with the control group both before and during pregnancy. The amount of visceral fat in early pregnancy has been shown to better predict GDM than pre-pregnancy BMI.(56) However, the observed changes in body composition may not have been large enough to improve the other cardiometabolic outcomes in our study.

There were no significant between-group differences in total energy intake or macronutrient distribution before or during pregnancy, which probably partly explain the neutral effect of the intervention on most outcome measures. In contrast, Haganes et al.(23) reported that 7 weeks of combined TRE and HIIT induced a reduction in energy intake of approximately 200 kcal/day compared with the control group, along with improvements in several cardiometabolic outcomes, among reproductive-aged women with overweight/obesity. Correspondingly, the combination of TRE and high-intensity functional training for 12 weeks resulted in a 175 kcal/day reduction in total energy intake among women with obesity.(57) In BEFORE THE BEGINNING, we chose a modified TRE regimen, allowing for unrestricted intake on 2 days per week. Even if such a regimen allows for more flexibility and potentially improved long-term adherence than a stricter TRE intervention, our TRE intervention was not sufficiently potent to reduce energy intake. Participants in dietary interventions often underreport their total energy intake(58), including when using the electronic application that we used.(59) Still, there is no reason to believe that such under-reporting would be different between the intervention and control group.

There is thus far little research on the safety and acceptability of TRE during pregnancy, and only one previous randomised controlled trial on the effects of TRE on glycaemic control.(27) In a recent online survey study, only around half of the participants agreed that a TRE pattern is safe during pregnancy and 23.7% would be willing to try TRE during pregnancy to improve their health.(60) There was a gradual decrease in the adherence to TRE in our study, from pre-pregnancy and throughout pregnancy. In our previous study of TRE in pregnancy,(27) the participants could adhere to a 10-hour TRE intervention on ∼ 5 days/week for 5 weeks in the second or third trimester of pregnancy. However, in that study, the trial period was markedly shorter, and the participants were already pregnant at inclusion, thus likely less bothered by nausea and other barriers to TRE than in the BEFORE THE BEGINNING study. Collectively, the experimental evidence to date on TRE during pregnancy indicates that there is no positive effect on maternal glycaemic control.

HIIT is now considered safe and feasible during pregnancy.(29–31) However, the long-term adherence to HIIT interventions during pregnancy remains to be explored. In our study, the participants in the intervention group significantly increased their physical activity levels in the pre-pregnancy period with a decrease during pregnancy, suggesting suboptimal adherence to the exercise component of the intervention. Only 43% of the participants in the intervention group met the goal of 100 weekly PAI-points pre-pregnancy, decreasing gradually to 10% of participants during the third trimester. Although self-reported physical activity levels did not differ significantly between the groups, the reported physical activity levels were higher in the intervention group throughout the study. Adherence to exercise training is typically lower in unsupervised compared with supervised situations.(61) The declining adherence to exercise during the study period was likely due to a combination of the unsupervised nature of the intervention, pregnancy-related side effects (e.g., nausea, pelvic pain), and decreasing motivation over a long study period. Overall, adherence to lifestyle interventions in pregnancy remains a significant challenge, especially in real-life settings without close supervision.

### Conclusion and implications for clinicians and policymakers

The combination of TRE and exercise training, initiated before and continued throughout pregnancy, had limited effects on glucose tolerance in late pregnancy among people with an increased risk of GDM. Despite the challenges with adherence to the intervention, the intervention group had lower weight and fat mass gain in gestational week 28. Emerging evidence highlights that the pre-pregnancy period is an ideal time to intervene in people at high risk of cardiometabolic diseases and it is critical to find optimal strategies to improve adherence to lifestyle interventions in this population.

### Unanswered questions and future research

We experienced challenges with adherences to the intervention, particularly to the exercise training component in pregnancy. Future studies should assess whether more organised high-intensity exercise training during pregnancy lead to better long-term exercise adherence. Using more interactive digital health technology (e.g., smartphone apps, automatically delivered daily reminders, weekly goals) should also be explored. The per-protocol analysis indicated longer time to pregnancy in the intervention group, which should be further investigated. Not all participants in our study had high BMI (some were included based on the other risk factors), and therefore, there were variations in body composition and cardiometabolic markers. Future studies should also determine whether interventions based on single risk factors (e.g., high BMI, previous GDM) to avoid large within-group variations, lead to better cardiometabolic outcomes.

### What is already known on this topic

- High pre-pregnancy BMI and excessive gestational weight gain are associated with a greater risk of developing GDM and adverse pregnancy and neonatal outcomes.
- Systematic reviews and meta-analyses suggest that pre-pregnancy lifestyle interventions are necessary to improve maternal and fetal cardiometabolic outcomes.
- Existing guidelines on diet and physical activity for pregnant individuals are insufficient to achieve clinically significant cardiometabolic benefits.

### What this study adds

- The BEFORE THE BEGINNING trial is the first trial to investigate the effect of combined TRE and exercise training before and during pregnancy in people at high risk of GDM.
- The intervention did not result in significantly improved glycaemic outcomes.
- There was some evidence of lower weight and fat mass gain in gestational week 28 in the intervention group compared with the control group.

## Ethical approval

The Regional Committees for Medical and Health Research Ethics in Norway approved the study (REK 143756).

## Data availability statement

Data reported in this paper can be shared by the corresponding author upon request.

## Acknowledgements

We are thankful to all the participants for their valuable contributions. NeXt Move, Norwegian University of Science and Technology (NTNU) provided the equipment and lab facilities for cardiorespiratory fitness testing, and the clinical measurements are obtained at the Clinical Research Facility, St. Olavs Hospital. We used eFORSK, a stand-alone form-based information and communications technology solution for electronic data collection, developed by Central Norway Regional Health Authority for sending invitations to the study.

## Footnotes

TM, KÅS, TF, and SLF conceived and contributed to the study design and data analysis plans. GR, MAJS, and HMSS coordinated the study, performed measurements, monitored participants, and supervised the exercise training. MAJS and HMSS drafted the manuscript with equal contribution. All authors provided feedback and approved the final manuscript.

## Funding

The trial is funded by the Novo Nordisk Foundation (NNF19SA058975), The Liaison Committee for education, research, and innovation in Central Norway, and The Joint Research Committee between St. Olavs Hospital and the Faculty of Medicine and Health Sciences, NTNU (FFU). The sponsors have no role in study design, data collection, analysis, and publication of results.

## Declaration of interests

The authors declare that they have no competing interests.

## Transparency

The authors confirm that this manuscript is an honest, accurate, and transparent account of the study being reported; that no important aspects of the study have been omitted; and that any discrepancies from the study as planned and registered have been explained.

## Dissemination to participants and related patient and public communities

We plan to disseminate all the results to study participants and patient organisations as soon as the study is published. The result of this trial will be presented at relevant national and international conferences.

## Notes

### Competing Interest Statement

The authors have declared no competing interest.

### Clinical Trial

NCT04585581

### Clinical Protocols

https://bmjopen.bmj.com/content/13/10/e073572.info

