## Supplementary file 1 for "Time-restricted eating and exercise training before and during pregnancy for people with increased risk of gestational diabetes: the BEFORE THE BEGINNING randomised controlled trial"

### Exercise Options

#### EXERCISE IN PREGNANCY

Good condition and strong muscles, joints and skeleton provide better conditions for a healthy pregnancy. During pregnancy, your workout should contain the same elements as before the pregnancy, but with a slightly lower intensity level.

Avoid any activities that has a lot of jerky, bouncing movements that may cause you to fall, like horseback riding, downhill skiing, off-road cycling, or gymnastics. Also, avoid sports in which you may be hit in the belly, such as ice hockey, boxing, soccer, or basketball.

Below you will find suggestions for workouts and exercise programs both before - and during pregnancy. These are only suggestions, and you decide for yourself what kind of physical activity you want to do. We are different and have different preferences for which activities are pleasurable and gives you joy.

#### WHAT IS TRAINING INTENSITY?

Training intensity is the amount of effort you are putting into whatever exercise you are doing. You can subjectively rate your level of exertion during exercise by using Borg scale. The figure below will help you evaluate where on the scale you are during exercise. Vigorous-intensity exercise is a physical activity done with a large amount of effort, resulting in a substantially higher heart rate and rapid breathing. Engaging in vigorous physical activity can provide many health advantages. Thus, it can be useful to know the distribution between light -, moderate – and vigorous intensity during your workouts. Evaluate the intensity while doing the activity. Try to be as sincere as possible. Remember that this is a subjective scale – what others thinks is irrelevant. Use the full scale and choose the number that suits your experience during activities best.

##### Borg`s Rating of Perceives Exertion (RPE) Scale

| Value | Perceived Exertion Rating | Description of Exertion |
|---|---|---|
| 6 | No exertion | Rest |
| 7 | Extremely light | |
| 8 | | |
| 9 | Very light | Comfortable walking pace. Conversation is easy. |
| 10 | | |
| 11 | Light | |
| 12 | | |
| 13 | Somewhat hard | You feel you could run/walk for a while at this pace. Still able to talk. |
| 14 | | |
| 15 | Hard | Hard, but you`re not struggling. You can talk but not in full sentences. |
| 16 | | |
| 17 | Very hard | Very hard – Starting to get uncomfortable and you`re getting tired. |
| 18 | | |
| 19 | Extremely hard | Extremely hard. Your body is screaming at you to stop. You can no longer talk because your breathing is heavy. |
| 20 | Maximal exertion | Max exertion. |

**7-11 = Light** intensity **12-15 = Moderate** intensity **16-20 = Vigorous** intensity

### PAI

To measure your activity level, we use the science-backed health score called PAI (Personal Activity Intelligence). You earn PAI points every time your heart rate increases: The higher your heart rate, the faster you earn PAI. Research shows that those who achieve 100 PAI or more every week over time live on average eight years longer than others.

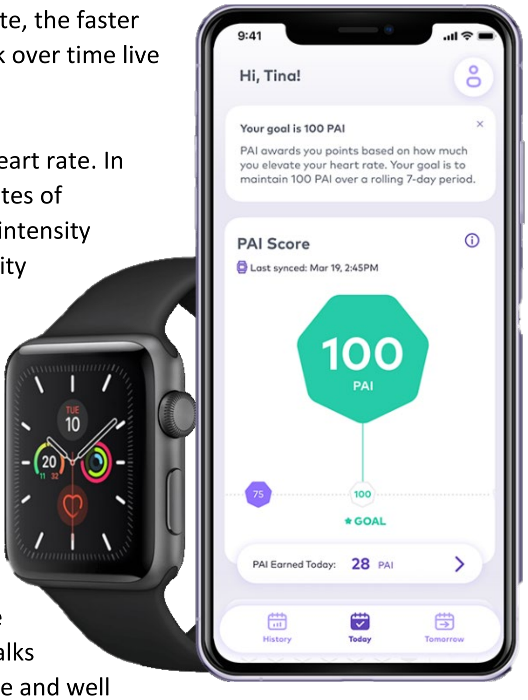

PAI considers your age, your gender, your resting heart rate and your maximum heart rate. In other words, PAI is not based on the number of steps you walk or how many minutes of physical activity you perform each day. Those measurements do not consider the intensity of the activity. Therefore, we think that PAI is a more accurate and attractive activity standard to tell you if you actually exercise enough.

**Your goal is to earn 100 PAI or more during a 7-day period**. It doesn't matter what type of activity you do to earn PAI – you could walk, run, cycle, row, swim or go skiing. All that matters is how high your heart rate is during the activity.

PAI is based on the only thing that reflects the intensity of your activity: your heart rate. Everything you need to do in order to use PAI is to measure your heart rate continuously. The better your fitness is, the more physical activity is needed to achieve 100 PAI. In other words, PAI adjusts to your progress. If you are untrained and out of shape, you could earn your 100 PAI just by going for short walks regularly throughout the week, as that will raise your heart rate. If you are in shape and well trained, you will need to do more.

It is easier to reach the first 50 PAI compared to the next 50. That is because the risk reduction for lifestyle diseases is greatest when progressing from total inactivity to some physical activity. This means that if you repeat a workout two days in a row, you will get less PAI on the second workout.

Remember to turn on the **“workout-mode”** during exercise.

### CONDITION: BEFORE PREGNANCY

Endurance sessions can be done both outside and indoors. The sessions can be done with any endurance equipment: Bicycle, treadmill, skiers, rowing machine, assault bike or elliptical machine.

| INTERVAL 1 | |
|---|---|
| **What?** | **Duration** |
| **Warm-up:** 5 minutes with low intensity (Borg scale: 7-11), thereafter 5 minutes with moderate intensity (Borg scale 12-15).<br><br>**Interval:** 4 intervals x 4 min with high intensity (Borg scale 16-20). 3 minutes active break after each interval.<br><br>**Cool-down:** 5 minutes with low intensity (Borg scale: 7-11). | 40 min |

| INTERVAL 2 | |
|---|---|
| **What?** | **Duration** |
| **Warm-up:** 5 minutes with low intensity (Borg scale: 7-11), thereafter 5 minutes with moderate intensity (Borg scale 12-15).<br><br>**Interval:** 10 intervals x 1 min with high intensity (Borg scale: 16-20). 1-minute active break after each interval.<br><br>**Cool-down:** 5 minutes with low intensity (Borg scale: 7-11). | 35 min |

### TABATA INTERVALS: BEFORE PREGNANCY

Tabata is a type of HIIT workout combining high intensity with strength training. Tabata is a four-minute workout consisting of 8 rounds of 20 seconds of work at maximum effort, followed by 10 seconds of rest. Between each station you have a total of 30 sec of rest. Target heart rate should be between 85-95 % of your maximum heart rate (Borg scale: 16-20). You should not be able to talk because your breathing is heavy.

**Warm-up:** 10 minutes with low intensity (Borg scale: 7-11) on endurance equipment of choice.
**Cool-down:** 5 minutes with low intensity (Borg scale: 7-11) on endurance equipment of choice.

| **TABATA** | | | |
|---|---|---|---|
| **Equipment**: Kettlebells, yoga mat, bench/step/box, weight plates or dumbbells | | | |
| **Exercises** | **Muscle group** | **Comment** | **Duration** |
| **Station 1:** Walking burpees | Thighs and gluteal muscle | Begin standing with your feet hip-distance apart (A). Then, squat down and step one foot back at a time (B). After you're in the plank position (C), walk your feet back to meet your hands and return to standing. That's one rep. | 34 min |
| **Station 2:** Reverse crunches | Stomach muscles | Start lying down with your arms by your sides. Raise your legs so your thighs are perpendicular to the floor and your knees are bent at a 90° angle. Breathe out and contract your abs to bring your knees up towards your chest and raise your hips off the floor. Hold for a beat in this position, then slowly lower your legs back to the starting position. | |
| **Station 3:** Bent over row w/kettlebells | Back and shoulders | With a kettlebell in each hand, bend over at about a 45-degree angle (no farther). Keep the back straight throughout the exercise. Brace your abdominals and breathe in. Lift the weights straight up, exhaling. While lifting, the arms should go no higher than parallel with the shoulders—slightly lower than the shoulders is fine. While lifting, try to keep the wrists from excessive extra movement down or to the side. Do not squat down and up after the initial pose. No movement of the legs occurs throughout the exercise. Lower the weights in a controlled manner while inhaling. Remain bent over until all repetitions are complete. | |
| **Station 4:** Push-ups<br><br>Modifications:<br>• Push-ups against a wall or bench, or on your knees | Chest and arms. | Get down on all fours, placing your hands slightly wider than your shoulders.<br>Straighten your arms and legs.<br>Lower your body until your chest nearly touches the floor/wall/bench. Pause, then push yourself back up.<br>Repeat. | |
| **Station 5:** Farmers walk w/kettlebells | Thighs, gluteal muscle, calves, back, neck, arms and stomach muscles. | Pick up a pair of kettlebells in each hand (crush the handles). Stand tall with kettlebells out to the sides a couple of inches (do not let the weight touch your legs)<br>Look straight ahead and pinch your shoulder blades (scapula) back slightly. Keep your core tight. Walk slowing and with small steps. | |
| **Station 6:** Biceps curl and shoulder press w/dumbbells | Arms and shoulders | Holding a pair of dumbbells, stand tall with your feet shoulder-width apart. Make sure your core is tight and your chest is up. Begin by curling the weight up towards your shoulders. Keep your upper arms tight at your sides.<br>Once the dumbbells reach your shoulders, twist the dumbbells to have your palms face out. Now, drive the dumbbells overhead. Slowly, lower the dumbbells to your shoulders. Now, flip them back so your palms are facing you. With arms tight at your sides, lower the dumbbells to the starting position. | |
| **Station 7:** Box step-ups | Thighs and gluteal muscle | Plant your right foot on the box, lean forward and step up so you're standing with both feet on the box. Then, step back with your right foot and plant it on the ground. Then, step back with your left foot. Next, step up with your left foot followed by the right foot. Each repetition you'll switch feet. | |
| **Stations 8:** Russian twist | Stomach muscles | Sit on your sit bones as you lift your feet from the floor, keeping your knees bent. Elongate and straighten your spine at a 45-degree angle from the floor, creating a V shape with your torso and thighs.<br>Reach your arms straight out in front, interlacing your fingers or clasping your hands together. Use your abdominals to twist to the right, then back to center, and then to the left. This is 1 repetition. If you want, you can hold a dumbbell, weight plate between both hands. | |

### EXERCISE PROGRAM: BEFORE PREGNANCY

What your weekly training plan could look like:

| **EXERCISE PROGRAM** | | | |
|---|---|---|---|
| **Day** | **What?** | **Intensity (Borg scale)** | **Duration** |
| Monday | **Warm-up:** 5 minutes with low intensity, thereafter 5 minutes with moderate intensity.<br>**Intervals:** 4 x 4 min intervals uphill. 3 min rest between each interval. If you choose to do the intervals outside, I recommend finding a long and slight hill. The break will be walking down to the bottom of the hill.<br>**Cool-down**: 5 min with low intensity. | During intervals, you should be between 16-20 on Borg scale. | Ca. 40 min |
| Tuesday | Rest | | |
| Wednesday | **Warm-up:** 5-10 minutes increasing your pace and heart rate gradually.<br>**Workout:** 50 minutes over distance training with a moderate pace.<br>**Cool-down**: 5 min with low intensity. | 12-15 on Borg scale. | Ca. 60 min |
| Thursday | Rest | | |
| Friday | **Warm-up:** 5 minutes with low intensity, thereafter 5 minutes with moderate intensity.<br>**Intervals:** 4 x 4 min intervals. 3 min rest between each interval.<br>**Cool-down:** 5 min with low intensity. | During intervals, you should be between 16-20 on Borg scale. | Ca. 40 min |
| Saturday | Rest | | |
| Sunday | Rest | | |

### CONDITION: DURING PREGNANCY

Endurance sessions can be done both outside and indoors. The sessions can be done with any endurance equipment: Bicycle, treadmill, skiers, rowing machine, assault bike or elliptical machine.

| WORKOUT 1 | |
|---|---|
| **What?** | **Duration** |
| **Warm-up:** 5 minutes with low intensity (Borg scale: 7-11)<br><br>**Workout:** 50 min with moderate intensity (Borg scale: 12-15)<br><br>**Cool-down:** 2-3 minutes with low intensity (Borg scale: 7-11) | Ca. 60 min |

| WORKOUT 2 | |
|---|---|
| **What?** | **Duration** |
| **Warm-up**: 5 minutes with low intensity (Borg scale: 7-11), thereafter 5 min med moderat**e** intensity (Borg scale 12-15).<br><br>**Workout:** Every second minute do a 30 second sprint with high intensity (Borg scale 16-17). Repeat until you have completed 5-10 intervals.<br><br>**Cool-down:** 5 minutes with low intensity (Borg scale: 7-11 | Ca. 25-35 min |

### TABATA INTERVALS: DURING PREGNANCY

Tabata is a type of HIIT workout combining high intensity with strength training. Tabata is a four-minute workout consisting of 8 rounds of 20 seconds of work at maximum effort, followed by 10 seconds of rest. Between each station, you have a total of 30 sec of rest. Target heart rate should be between not higher than 85 % of your maximum heart rate (Borg scale: 16-20).

| **TABATA** | | | |
|---|---|---|---|
| **Equipment:** Dumbbells with different load and a yoga mat | | | |
| **Exercise** | **Muscle group** | **Comment** | **Duration** |
| **Station 1:** Air squats<br><br>Modifications:<br>• Wall ball squat<br>• Squat to bench/box | Thighs and gluteal muscle. | Stand with feet shoulder-width apart, toes pointed slightly outward. Engage core muscles and pull shoulder blades together slightly to push out your chest.<br>Squat back as if you were about to sit in a chair. Keep your weight in your heels so you don't lean forward. Your hips should move down and back.<br>Make sure your lower back curve is maintained and keep your heels flat on the floor the whole time. Hips will descend lower than knees. (The eventual goal is to touch your glutes to the backs of your calves.) Hold for a few seconds, then rise up by pushing through heels and using glutes to return to a standing position. | 34 min |
| **Station 2:** Glute Bridge | Thighs, gluteal muscle, stomach and back. | Start flat on your back with your legs bent at a 90-degree angle and feet placed flat on the ground. Make sure your toes are turned outward at 45-degree angles and your knees are facing in the same direction as your toes. Drive down through your feet and push your hips up. You should feel this variation fatiguing the outer portion of your thighs. Make sure you keep your knees over your toes throughout the entire movement. Don't let them move forward over the toes. In a controlled motion, let your hips sink back down toward the ground. This completes 1 repetition. | |
| **Station 3:** Side plank<br><br>Modifications:<br>• Sideplank with bent knees | Stomach and back. | Lie on your right side, legs extended and stacked from hip to feet. The elbow of your right arm is directly under your shoulder. Ensure your head is directly in line with your spine. Your left arm can be aligned along the left side of your body.<br>Engage your abdominal muscles, drawing your navel toward your spine. Lift your hips and knees from the mat while exhaling. Your torso is straight in line with no sagging or bending. Hold the position. 4 times on each side. | |
| **Station 4:** Diagonal lift, standing on all fours | Back, gluteal muscle and thighs. | Stand on all fours with your head extending your body. Flex your abdomen and lower back to stabilise. Alternately stretch one arm and the opposite leg to form extensions of your body. | |
| **Station 5:** Push-ups<br><br>Modifications:<br>• Push-ups against wall/bench<br>• Push-ups on your knees | Chest and arms | Get down on all fours, placing your hands slightly wider than your shoulders. Straighten your arms and legs.<br>Lower your body until your chest nearly touches the floor/wall/bench. Pause, then push yourself back up.<br>Repeat. | |
| **Station 6:** Standing alternating dumbbell curls<br><br>Modifications:<br>• Sitting dumbbell curls | Arms. | Stand with your feet shoulder-width apart and holding a dumbbell in each hand with an overhand grip, with your palms facing your sides. This is your starting position. Raise one dumbbell toward your shoulder while simultaneously rotating the back of your hand. Pause with your palm facing your shoulder. Reverse the movement to lower the weight to the starting position. | |

| | | |
|---|---|---|
| **Station 7:** Dumbbell bent over row on bench | Back. | Choose a flat bench and place a dumbbell on each side of it. Place the right leg on top of the end of the bench, bend your torso forward from the waist until your upper body is parallel to the floor, and place your right hand on the other end of the bench for support. Use the left hand to pick up the dumbbell on the floor and hold the weight while keeping your lower back straight. The palm of the hand should be facing your torso. This will be your starting position. Pull the resistance straight up to the side of your chest, keeping your upper arm close to your side and keeping the torso stationary. Breathe out as you perform this step. Lower the resistance straight down to the starting position. Breathe in as you perform this step. Repeat the movement for the specified number of repetitions. 4 times on each arm. |
| **Station 8:** Dumbbell lateral raise<br><br>Modifications<br>• Sitting. | Shoulders. | Stand or sit with a dumbbell in each hand at your sides. Keep your back straight, brace your core, and then slowly lift the weights out to the side until your arms are parallel with the floor, with the elbow slightly bent. Then lower them back down, again in measured fashion – you'll find it all the harder if you avoid speeding up. A lot of people will cheat by "shrugging" the weights up using their traps. Resist the urge to do that by not raising your shoulder blades during the rep – instead focus on the delts. |

#### EXERCISE PROGRAM: DURING PREGNANCY

What your weekly training plan could look like:

| **EXERCISE PROGRAM** | | | |
|---|---|---|---|
| **Day** | **What?** | **Intensity (Borg scale)** | **Duration** |
| Monday | **Warm-up:** 5 minutes with low intensity, thereafter 5 minutes with moderate intensity.<br>**Intervals:** 4 x 4 min intervals uphill. 3 min rest between each interval. If you choose to do the intervals outside, I recommend finding a long and slight hill. The break will be walking down to the bottom of the hill.<br>**Cool-down**: 5 min with low intensity. | During intervals, you should be between 16-20 on Borg scale. | Ca. 60 min |
| Tuesday | Rest | | |
| Wednesday | **Warm-up:** 5 minutes with low intensity (Borg scale: 7-11), thereafter 5 min med moderate intensity (Borg scale 12-15).<br>**Workout:** Every second minute do a 30 second sprint with high intensity (Borg scale 16-17). Repeat until you have completed 5-10 intervals.<br>**Cool-down:** 5 minutes with low intensity (Borg scale: 7-11 | 16-17 on Borg scale. | Ca. 25-35 min |
| Thursday | Rest | | |
| Friday | **Warm-up:** 5-10 minutes increasing your pace and heart rate gradually.<br>**Workout:** 50 minutes over distance training with a moderate pace.<br>**Cool-down**: 5 min with low intensity. | 12-15 on Borg scale. | Ca. 60 min |
| Saturday | Rest | | |
| Sunday | Rest | | |
