## Supplementary file 2 for "Time-restricted eating and exercise training before and during pregnancy for people with increased risk of gestational diabetes: the BEFORE THE BEGINNING randomised controlled trial"

**Supplementary table 1.** Intention-to-treat analyses of secondary outcomes specific to pregnancy. Data are reported as mean values with standard deviations (SD) of observed values in gestational week 12 and gestational week 28 during the intervention period for *n* participants in each group. Results from linear mixed model analyses are presented as estimated mean difference (Est. effect) in the intervention group compared with the control group, with corresponding 95% confidence interval (CI) and p-values.
*Student's t-test
**Fisher's exact test in gestational week 12 and chi-square test in gestational week 28.

| | | Control (*n* = 54) | Intervention (*n* = 55) | Difference (intervention-control) | | |
|---|---|---|---|---|---|---|
| Outcome | Time | Mean (SD) | Mean (SD) | Est. effect | 95% CI | *p* |
| AUC, OGTT | Gestational week 12 | 796.7 (150.5) | 848.3 (149.1) | | | |
| | Gestational week 28 | 915.1 (132.0) | 977.1 (162.2) | 8.1 | -44.2 to 60.4 | .761 |
| Incremental AUC, OGTT | Gestational week 12 | 195.1 (141.4) | 245.4 (135.1) | | | |
| | Gestational week 28 | 310.5 (124.0) | 364.7 (141.3) | -0.3 | -47.3 to 46.6 | .989 |
| Beta cell function, OGTT $AUC_{insulin}/AUC_{glucose}$ | Gestational week 12 | 6.2 (3.2) | 5.2 (3.0) | | | |
| | Gestational week 28 | 8.2 (3.5) | 6.6 (2.8) | -0.5 | -1.6 to 0.6 | .360 |
| Insulinogenic index during 1st 30 minutes of OGTT | Gestational week 12 | 2.5 (2.1) | 1.9 (2.0) | | | |
| | Gestational week 28 | 2.1 (1.1) | 1.7 (0.8) | 0.2 | -0.6 to 1.0 | .619 |
| Insulin sensitivity $index_{0.120}$ | Gestational week 12 | 84.5 (29.9) | 93.7 (35.8) | | | |
| | Gestational week 28 | 63.6 (13.9) | 65.8 (23.7) | -7.4 | -19.4 to 4.6 | .227 |
| Time to pregnancy*, days | | 83 (69) | 112 (105) | 29.4 | -4.4 to 63.2 | .087 |
| GDM incidence**, *n (%)* | Gestational week 12 | 3 (5.6) | 3 (5.5) | | | 1.000 |
| | Gestational week 28 | 6 (11.1) | 8 (14.5) | | | .776 |

AUC = Area under the curve for glucose levels. OGTT = Oral glucose tolerance test. iAUC = Incremental area under the curve for glucose levels.

**Supplementary Table 2.** Self-reported dietary intake for 3 weekdays and 1 weekend day every 8 weeks. Data are means of observed values with standard deviation (SD) in baseline week, pre-pregnancy period, and 1st, 2nd, and 3rd trimesters of pregnancy for *n* participants in each group. Results from linear mixed model analyses are presented as estimated mean difference (Est. effect) in the intervention group compared with the control group, with corresponding 95% confidence interval (CI) and p-values.

| | | Control (*n* = 83) | Intervention (*n* = 83) | Difference (intervention-control) | | |
|---|---|---|---|---|---|---|
| Outcome | Time | Mean (SD) | Mean (SD) | Est. effect | 95% CI | *p* |
| Total calorie intake, Kcal/day | Baseline | 1792 (396) | 1827 (434) | | | |
| | Pre-pregnancy | 1751 (402) | 1700 (383) | -57.0 | -183.9 to 69.9 | .378 |
| | 1st trimester | 1705 (381) | 1790 (443) | 41.2 | -92.6 to 175.1 | .545 |
| | 2nd trimester | 1856 (375) | 1907 (441) | 39.7 | -101.4 to 180.9 | .580 |
| | 3rd trimester | 2000 (385) | 1914 (487) | -110.3 | -248.0 to 27.3 | .116 |
| Fat, % of total intake | Baseline | 36 (5) | 38 (6) | | | |
| | Pre-pregnancy | 37 (5) | 39 (5) | 0.4 | -1.4 to 2.3 | .646 |
| | 1st trimester | 37 (6) | 37 (5) | 0.1 | -1.9 to 2.1 | .917 |
| | 2nd trimester | 37 (6) | 37 (6) | 0.2 | -1.9 to 2.3 | .878 |
| | 3rd trimester | 36 (6) | 37 (7) | 0.3 | -1.7 to 2.4 | .751 |
| Carbohydrate, % of total intake | Baseline | 46 (6) | 45 (7) | | | |
| | Pre-pregnancy | 45 (5) | 44 (6) | -0.6 | -2.8 to 1.6 | .585 |
| | 1st trimester | 47 (6) | 47 (6) | -0.1 | -2.4 to 2.1 | .908 |
| | 2nd trimester | 47 (6) | 47 (7) | -0.1 | -2.5 to 2.3 | .912 |
| | 3rd trimester | 48 (7) | 47 (8) | -0.7 | -3.1 to 1.6 | .540 |
| Protein, % of total intake | Baseline | 18 (4) | 17 (4) | | | |
| | Pre-pregnancy | 17 (3) | 17 (3) | 0.3 | -0.8 to 1.3 | .616 |
| | 1st trimester | 17 (3) | 17 (3) | 0.1 | -1.0 to 1.1 | .899 |
| | 2nd trimester | 16 (3) | 16 (3) | 0.0 | -1.1 to 1.1 | .977 |
| | 3rd trimester | 16 (3) | 16 (3) | 0.3 | -0.8 to 1.4 | .541 |

**Supplementary Table 3.** Self-reported continuous indicators of weekly physical activity from the International Physical Activity Questionnaire, reported every 8 weeks throughout the study. Data are means of observed values with standard deviation (SD) in baseline week, pre-pregnancy period, and 1st, 2nd, and 3rd trimesters of pregnancy for *n* participants in each group. Results from linear mixed model analyses are presented as estimated mean difference (Est. effect) in the intervention group compared with the control group, with corresponding 95% confidence interval (CI) and p-values.
*95% CI and *p*-values are from bias corrected and accelerated confidence intervals based on bootstrap with 3000 samples, due to non-normally distributed residuals.

| | | Control (*n* = 83) | Intervention (*n* = 83) | Difference (group x time) | | |
|---|---|---|---|---|---|---|
| Outcome | Time | Mean (SD) | Mean (SD) | Est. effect | 95% CI | *p* |
| IPAQ Score, MET minutes/week* | Baseline | 1179 (856) | 1459 (1729) | | | |
| | Pre-pregnancy | 1522 (1215) | 1571 (1474) | 13.5 | -491.9 to 518.8 | .958 |
| | 1st trimester | 916 (1051) | 1161 (1149) | 206.0 | -173.0 to 584.9 | .287 |
| | 2nd trimester | 863 (665) | 1040 (979) | 134.0 | -240.8 to 508.8 | .484 |
| | 3rd trimester | 682 (478) | 798 (521) | 51.7 | -168.7 to 272.1 | .646 |
| Vigorous activity, MET minutes/week* | Baseline | 657 (570) | 694 (856) | | | |
| | Pre-pregnancy | 850 (719) | 775 (443) | -77.2 | -301.1 to 146.7 | .499 |
| | 1st trimester | 577 (465) | 668 (370) | 88.3 | -124.8 to 301.4 | .417 |
| | 2nd trimester | 830 (758) | 683 (2070) | -111.2 | -502.1 to 279.7 | .577 |
| | 3rd trimester | 560 (310) | 751 (406) | 257.8 | -24.6 to 540.1 | .074 |
| Moderate activity, MET minutes/week* | Baseline | 576 (717) | 359 (303) | | | |
| | Pre-pregnancy | 623 (1021) | 442 (380) | -88.8 | -331.7 to 154.1 | .474 |
| | 1st trimester | 470 (499) | 358 (226) | -86.9 | -293.3 to 119.6 | .410 |
| | 2nd trimester | 386 (324) | 294 (243) | -30.5 | -213.9 to 152.8 | .744 |
| | 3rd trimester | 508 (389) | 317 (200) | -124.3 | -317.9 to 69.3 | .208 |
| Walking, MET minutes/week* | Baseline | 527 (424) | 906 (1677) | | | |
| | Pre-pregnancy | 733 (1545) | 903 (1577) | 134.4 | -466.6 to 735.3 | .661 |
| | 1st trimester | 585 (713) | 667 (1054) | 55.8 | -269.1 to 380.7 | .736 |
| | 2nd trimester | 451 (323) | 625 (942) | 153.5 | -196.7 to 503.8 | .390 |
| | 3rd trimester | 508 (390) | 317 (200) | -248.9 | -459.5 to -38.2 | .021 |
| Sitting, minutes/week* | Baseline | 490 (183) | 517 (261) | | | |
| | Pre-pregnancy | 483 (183) | 403 (199) | -81.8 | -143.8 to -19.9 | .010 |
| | 1st trimester | 532 (173) | 472 (148) | -70.8 | -125.2 to -16.3 | .011 |
| | 2nd trimester | 496 (147) | 493 (191) | -40.8 | -124.1 to 42.5 | .337 |
| | 3rd trimester | 508 (389) | 317 (200) | -202.7 | -387.5 to -18.0 | .031 |

IPAQ – International Physical Activity Questionnaire. Metabolic equivalent of task

**Supplementary Table 4.** Time window of energy intake. measured from time of first and last energy intake of the day, reported in the study handbook every 8 weeks throughout the study. Data are means of observed values with standard deviation (SD) in baseline week, pre-pregnancy period, and 1st, 2nd, and 3rd trimesters of pregnancy for *n* participants in each group. Results from linear mixed model analyses are presented as estimated mean difference (Est. effect) in the intervention group compared with the control group, with corresponding 95% confidence interval (CI) and p-values.

| | | Control (*n* = 83) | Intervention (*n* = 83) | Difference (intervention-control) | | |
|---|---|---|---|---|---|---|
| Outcome | Time | Mean (SD) | Mean (SD) | Est. effect | 95% CI | *p* |
| Time window of energy intake, hours/day | Baseline | 12.0 (1.8) | 11.7 (1.7) | | | |
| | Pre-pregnancy | 11.9 (1.4) | 9.9 (1.2) | -2.0 | -2.4 to -1.5 | <.001 |
| | 1st trimester | 12.0 (1.5) | 10.4 (1.3) | -1.4 | -1.9 to -0.8 | <.001 |
| | 2nd trimester | 12.2 (1.4) | 10.7 (1.4) | -1.3 | -1.9 to -0.7 | <.001 |
| | 3rd trimester | 12.4 (1.2) | 10.5 (1.5) | -1.5 | -2.1 to -1.0 | <.001 |
| Time of first energy intake, hours | Baseline | 8.9 (1.2) | 9.0 (1.4) | | | |
| | Pre-pregnancy | 9.0 (1.1) | 9.4 (1.0) | 0.4 | -0.2 to 0.9 | .882 |
| | 1st trimester | 8.9 (1.1) | 8.9 (1.0) | 0.0 | -0.6 to 0.6 | .553 |
| | 2nd trimester | 8.8 (0.9) | 8.8 (0.9) | 0.0 | -0.6 to 0.7 | .668 |
| | 3rd trimester | 8.9 (1.0) | 9.1 (0.9) | 0.2 | -0.4 to 0.8 | .808 |
| Time of last energy intake, hours | Baseline | 20.8 (1.3) | 20.7 (1.3) | | | |
| | Pre-pregnancy | 20.9 (1.0) | 19.2 (1.2) | -1.8 | -2.1 to -1.4 | <.001 |
| | 1st trimester | 20.9 (1.0) | 19.3 (0.9) | -1.4 | -1.7 to -1.0 | <.001 |
| | 2nd trimester | 21.1 (1.1) | 19.6 (1.3) | -0.3 | -0.7 to 0.1 | .170 |
| | 3rd trimester | 21.3 (1.1) | 19.6 (1.2) | -1.4 | -1.8 to -1.0 | <.001 |

**Supplementary Table 5.** Personal activity intelligence (PAI) points and percentage (%) of time spent at high. moderate and low intensity recorded by smartwatches throughout the study in the intervention group. Data are means of observed values with standard deviations (SD) in baseline week, pre-pregnancy period, and 1st, 2nd and 3rd trimesters of pregnancy for *n* participants. Within-group differences were estimated using linear mixed models and reported as estimated mean difference (Est. effect) compared with baseline with corresponding 95% confidence intervals (CI) and p-values.
*95% CI and *p*-values are from bias corrected and accelerated confidence intervals based on bootstrap with 3000 samples, due to non-normally distributed residuals.

| | | Intervention (n = 83) | Within group difference | | |
|---|---|---|---|---|---|
| Outcome | Time | Mean (SD) | Est. effect | 95% CI | *p* |
| PAI points/week | Baseline | 76 (62) | | | |
| | Pre-pregnancy | 111 (54) | 36.3 | 23.0 to 47.8 | <.001 |
| | 1st trimester | 85 (43) | 3.3 | -11.8 to 17.5 | .703 |
| | 2nd trimester | 71 (39) | -16.8 | -33.2 to -1.2 | .035 |
| | 3rd trimester | 77 (55) | -13.0 | -31.4 to 4.5 | .143 |
| Time spent at high intensity, %* | Baseline | 3 (7) | | | |
| | Pre-pregnancy | 4 (6) | 1.7 | 0.1 to 3.3 | .037 |
| | 1st trimester | 3 (3) | 0.4 | -1.1 to 1.9 | .600 |
| | 2nd trimester | 2 (2) | -1.1 | -2.9 to 0.6 | .209 |
| | 3rd trimester | 1 (1) | -1.8 | -3.6 to 0.0 | .055 |
| Time spent at moderate intensity, % | Baseline | 16 (17) | | | |
| | Pre-pregnancy | 15 (12) | -1.9 | -4.8 to 1.1 | .212 |
| | 1st trimester | 14 (11) | -2.6 | -6.0 to 0.7 | .124 |
| | 2nd trimester | 14 (9) | -3.6 | -7.3 to 0.1 | .058 |
| | 3rd trimester | 12 (11) | -5.1 | -9.3 to -1.0 | .016 |
| Time spent at low intensity, % | Baseline | 82 (20) | | | |
| | Pre-pregnancy | 81 (16) | 0.2 | -3.2 to 3.6 | .901 |
| | 1st trimester | 83 (13) | 2.1 | -1.9 to 6.0 | .308 |
| | 2nd trimester | 84 (11) | 5.0 | 0.7 to 9.4 | .023 |
| | 3rd trimester | 88 (12) | 7.0 | 2.2 to 11.9 | .004 |
