## Supplementary file 3 for "Time-restricted eating and exercise training before and during pregnancy for people with increased risk of gestational diabetes: the BEFORE THE BEGINNING randomised controlled trial"

**Supplementary Table 1 - Per protocol analysis.** Data are observed means and standard deviations (SD) at baseline, in pre-pregnancy week 8, gestational week 12, and gestational week 28 for *n* participants in each group. Results from linear mixed model analyses are presented as estimated mean difference (Est. effect) in the intervention group compared with the control group, with corresponding 95% confidence interval (CI) and *p*-values.
*95% CI and *p*-values are from bias corrected and accelerated confidence intervals based on bootstrap with 3000 samples, due to non-normally distributed residuals.

| | | Control ($n$ = 54) | Intervention ($n$ = 31) | Difference (group x time) | | |
|---|---|---|---|---|---|---|
| Outcome | Time | Mean (SD) | Mean (SD) | Est. effect | 95% CI | *p* |
| ***Primary outcome*** | | | | | | |
| 2-hour blood glucose after OGTT, mmol/L | Gestational week 12 | 5.8 (1.4) | 5.3 (1.1) | | | |
| | Gestational week 28 | 6.4 (1.1) | 6.5 (1.5) | 0.64 | 0.0 to 1.3 | .049 |
| ***Secondary outcomes*** | | | | | | |
| | | Control ($n$ = 83) | Intervention ($n$ = 31) | Difference (group x time) | | |
| Outcome | Time | Mean (SD) | Mean (SD) | Est. effect | 95% CI | *p* |
| Fasting blood glucose, mmol/L | Baseline | 5.0 (0.4) | 4.9 (0.4) | | | |
| | Pre-pregnancy week 8 | 5.1 (0.4) | 4.9 (0.3) | -0.1 | -0.2 to 0.0 | .094 |
| | Gestational week 12 | 4.5 (0.3) | 4.4 (0.3) | -0.1 | -0.2 to 0.1 | .431 |
| | Gestational week 28 | 4.5 (0.4) | 4.5 (0.6) | 0.0 | -0.1 to 0.2 | .722 |
| HbA1c, mmol/mol | Baseline | 34.3 (3.1) | 33.9 (3.0) | | | |
| | Pre-pregnancy week 8 | 34.5 (3.2) | 33.7 (2.2) | -0.6 | -1.4 to 0.3 | .191 |
| | Gestational week 12 | 32.7 (2.3) | 33.0 (1.9) | 0.5 | -0.5 to 1.4 | .306 |
| | Gestational week 28 | 32.3 (2.8) | 31.7 (3.1) | -0.3 | -1.2 to 0.6 | .536 |
| Fasting insulin, µIU/mL* | Baseline | 19.1 (11.7) | 14.9 (9.9) | | | |
| | Pre-pregnancy week 8 | 19.3 (9.7) | 14.4 (8.1) | -2.6 | -5.7 to 0.5 | .104 |
| | Gestational week 12 | 12.8 (10.7) | 13.6 (8.7) | 2.5 | -2.7 to 7.6 | .353 |
| | Gestational week 28 | 19.4 (12.2) | 19.9 (11.5) | 1.7 | -4.2 to 7.5 | .578 |
| HOMA2-B* | Baseline | 167.8 (58.9) | 151.5 (72.1) | | | |
| | Pre-pregnancy week 8 | 171.2 (59.2) | 148.1 (54.5) | -15.6 | -36.2 to 4.9 | .136 |
| | Gestational week 12 | 153.8 (81.4) | 171.1 (75.2) | 25.0 | -16.2 to 66.1 | .235 |
| | Gestational week 28 | 212.4 (84.9) | 220.8 (73.5) | 11.5 | -25.4 to 48.5 | .541 |
| HOMA2-IR* | Baseline | 2.4 (1.4) | 1.9 (1.1) | | | |
| | Pre-pregnancy week 8 | 2.4 (1.2) | 1.8 (1.0) | -0.3 | -0.7 to 0.1 | .091 |
| | Gestational week 12 | 1.6 (1.2) | 1.7 (1.0) | 0.3 | -0.3 to 1.0 | .265 |
| | Gestational week 28 | 2.3 (1.4) | 2.4 (1.4) | -2.6 | -0.4 to 1.0 | .462 |
| Total Cholesterol, mmol/L | Baseline | 4.6 (0.7) | 4.7 (0.9) | | | |
| | Pre-pregnancy week 8 | 4.5 (0.7) | 4.8 (0.9) | 0.2 | 0.0 to 0.5 | .092 |
| | Gestational week 12 | 4.4 (0.7) | 4.4 (0.7) | -0.1 | -0.4 to 0.2 | .606 |
| | Gestational week 28 | 5.9 (1.2) | 5.8 (0.8) | -0.2 | -0.5 to 0.1 | .288 |
| LDL cholesterol, mmol/L | Baseline | 3.0 (0.9) | 1.5 (0.4) | | | |
| | Pre-pregnancy week 8 | 2.9 (0.8) | 1.5 (0.3) | 0.2 | 0.0 to 0.5 | .296 |
| | Gestational week 12 | 2.8 (0.7) | 1.6 (0.3) | -0.1 | -0.4 to 0.2 | .559 |

| | | | | | | |
|---|---|---|---|---|---|---|
| | Gestational week 28 | 3.9 (1.2) | 1.8 (0.4) | -0.2 | -0.5 to 0.1 | .173 |
| HDL cholesterol, mmol/L | Baseline | 1.4 (0.3) | 3.1 (1.0) | | | |
| | Pre-pregnancy week 8 | 1.4 (0.3) | 3.1 (0.9) | 0.0 | -0.1 to 0.2 | .418 |
| | Gestational week 12 | 1.6 (0.4) | 2.7 (0.8) | 0.0 | -0.2 to 0.1 | .547 |
| | Gestational week 28 | 1.8 (0.4) | 3.8 (0.7) | 0.0 | -0.1 to 0.1 | .965 |
| Triglycerides, mmol/L | Baseline | 1.0 (0.5) | 0.9 (0.4) | | | |
| | Pre-pregnancy week 8 | 1.0 (0.5) | 0.9 (0.3) | 0.0 | -0.2 to 0.1 | .690 |
| | Gestational week 12 | 1.1 (0.4) | 1.0 (0.3) | 0.0 | -0.2 to 0.2 | .837 |
| | Gestational week 28 | 2.0 (0.7) | 1.8 (0.5) | -0.1 | -0.3 to 0.1 | .151 |
| Weight, kg | Baseline | 81.5 (13.2) | 80.0 (16.0) | | | |
| | Pre-pregnancy week 8 | 81.6 (12.5) | 78.8 (15.6) | -1.0 | -2.4 to 0.5 | .186 |
| | Gestational week 12 | 79.5 (12.9) | 78.2 (15.4) | -2.1 | -3.7 to -0.5 | .012 |
| | Gestational week 28 | 86.8 (12.3) | 85.7 (15.7) | -2.5 | -4.1 to -0.8 | .003 |
| Fat mass, kg | Baseline | 31.6 (9.8) | 29.3 (10.9) | | | |
| | Pre-pregnancy week 8 | 31.2 (9.5) | 28.0 (10.4) | -1.1 | -2.3 to 0.1 | .068 |
| | Gestational week 12 | 30.6 (9.5) | 28.9 (10.6) | -1.3 | -2.6 to 0.1 | .072 |
| | Gestational week 28 | 34.2 (9.0) | 32.7 (11.0) | -1.9 | -3.2 to -0.5 | .007 |
| Fat percentage, % | Baseline | 37.9 (7.2) | 35.7 (7.6) | | | |
| | Pre-pregnancy week 8 | 37.6 (7.0) | 34.6 (7.3) | -1.0 | -1.9 to 0.0 | .048 |
| | Gestational week 12 | 37.7 (7.0) | 36.2 (6.8) | -0.7 | -1.8 to 0.4 | .188 |
| | Gestational week 28 | 38.8 (6.0) | 37.4 (7.2) | -1.0 | -2.1 to 0.0 | .054 |
| Visceral fat area, $cm^2$ | Baseline | 154 (52) | 138 (55) | | | |
| | Pre-pregnancy week 8 | 151 (50) | 130 (54) | -6.3 | -13.5 to 1.0 | .091 |
| | Gestational week 12 | 148 (51) | 132 (52) | -6.9 | -15.2 to 1.4 | .101 |
| | Gestational week 28 | 165 (47) | 152 (54) | -8.8 | -16.9 to -0.7 | .034 |
| Muscle mass, kg | Baseline | 27.7 (3.6) | 28.2 (4.5) | | | |
| | Pre-pregnancy week 8 | 28.0 (3.6) | 28.2 (4.4) | 0.1 | -0.3 to 0.4 | .777 |
| | Gestational week 12 | 27.0 (3.2) | 27.2 (4.2) | -0.3 | -0.7 to 0.2 | .239 |
| | Gestational week 28 | 29.1 (3.1) | 29.4 (4.4) | -0.3 | -0.8 to 0.1 | .136 |
| Waist circumference, cm | Baseline | 94.5 (11.8) | 92.9 (12.7) | | | |
| | Pre-pregnancy week 8 | 93.9 (11.2) | 90.4 (10.7) | -2.1 | -5.0 to 0.8 | .157 |
| | Gestational week 12 | 95.1 (11.5) | 93.6 (12.8) | -1.7 | -4.9 to 1.5 | .302 |
| | Gestational week 28 | 107.3 (8.4) | 104.7 (10.4) | -3.5 | -6.8 to -0.3 | .033 |
| Systolic blood pressure, mmHg | Baseline | 121.0 (9.9) | 120.1 (8.3) | | | |
| | Pre-pregnancy week 8 | 119 (8) | 117.5 (9.2) | -0.8 | -4.0 to 2.4 | .638 |
| | Gestational week 12 | 114 (10) | 110.3 (8.1) | -3.2 | -6.8 to 0.4 | .078 |
| | Gestational week 28 | 110 (9) | 109.1 (9.6) | -1.4 | -5.0 to 2.3 | .469 |
| Diastolic blood pressure, mmHg | Baseline | 79 (7) | 80.1 (6.3) | | | |
| | Pre-pregnancy week 8 | 77 (6) | 77.0 (5.7) | -0.8 | -3.4 to 1.7 | .519 |
| | Gestational week 12 | 71 (8) | 70.4 (6.7) | -1.5 | -4.4 to 1.3 | .29 |
| | Gestational week 28 | 69 (89) | 69.3 (9.4) | -0.7 | -3.7 to 2.3 | .643 |
| Resting heart rate, beats per minute | Baseline | 72 (11) | 67.6 (8.2) | | | |
| | Pre-pregnancy week 8 | 69 (9) | 64.8 (8.9) | -2.6 | -6.8 to 1.5 | .212 |
| | Gestational week 12 | 71 (11) | 69.1 (17.3) | 0.0 | -4.6 to 4.7 | .985 |
| | Gestational week 28 | 78 (12) | 75.9 (15.7) | -0.8 | -5.6 to 4.0 | .738 |

OGTT = Oral glucose tolerance test, HbA1c = Glycated haemoglobin, HOMA2-B = Homeostatic model assessment of beta cell function, HOMA2-S = Homeostatic model assessment of insulin sensitivity, HOMA2-IR = Homeostatic model assessment of insulin resistance, HDL = High-density lipoprotein cholesterol, LDL = Low-density lipoprotein cholesterol.

**Supplementary Table 2 - Per protocol analysis:** Self-reported dietary intake for 3 weekdays and 1 weekend day every 8 weeks. Data are means of observed values with standard deviation (SD) in baseline week, pre-pregnancy period, and 1$^{st}$, 2$^{nd}$, and 3$^{rd}$ trimesters of pregnancy for *n* participants in each group. Results from linear mixed model analyses are presented as estimated mean difference (Est. effect) in the intervention group compared with the control group, with corresponding 95% confidence interval (CI) and p-values.

| | | Control (*n* = 83) | Intervention (*n* = 31) | Difference (intervention - control) | | |
|---|---|---|---|---|---|---|
| Outcome | Time | Mean (SD) | Mean (SD) | Est. effect | 95% CI | *p* |
| Total calorie intake, Kcal/day | Baseline | 1792 (396) | 1838 (466) | | | |
| | Pre-pregnancy | 1751 (402) | 1751 (407) | -14.3 | -168.6 to 139.9 | .855 |
| | 1st trimester | 1705 (381) | 1811 (450) | 83.6 | -83.7 to 251.0 | .326 |
| | 2nd trimester | 1856 (375) | 1845 (469) | 1.9 | -179.3 to 183.1 | .983 |
| | 3rd trimester | 2000 (385) | 1926 (502) | -103.1 | -278.1 to 71.9 | .247 |
| Fat, % of total intake | Baseline | 36 (5) | 38 (5) | | | |
| | Pre-pregnancy | 37 (5) | 38 (5) | 0.4 | -1.9 to 2.6 | .756 |
| | 1st trimester | 37 (6) | 36 (6) | -0.9 | -3.3 to 1.5 | .460 |
| | 2nd trimester | 37 (6) | 37 (7) | 0.4 | -2.3 to 3.0 | .788 |
| | 3rd trimester | 36 (6) | 37 (8)) | 0.5 | -2.0 to 3.0 | .694 |
| Carbohydrate, % of total intake | Baseline | 46 (6) | 45 (6) | | | |
| | Pre-pregnancy | 45 (5) | 45 (7) | -0.1 | -2.7 to 2.4 | .920 |
| | 1st trimester | 47 (6) | 48 (7) | 1.0 | -1.8 to 3.7 | .489 |
| | 2nd trimester | 47 (6) | 47 (8) | -0.2 | -3.2 to 2.8 | .890 |
| | 3rd trimester | 48 (7) | 47 (10) | -0.9 | -3.8 to 2.0 | .542 |
| Protein, % of total intake | Baseline | 18 (4) | 17 (4) | | | |
| | Pre-pregnancy | 17 (3) | 17 (3) | -0.1 | -1.4 to 1.1 | .840 |
| | 1st trimester | 17 (3) | 17 (3) | -0.1 | -1.4 to 1.3 | .909 |
| | 2nd trimester | 16 (3) | 16 (2) | -0.2 | -1.7 to 1.2 | .763 |
| | 3rd trimester | 16 (3) | 16 (3) | 0.3 | -1.1 to 1.7 | .720 |

**Supplementary Table 3 – Per protocol analysis:** Self-reported continuous indicators of weekly physical activity from the International Physical Activity Questionnaire, reported every 8 weeks throughout the study. Data are means of observed values with standard deviation (SD) in baseline week, pre-pregnancy period, and 1st, 2nd, and 3rd trimesters of pregnancy for *n* participants in each group. Results from linear mixed model analyses are presented as estimated mean difference (Est. effect) in the intervention group compared with the control group, with corresponding 95% confidence interval (CI) and p-values.
*95% CI and *p*-values are from bias corrected and accelerated confidence intervals based on bootstrap with 3000 samples, due to non-normally distributed residuals.

| | | Control (*n* = 83) | Intervention (*n* = 31) | Difference (intervention - control) | | |
|---|---|---|---|---|---|---|
| Outcome | Time | Mean (SD) | Mean (SD) | Est. effect | 95% CI | *p* |
| IPAQ Score, MET minutes/week* | Baseline | 1179 (856) | 1715 (1736) | | | |
| | Pre-pregnancy | 1522 (1215) | 1547 (1077) | -69.3 | -558.4 to 419.9 | .781 |
| | 1st trimester | 916 (1051) | 1091 (663) | 100.8 | -255.3 to 456.8 | .579 |
| | 2nd trimester | 863 (665) | 1093 (750) | 96.8 | -364.1 to 557.7 | .681 |
| | 3rd trimester | 682 (478) | 768 (522) | -41.2 | -289.2 to 206.8 | .745 |
| Vigorous activity, MET minutes/week* | Baseline | 657 (570) | 844 (1085) | | | |
| | Pre-pregnancy | 850 (719) | 856 (374) | -20.8 | -267.7 to 226.1 | .869 |
| | 1st trimester | 577 (465) | 666 (319) | 66.2 | -162.9 to 295.3 | .571 |
| | 2nd trimester | 830 (758) | 833 (225) | 15.2 | -401.0 to 431.5 | .943 |
| | 3rd trimester | 560 (310) | 673 (412) | 195.3 | -172.6 to 563.1 | .298 |
| Moderate activity, MET minutes/week* | Baseline | 576 (717) | 349 (316) | | | |
| | Pre-pregnancy | 623 (1021) | 422 (379) | -78.2 | -350.9 to 194.5 | .574 |
| | 1st trimester | 470 (499) | 415 (252) | -0.4 | -248.2 to 247.5 | .998 |
| | 2nd trimester | 386 (324) | 332 (282) | 20.8 | -231.0 to 272.7 | .871 |
| | 3rd trimester | 508 (389) | 297 (189) | -97.3 | -328.2 to 133.7 | .409 |
| Walking, MET minutes/week* | Baseline | 527 (424) | 1064 (1402) | | | |
| | Pre-pregnancy | 733 (1545) | 977 (1347) | 199.2 | -481.1 to 879.4 | .566 |
| | 1st trimester | 585 (713) | 515 (333) | -96.1 | -346.4 to 154.1 | .452 |
| | 2nd trimester | 451 (323) | 450 (357) | -15.1 | -232.8 to 202.5 | .892 |
| | 3rd trimester | 508 (390) | 297 (189) | -258.1 | -474.9 to -41.4 | .020 |
| Sitting, minutes/week* | Baseline | 490 (183) | 470 (261) | | | |
| | Pre-pregnancy | 483 (183) | 378 (198) | -99.3 | -558.4 to 419.9 | .781 |
| | 1st trimester | 532 (173) | 446 (136) | -84.4 | -255.3 to 456.8 | .579 |
| | 2nd trimester | 496 (147) | 483 (222) | -45.8 | -364.1 to 557.7 | .681 |
| | 3rd trimester | 508 (389) | 297 (189) | -212.7 | -289.2 to 206.8 | .745 |

IPAQ – International Physical Activity Questionnaire, Metabolic equivalent of task

**Supplementary table 4 - Per protocol analysis of secondary outcomes specific to pregnancy:** Data are reported as mean values with standard deviations (SD) of observed values in gestational week 12 and gestational week 28 during the intervention period for *n* participants in each group. Results from linear mixed model analyses are presented as estimated mean difference (Est. effect) in the intervention group compared with the control group, with corresponding 95% confidence interval (CI) and p-values.
*Student's t-test
**Fisher's exact test in gestational week 12 and chi-square test in gestational week 28.

| | | Control (*n* = 54) | Intervention (*n* = 24) | Difference (intervention - control) | | |
|---|---|---|---|---|---|---|
| Outcome | Time | Mean (SD) | Mean (SD) | Est.effect | 95% CI | p |
| AUC, OGTT | Gestational week 12 | 796.7 (150.5) | 815.3 (130.6) | | | |
| | Gestational week 28 | 915.1 (132.0) | 959.3 (173.7) | 26.4 | -40.7 to 93.4 | .441 |
| Incremental AUC, OGTT | Gestational week 12 | 186.8 (150.5) | 219.9 (126.3) | | | |
| | Gestational week 28 | 310.5 (124.0) | 349.1 (140.7) | 12.1 | -47.9 to 72.0 | .689 |
| Beta cell function, OGTT AUCinsulin/AUCglucose | Gestational week 12 | 6.2 (3.2) | 5.3 (3.2) | | | |
| | Gestational week 28 | 8.1 (3.5) | 6.8 (2.9) | -0.2 | -1.7 to 1.2 | .751 |
| Insulinogenic index during 1st 30 minutes of OGTT | Gestational week 12 | 2.5 (2.1) | 1.6 (0.8) | | | |
| | Gestational week 28 | 2.0 (1.2) | 1.8 (0.8) | 0.6 | -0.2 to 1.4 | .118 |
| Insulin sensitivity index 0,120 | Gestational week 12 | 87.4 (36.3) | 98.0 (33.4) | | | |
| | Gestational week 28 | 67.7 (12.6) | 68.1 (24.3) | -9.4 | -23.1 to 4.3 | .180 |
| Time to pregnancy*, days | | 83 (69) | 132 (65) | 50 | 16 to 82 | .004 |
| GDM incidence**, *n (%)* | Gestational week 12 | 3 (5.6 %) | 0 (0 %) | | | .549 |
| | Gestational week 28 | 6 (11.1 %) | 3 (14.5 %) | | | 1.000 |

AUC = Area under the curve for glucose levels, OGTT = Oral glucose tolerance test, iAUC = Incremental area under the curve for glucose levels, GDM = Gestational diabetes
