## Supplementary file 4 for "Time-restricted eating and exercise training before and during pregnancy for people with increased risk of gestational diabetes: the BEFORE THE BEGINNING randomised controlled trial"

### PRECONCEPTION LIFESTYLE INTERVENTIONS TO IMPROVE FUTURE METABOLIC HEALTH

**Protocol Identification:** Before the Beginning

| | |
|---|---|
| **SPONSOR:** | **Øivind Rognmo, Head of Department**<br>Address: Department of Circulation and Medical Imaging<br>Norwegian University of Science and Technology<br>Postbox 8905<br>7491 Trondheim, Norway<br>Phone: +47 92613734<br>E-Mail: |
| **PRINCIPAL INVESTIGATOR (PI):** | **Trine Moholdt, Research Scientist**<br>Address: Department of Circulation and Medical Imaging, NTNU DMF<br>Phone: +47 97098594<br> |

**PROTOCOL VERSION NO. 8.0 – 03.10.2023**

### CONTACT DETAILS

**Sponsor:** **Øivind Rognmo, Head of Department**

Address: Department of Circulation and Medical Imaging

Norwegian University of Science and Technology

Postbox 8905

7491 Trondheim, Norway

**Principal investigator:** **Trine Moholdt**

Address: Department of Circulation and Medical Imaging, NTNU DMF

### SIGNATURE PAGE

| | |
|---|---|
| Title | BEFORE THE BEGINNING:<br>Preconception lifestyle interventions to improve future metabolic health |
| Protocol ID no: | Before the Beginning |

***I hereby declare that I will conduct the study in compliance with the Protocol, ICH GCP and the applicable regulatory requirements:***

| Name | Title | Role | Signature | Date |
|---|---|---|---|---|
| Trine Moholdt | Research Scientist | Principal Investigator | | |
| Guro Rosvold | Technician | Investigator | | |
| Kjell Å. Salvesen | Professor | Investigator | | |
| Siri Ann Nyrnes | Post Doc fellow | Investigator | | |
| Ann-Charlotte Iversen | Professor | Investigator | | |
| Elisabeth Eide Axe | Nurse | Investigator | | |
| Md Abu Jafar Sujan | PhD Student | Investigator | | |
| Hilde Normann Lund | Research Assistant | Investigator | | |
| Hanna Skarstad | Medical student | Investigator | | |

| | | |
|---|---|---|
| Kamilla Haganes | PhD candidate | Investigator |
| Stine Fougner | Research Scientist | Investigator |
| Svein-Erik Måsøy | Researcher | Investigator |
| Anders Emil Vrålstad | PhD candidate | Investigator |
| Karin Deibele | PhD candidate | Investigator |
| Mariana Flores Gutierrez | Master student | Investigator |
| Adeyori Peter Oladewa | Master student | Investigator |
| Wajjiha Amir | Master student | Investigator |
| Valentina Ivanovic | PhD student | Investigator |
| Ole Jakob Mengshoel | Professor | Investigator |

### PROTOCOL SYNOPSIS

BEFORE THE BEGINNING: Preconception lifestyle interventions to improve future metabolic health

| | |
|---|---|
| Sponsor | Øivind Rognmo, Head of Department, Department of Circulation and Medical Imaging, NTNU |
| Phase and study type | Interventional RCT study |
| Centers: | Single center (Trondheim, NTNU) |
| Study Period: | Estimated date of first participant enrolled:<br>01.09. 2020 in Trondheim<br>Anticipated recruitment period:<br>2020 to 2023 in Trondheim<br>Estimated date of last participant completed:<br>01.06.2024 in Trondheim |
| Treatment Duration: | Expected treatment duration pr. participant:<br>Preconception: 0-12 months (0-6 months from December 2022)<br>Pregnancy: 9 months<br><br>Expected follow-up period pr. participant:<br>Post delivery: 6 weeks.<br>Consents for potential long-term follow-up of offspring. |

| | |
|---|---|
| Objectives | Main study objective: To determine if a lifestyle intervention, initiated preconception and continued throughout pregnancy, will improve maternal glycaemic control in pregnancy and thereby offspring health.<br><br>Exploratory: Extensive data collection from mother-infant pairs will allow for exploratory investigations of underlying mechanisms for the influence of maternal health on offspring health. |
| Endpoints: | Primary endpoint: Plasma glucose concentration (after 2 h oral glucose tolerance testing) in gestational week 28.<br><br>Secondary endpoints: Insulin sensitivity, circulating lipids, cytokines, HbA1c, continuous glucose regulation, body composition, cardiorespiratory fitness, blood pressure, and offspring body composition and cardiac function. |
| Study Design: | Randomized controlled trial with an intervention group and a control group. |
| Main Inclusion Criteria: | • Women who are contemplating pregnancy within the next six months<br>• Age: 18-39 years old<br>• At least one of the following criteria: BMI ≥ 25 < 40 kg/m$^2$, gestational diabetes in a previous pregnancy, close relatives with diabetes (either parents, siblings or children with diabetes) previous infant > 4.5 kg, fasting plasma glucose > 5.3 mmol/L, or Non-European ethnicity |

| | |
|---|---|
| Main Exclusion Criteria | • On-going pregnancy<br>• Trying to conceive ≥ 6 cycles at study entry<br>• Known diabetes (type 1 or 2)<br>• Shift work that includes night shifts > 2 days per week<br>• Previous hyperemesis<br>• Cardiovascular diseases<br>• High intensity exercise ≥ 2x/week the last 3 months<br>• Habitual eating window ≤ 12 hours<br>• Bariatric surgery<br>• Any other reason which according to the researchers makes the potential participant ineligible |
| Sample Size: | Minimum 200 participants, or the number of participants needed to reach minimum 45 pregnant participants in each group in gestational week 12. |

### TABLE OF CONTENTS

**CONTACT DETAILS** ... **1**
**SIGNATURE PAGE** ... **2**
**PROTOCOL SYNOPSIS** ... **4**
**TABLE OF CONTENTS** ... **7**
**1 INTRODUCTION** ... **10**
1.1 Background ... 10
**2 STUDY OBJECTIVES AND RELATED ENDPOINTS** ... **13**
2.1 Primary and Secondary Endpoint ... 13
**3 OVERALL STUDY DESIGN** ... **15**
**4 STUDY POPULATION** ... **16**
4.1 Selection of Study Population ... 16
4.2 Number of Participants ... 16
4.3 Inclusion Criteria ... 16
4.4 Exclusion Criteria ... 16
**5 TREATMENT** ... **17**
5.1 Intervention ... 17
5.2 Duration of Therapy ... 18
5.3 User involvement ... 18
5.4 Schedule Modifications ... 18
5.5 Concomitant Medication ... 18
**6 STUDY PROCEDURES** ... **19**
6.1 Flow Chart ... 19
6.2 By Visit ... 20
6.2.1 Preconception ... 20
6.2.2 Pregnancy ... 20
6.2.3 After End of Treatment (Follow-up) ... 20
6.2.4 Withdrawal ... 20
6.3 Criteria for Participant Discontinuation ... 21
6.4 Procedures for Discontinuation ... 21
6.4.1 Participant Discontinuation ... 21
6.4.2 Trial Discontinuation ... 21
**7 ASSESSMENTS** ... **22**
7.1 Assessment of Efficacy ... 22
7.2 Assessments of Compliance ... 24
7.3 Predictions ... 24
**8 SAFETY MONITORING AND REPORTING** ... **24**

### 1 INTRODUCTION

#### 1.1 Background

The global prevalence of gestational diabetes, i.e., high plasma glucose first identified during pregnancy, continues to increase. Both environmental factors and genetics contribute to the development of gestational diabetes, with up to 14% of live births negatively impacted by this condition.[1] Obesity and insulin resistance not only predispose the mother to later type 2 diabetes and cardiovascular disease, but also affect egg cell quality and the intrauterine environment, thus programming the child for cardiometabolic diseases.[2] Gestational diabetes is independently associated with impaired glucose tolerance in childhood and exposure to hyperglycaemia in utero is strongly related with childhood obesity.[2] Even glucose concentrations lower than those diagnostic of gestational diabetes are associated with increased birth weight and elevated levels of cord-blood C-peptide, childhood obesity, and elevated blood pressure, independent of maternal body mass index (BMI).[2] Exercise-diet interventions to prevent adverse pregnancy outcomes typically start in the second trimester and have proven to be challenging, with low adherence and limited efficacy.[3] Although genes from the mother partly explain inherited risk, recent data demonstrate that epigenetic modifications (i.e. heritable changes in phenotype that do not involve alterations of the genetic code itself) play a major role in the effects of maternal health on the next generation (**Fig. 1**).[4]

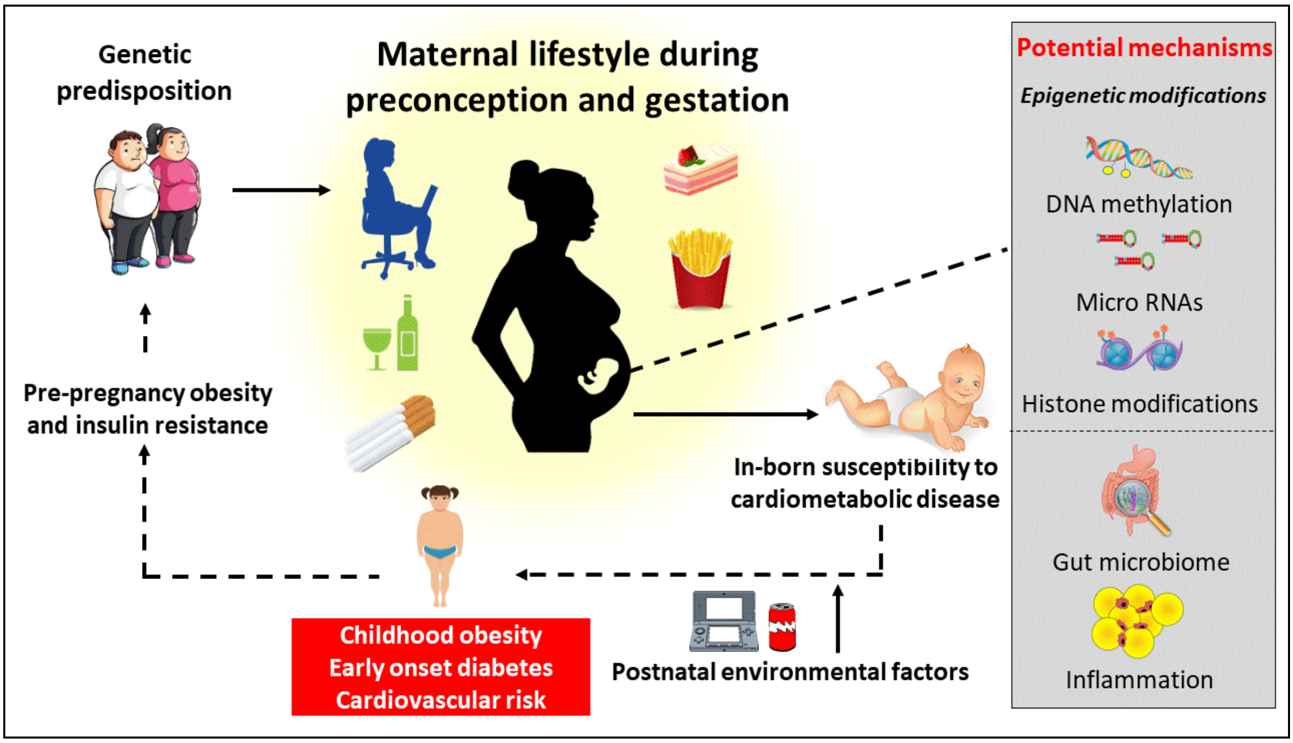

**Fig. 1. The intergenerational cycle of chronic cardio-metabolic disorders.** Poor preconception and gestational maternal lifestyle predispose both the mother and the baby to unfavourable pregnancy outcomes, creating an intergenerational cycle of obesity, insulin resistance, and associated disorders (*from Moholdt & Hawley, 2020*).

Several other possible mechanisms underpin the adverse cardiometabolic consequences for the child of maternal hyperglycaemia and/or obesity, including pre-pregnancy insulin resistance with accompanying hyperinsulinemia and low-grade inflammation, as well as modifications of the gut microbiome.[2]

Several recent randomised controlled trials (RCTs) and reviews on lifestyle interventions in pregnancy conclude that pre-pregnancy interventions are urgently needed to improve pregnancy outcomes for mother and child.[5-7] In our ETIP (Exercise Training in Pregnancy) trial[8] including 91 women with a pre-pregnancy BMI ≥28 kg/m$^2$, we reported no effect of exercise on gestational weight gain but significantly reduced incidence of gestational diabetes in the exercise group (6%) compared to the control group (27%).[9] We also found reduced levels of circulating insulin postpartum among the women in the exercise group compared to the control group.[10] Despite the improved glycaemic control, 35% of women in the exercise group and 52% in the control group delivered a baby > 4 kg.[11] Our exercise program included 140 min of moderate intensity endurance training per week, but only 50% of the participants adhered to this program, similar to other exercise interventions in pregnancy. After discussions with the participants in ETIP, we discovered that the main reasons for such low adherence were that the participants did not enjoy the exercise program and had difficulties scheduling time to exercise. Practical, enjoyable approaches and motivational support are clearly needed to improve adherence to exercise in pregnancy. Since pre-pregnancy patterns of physical activity and exercise is an important determinant of exercise during pregnancy[12], commencing exercising before pregnancy is necessary.

A preconception diet and physical activity intervention has only been reported by one previous study: the Finnish Gestational Diabetes Prevention Study (RADIEL), which involved a sub-group of women at high risk of gestational diabetes.[13] The intervention included an initial group visit with a dietician and visits to study nurses every 3 months preconception but showed no effect on gestational diabetes incidence. Probably more intense, yet practical, programs are required, with closer follow-up and extensive motivational support.

We discovered impaired cardiac function and thicker interventricular septum in the newborns of women in the ETIP trial, compared to newborns of normal weight women[7], with reduced cardiac function already present in gestational week 14.[14] The reduced cardiac function was sustained also when the babies were 6-8 weeks old Maternal insulin resistance in the very early stages of pregnancy alters placental physiology, which will influence the supply of nutrients to the fetus, and thereby organ development.[15] No prior study have determined if a maternal lifestyle change, initiated before pregnancy and continued throughout gestation, can improve the intrauterine environment during this crucial time for fetal heart development. Since completion of the ETIP trial, there has been major advancements in cardiovascular ultrasound imaging at our department and we are now able to also measure the stiffness of the heart in the babies. This is highly relevant in this population since offspring of diet-induced obese mice[16] and sheep[17] have increased myocardial fibrosis, but no human data exist.

**High intensity interval training (HIT)**, defined as short periods of intense activity separated by low-intensity breaks, induces superior improvements in insulin sensitivity and fitness compared to continuous moderate intensity training in subjects at increased risk for cardiometabolic diseases.[18] Even short-term (6 weeks) interventions with brief (15-60 sec) work-bouts and a total time commitment of <45 min per week, improves insulin sensitivity similar to that attained after 6 months of traditional endurance training.[19] HIT also enhances exercise enjoyment and adherence compared to continuous, prolonged moderate intensity training.[20] At present, there is limited data on the effect of HIT on glycaemic control in pregnancy, probably due to concerns about fetal well-being. However, during brief, intense exercise bouts the maternal heart rate does not exceed 90% of maximum and is within the safety zone for fetal wellbeing.[21] We have pilot data showing normal fetal heart rates after repeated 30 sec maximum effort exercise bouts in pregnant women. Furthermore, results from our group show that HIT for 10 weeks improves insulin sensitivity by ~20% in overweight women of reproductive age.[22,23] In an on-going trial of HIT in women with polycystic ovary syndrome, we see a 56% reduction in the prevalence of impaired glucose tolerance after 16 weeks. Of note, eight women allocated to HIT, versus none in the control group, got pregnant during the intervention period or the following 36 weeks of unsupervised exercise. **HIT is therefore a highly potent intervention that elicits important changes in a range of clinically relevant health outcomes in reproductive-aged females.** In summary, there is a scientific rationale for HIT to optimise exercise prescription before and during pregnancy without risk of adverse effects for the mother or baby.

**Time-restricted eating (TRE)** is a novel eating regimen whereby the duration of fasting between the last evening meal and the first meal of the next day is prolonged. Such an eating pattern reduces obesity, inflammation and insulin resistance.[24] One 'proof-of-concept' study reported significant improvements in insulin sensitivity after only 5 weeks of TRE in men with increased risk of diabetes.[24] The participants reported lower appetite in the evening and had fully adjusted to the new eating schedule within two weeks of initiation.[24] In BEFORE THE BEGINNING, women are therefore able to adapt to the new eating regimen prior to pregnancy. No previous study has determined the effect of TRE on glycaemic control in pregnancy. We are currently undertaking a pilot study on TRE during pregnancy and our initial data indicate that this eating regimen is feasible in pregnancy; our participants can restrict the time-window of energy intake to 9.4 ± 0.8 h/day (unpublished).

**Our primary hypothesis** is that the combination of TRE and HIT, commencing pre-conception and continuing throughout pregnancy, will substantially improve maternal glucose tolerance in pregnancy. We also hypothesise that the maternal intervention will prevent cardiac dysfunction in the newborn, will reduce the number of offspring with birth weight > 4 kg, and promote anti-inflammatory responses.

### 2 STUDY OBJECTIVES AND RELATED ENDPOINTS

Our main objective is to establish an optimal diet-exercise prevention strategy that commences *before* conception aiming at reducing the incidence of gestational diabetes (GDM). This conceptual shift in maternal care provides a platform for improved health outcomes for mother and offspring, breaking the intergenerational cycle of cardiometabolic disorders, therefore reducing the risk of diabetes in the future generations.

#### 2.1 Primary and Secondary Endpoint

| | Objectives | Endpoints | Assessments |
|---|---|---|---|
| Primary | Glycaemic control | Plasma glucose concentration | 2h oral glucose tolerance test (75 g glucose) |
| Secondary Maternal | Insulin sensitivity | Fasting circulating insulin and glucose | HOMA-IR |
| | Glycaemic control | Area under the curve for insulin and glucose | 2h oral glucose tolerance test (75 g glucose) |
| | Body composition | Fat mass<br>Fat-free mass<br>Visceral fat<br>Body weight | InBody720 bioimpedance scale |
| | | Waist circumference | Measuring tape |
| | Fasting plasma, serum and full blood* | Lipids, cytokines, HbA1c | Blood sampling. Some material will be frozen for later analyses (not yet determined) |
| | Continuous glucose monitoring | Glucose | FreeStyle Libre, worn for 14 days |
| | Cardiorespiratory fitness | Maximum oxygen uptake | Metalyzer |
| | Blood pressure | Systolic and diastolic | Automatic cuff |

| | | | |
|---|---|---|---|
| | Physical activity | Physical activity level | Sensewear and smart watches |
| | Nutrition | Diet diary | Calorie counter by FatSecretapp |
| | Health and lifestyle | Score | Questionnaire |
| | Urine Sampling* | Metabolic | Metabolomics |
| Secondary Offspring | Fetal ultrasound | Fetal growth and cardiac measurements | Standard fetal exam including adaptive imaging |
| | Standard clinical neonatal outcomes | Birth weight, head circumference, Apgar score, days at the hospital, admission to ICU. | Hospital birth records |
| | Epigenetic modifications* | Umbilical cord blood | Cardiac function markers, insulin, glucose, DNA methylation and RNA sequencing. |
| | Body composition in newborn | Body fat, muscle mass, bone density, hydration status | Bioimpedance (BioScan tough i8-nano). |
| | Blood pressure and cardiac function | Systolic and diastolic | Automatic cuff |
| | Cardiac function | Blood flow and myocardial function, including tissue stiffness | Standard echocardiography, including mechanical wave imaging (MWI).<br>Vivid E95 scanner (GE Vingmed Ultrasound, Norway), and a GE 12s, 6s and/or M5s phased-array transducer |

Exploratory*
In addition to the specified outcome measures, we will collect bio samples for future analyses including maternal and cord-blood and urine samples.

**Table 1:** Overview of assessments in the study.

### 3 OVERALL STUDY DESIGN

The study is an RCT with two parallel groups (Fig.2): intervention group (HIT + TRE) and standard care control group. After screening, and initial assessments, women will be randomly allocated (1:1) after stratifying gestational diabetes in a previous pregnancy (yes/no) to the intervention or a standard care control group.

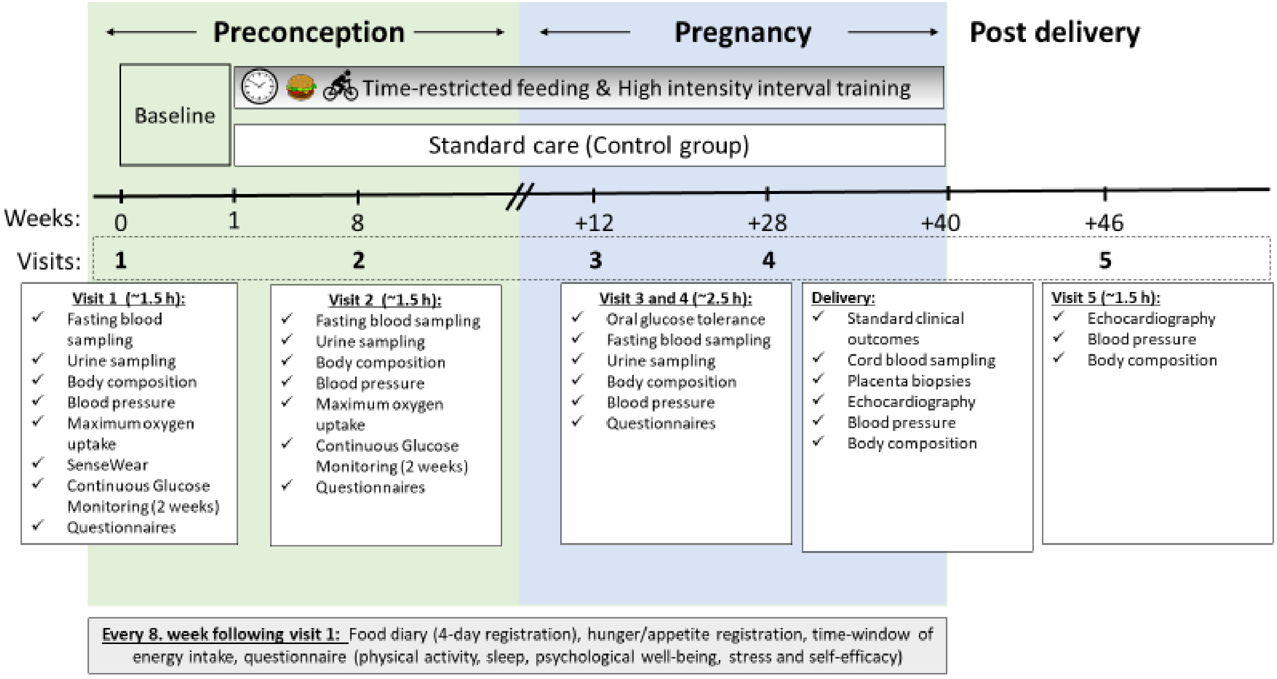

**Fig.2 Schematic overview of the trial.** Women will visit the lab twice during preconception (Visit 1 and 2) and twice during pregnancy (Visit 3 in gestational week 12 and Visit 4 in gestational week 28) and 6 weeks postpartum (Visit 5). Additional data will be collected by questionnaires, physical activity tracking and diet diaries at baseline and every 8 weeks following visit 1. Comprehensive bio-sampling, metabolic and cardiovascular measurements at each visit, as described in subsequent text. The outcomes at delivery and Visit 5 will be measured in the new-borns.

| | |
|---|---|
| Study Period | Estimated date of first participant enrolled: 01.09.2020 |
| | Anticipated recruitment period: 2020 to 2022 |
| | Estimated date of last participant completed: 01.06.2024 |
| Treatment Duration: | Expected treatment duration pr. participant: |
| | Preconception: 0-6 months |
| | Pregnancy: 9 months |
| | Expected follow-up period pr. participant: |
| | Post-delivery: 6 weeks |

Consent for potential long-term follow-up of offspring.

### 4 STUDY POPULATION

#### 4.1 Selection of Study Population

**Description of the study settings:**
The study centre is the Norwegian University of Science and Technology (NTNU). Measurements will be undertaken in an academic hospital (St. Olavs hospital) in Norway. Subjects will be recruited by mailed invitations and through posts on social media. Additional participants will be recruited through local health clinics, where ~ 95% of Norwegian children come for vaccination. The recruitment letter will prompt women to visit the study website, which will contain a short description of the trial and allows women to self-screen for eligibility before further screening by telephone.

#### 4.2 Number of Participants

We will include minimum 200 participants in this study, or the number needed to reach at least 45 pregnant participants in each group in gestational week 12.

#### 4.3 Inclusion Criteria

All the following conditions must apply to the prospective participant at screening prior to study participation:

- Female
- Age: 18-39 years old
- Contemplating pregnancy within the next six months
- Understands oral and written Norwegian or English
- At least one of the following criteria must apply: BMI ≥ 25 < 40 $kg/m^2$, GDM in a previous pregnancy, close relative with diabetes (either parents, siblings or children with diabetes) previous infant > 4.5 kg, fasting plasma glucose > 5.3 mmol/L, or Non-European ethnicity (with one or both parents originating from an area outside Europe).

#### 4.4 Exclusion Criteria

Participants will be ineligible for participation in the study if they meet any of the following criteria at baseline:

- On-going pregnancy
- Trying to conceive ≥ 6 cycles at study entry
- Known diabetes (type 1 or 2)

- Shift work that includes night shifts > 2 days per week
- Previous hyperemesis
- Known cardiovascular diseases
- High intensity exercise > 2x/week the last 3 months
- Habitual eating window ≤ 12 hours
- Bariatric surgery
- Any other reason which according to the researchers makes the potential participant ineligible

### 5 TREATMENT

#### 5.1 Intervention

Participants will be counselled by a registered dietician to change their “time window” of food intake to an 8-10 h period of their choice but ending no later than 19:00 for five days per week throughout the intervention. The remaining two days will be “days off” where they can consume food *ad libitum* if they wish (some might want to keep to TRE for all 7 days). Depending on participants’ preferences, the days off will typically be weekend days or days they choose to exercise in the evening. Apart from current recommendations about preconception/pregnancy nutrition, no advice regarding food choices/macronutrient composition will be given, nor any attempt to encourage a reduced total energy intake. We will use Personal Activity Intelligence (PAI), a science-backed activity metric based on heart rate (https://www.ntnu.edu/cerg/personal-activity-intelligence) to prescribe exercise. The goal for the participants in the intervention group will be to earn 100 PAI per rolling 7 days, which can be reached with one hour of exercise if the intensity is ≥ 80% of the heart rate maximum. We will motivate and encourage the participants to do so by providing a “menu” of exercise options to reach this goal. It will also be arranged a more structured training program, for those who want that. The mode of exercise (e.g., treadmill walking/running, cycling) during pre-pregnancy will be individualised based on their preferences. This program will continue until pregnancy (for up to 12 months, or up to 6 months from December 2022). For participants who experience spontaneous abortions, we will add the number of weeks that the participant has been pregnant to the 6-12 months’ time-window. Once pregnant, we will advise the participants to either do short intervals or longer intervals with lower intensity (up to 85% of heart rate maximum). In addition, we will use “friendly competition” among participants as a tool to increase adherence. Participants who may be concerned about the safety of their foetus during HIT, can exercise at the hospital with monitoring of the foetal heart rate during training.

Adherence is an essential aspect of the study: encouragement, individual programming and ongoing support is vital to ensure maximal adherence (i.e., >80 % of all scheduled training sessions). Motivational support will be provided face-to-face on one weekly exercise session, as well as through an electronic application providing notifications, guidance and feedback on the exercise sessions. Participants will also attend researcher-led telephone consultations every other week to provide encouragement, support and monitoring to improve adherence to the TRE protocol. On the trial website, an element of (anonymous) “friendly competition” between participants will be added, where they can log their weekly exercise and adherence to the TRE. Combining motivational human interaction with digital interventions increases engagement and the effectiveness of behaviour change interventions.[25] Adherence will be closely monitored on a weekly basis for all participants, through recordings of PAI-data (using an online dashboard) and through self-reported weekly summary reports of TRE adherence, delivered through the trial website.

The control group will receive standard care and will not be discouraged from being physically active or healthy eating. All participants will receive a brochure from the Norwegian Health Directorate with the current recommendations for physical activity, diet, folic acid and iodine supplements at inclusion. The participants will be invited to an additional ultrasound examination in week 12 and 32 if they become pregnant.

#### 5.2 Duration of Therapy

Women will visit the lab twice during preconception; baseline assessments (visit 1) and after 8 weeks (visit 2). Also, twice during pregnancy; gestational week 12 (visit 3) and gestational week 28 (visit 4). Last visit will be 6 weeks postpartum (Visit 5). We changed the maximum time before pregnancy from 12 to 6 months in December 2022 to allow for the trial to terminate in time for us to analyse the data within the project period.

#### 5.3 User involvement

We have involved users in the planning of the study and will involve users in the implementation and dissemination. In the planning phase, we have arranged a one-hour digital workshop with the users, where we encouraged the audience to ask questions and give us feedback about relevant topics or issues related to research participation. A highly relevant topic when it comes to user involvement is the long-term adherence to lifestyle changes. This will be discussed with the users. Feedback from the participants regarding their challenges to incorporate exercise training and TRE into their daily life will be very valuable for our further research and for the implementation of exercise programs in clinical practice.

#### 5.4 Schedule Modifications

If the participants are not able to complete either exercising or TRE, we will focus on the other component of the intervention.

#### 5.5 Concomitant Medication

We will ask the participants to report any medications, changes in medications and changes in their diet and/or training regime.

### 6 STUDY PROCEDURES

#### 6.1 Flow Chart

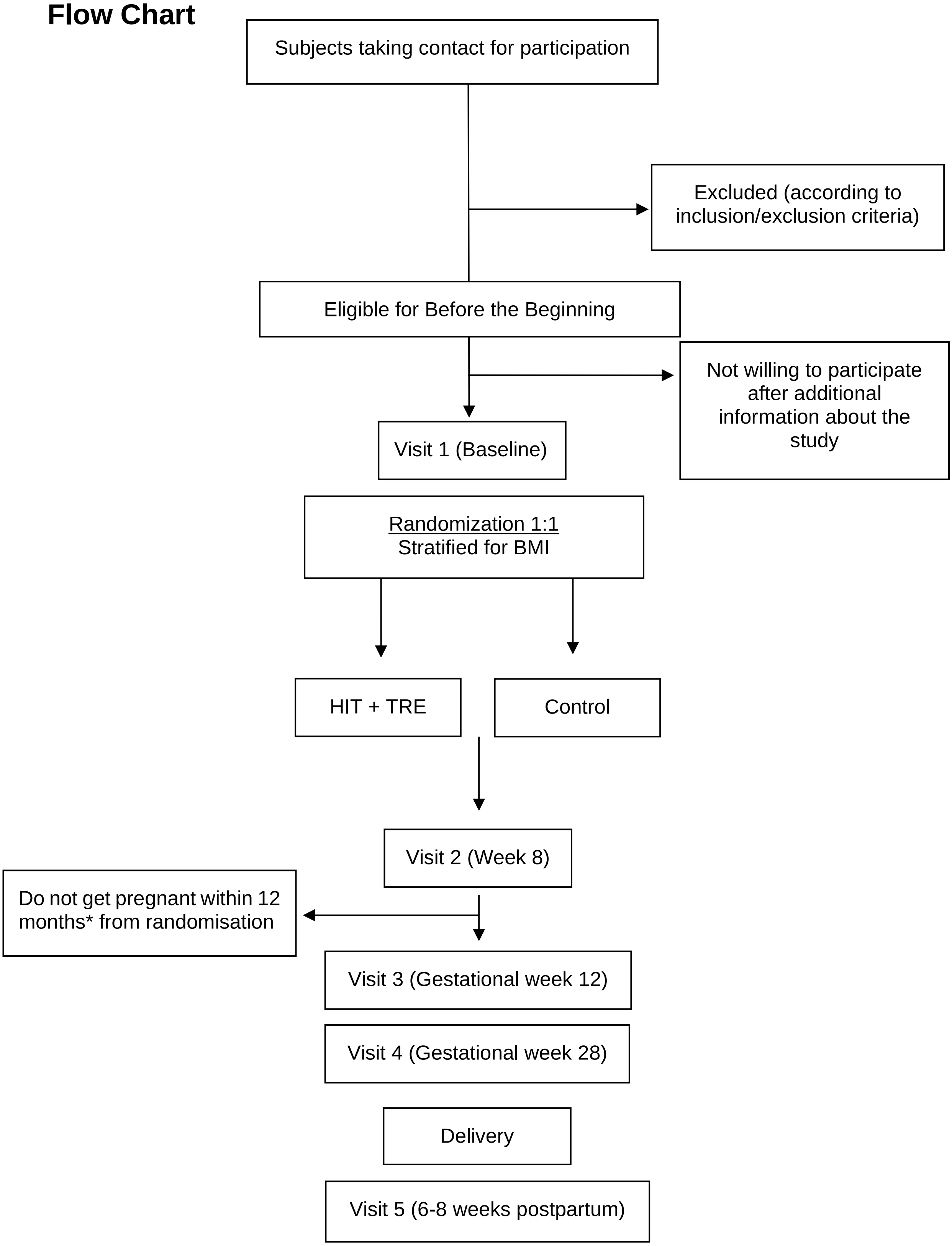

**Fig. 3.** Participant flow throughout Before the Beginning – study. * From December 2022, the time-window for exclusion if not pregnant was reduced from 12 to 6 months.

#### 6.2 By Visit

**Informed consent**

Each subject must have given informed consent voluntarily before any study specific procedures are initiated.

**Clinical status**

| | |
|---|---|
| Questionnaires: | Physical activity, psychological well-being, diet, background information, hunger and appetite, time-window for daily energy intake. |
| Physical examination: | Height and weight, body composition, waist circumference, blood pressure, blood samples and oral glucose tolerance test, insulin sensitivity, continuous glucose monitoring, physical activity tracking, cardiorespiratory fitness testing.<br><br>Standard clinical neonatal outcome, umbilical cord blood, subcutaneous adipose tissue thickness, blood pressure and cardiac function. |

**Laboratory analysis**

##### 6.2.1 Preconception

Fasting blood sampling, urine sampling, body composition, waist circumference, blood pressure, maximum oxygen uptake, questionnaires, continuous glucose monitoring and physical activity monitoring.

##### 6.2.2 Pregnancy

Oral glucose tolerance, urine sampling, fasting blood sampling, body composition, blood pressure, questionnaires, continuous glucose monitoring and physical activity monitoring.

##### 6.2.3 After End of Treatment (Follow-up)

Standard clinical outcomes at delivery, cord blood sampling, echocardiography, blood pressure and body composition.

##### 6.2.4 Withdrawal

The participants can at any time withdraw from the study without further explanation. However, we will include participants that withdraw from the study in our analysis (intention to treat) with all available data on the participants.

#### 6.3 Criteria for Participant Discontinuation

Participants may be discontinued from study treatment and assessments at any time. Specific reasons for discontinuing a participant for this study are:

- Voluntary discontinuation by the participant who is at any time free to discontinue her participation in the study, without prejudice to further treatment.
- Safety reason as judged by the Principal Investigator
- Major protocol deviation
- Incorrect enrolment i.e., the participant does not meet the required inclusion/exclusion criteria for the study
- Participant lost to follow-up
- Participants who do not get pregnant within 12 months from randomisation (within 6 months from December 2022).

#### 6.4 Procedures for Discontinuation

##### 6.4.1 Participant Discontinuation

Participants who withdraw or are withdrawn from the study, will stop further intervention. However, we will include participants that withdraw or are withdrawn from the study in our analysis (intention to treat). This includes data from participants who are withdrawn due to not falling pregnant within 12 months from randomisation (within 6 months from December 2022).

If possible, a final assessment will be made (end of study intervention and follow-up). The reason for discontinuation will be recorded. The investigator will follow up any significant adverse events until the outcome is either recovered or resolved.
Participants who withdraw or are withdrawn from the study before start of intervention, will be replaced.

##### 6.4.2 Trial Discontinuation

The whole trial may be discontinued at the discretion of the Principal Investigator or the Sponsor in the event of any of the following:

- Occurrence of Adverse Events unknown to date in respect of their nature, severity, and duration
- Medical or ethical reasons affecting the continued performance of the trial
- Difficulties in the recruitment of participants.

The Sponsor and Principal Investigator will inform all investigators, the relevant Competent Authorities and Ethics Committees of the termination of the trial along with the reasons for such action. If the study is terminated early on grounds of safety, the Competent Authorities and Ethics Committees will be informed within 30 days.

### 7 ASSESSMENTS

#### 7.1 Assessment of Efficacy

We assess the participants twice at preconception, twice during pregnancy and 6 weeks postpartum. All methods are established in our laboratories.

The following outcomes will be recorded:

**The primary outcome measure** is plasma glucose concentration and will be obtained after a 2 h oral glucose tolerance test (OGGT, 75 g glucose) in gestation week 28, the standard time-point for GDM diagnosis.

**Secondary maternal outcome measures that will be obtained at Visit 1-4, if not otherwise described:** Insulin sensitivity will be calculated using homeostasis model assessment of insulin resistance (HOMA-IR)[26] and pancreatic beta cell function using HOMA-β.[26] At visit 3 and 4, area under the curve (AUC) and incremental AUC (iAUC) glucose and insulin concentrations will be calculated from venous blood sampling every 30 min during the 120 min oral glucose tolerance testing. Insulin Sensitivity Index, $ISI_{0,120}$.[27], insulinogenic index during the first 30 min of the oral glucose tolerance testing[28] and beta cell function ($AUC_{ins}/AUC_{glu}$) will be estimated.[29] From all visits, additional fasting plasma, serum, full blood and urine will be stored in a biobank at -80°C for later analyses. Blood lipids, glucose and HbA1c will be measured immediately after sampling, at St. Olavs Hopital, following local procedures.

Continuous glucose monitoring will be undertaken for 7 days pre-intervention followed by the first 7 days of intervention/control, and in weeks 6-8 from randomisation, using Freestyle Libre (Abbott Diabetes Care).

GDM will be recorded at Visit 3 and 4, according to the WHO 2013 criteria (fasting plasma glucose 5.1-6.9 mmol/L and/or 2 h plasma glucose 8.5-11.0 mmol/L after 75 g OGGT).[30]

Body composition will be estimated using bioelectrical impedance. To account for the increase in fat-free mass hydration as pregnancy progresses, a regression equation that estimates fat-free mass density as a function of gestational age will be used.[31]

Waist circumference will be measured using a measuring tape at the level of the belly button.

Cardiorespiratory fitness (maximum oxygen uptake, $VO_2$max) will be measured by maximum effort treadmill tests using direct analyses of expired gas at visit 1 and 2.

Blood pressure will be measured in the seated position after 15 minutes of rest with an automatic blood pressure device (Welch Allyn, Germany) three times with one-minute intervals, in left arm (diastolic and systolic, in mmHg).

Physical activity will be recorded using the activity monitors Sensewear and Amazefit GTS. Sensewear will be worn for 14 days at baseline (Visit 1), while Amazefit will be worn throughout the intervention for participants in the intervention group. Physical activity will additionally be self-reported (using the International Physical Activity Questionnaire) every 8 weeks (Appendix A).

Diet will be registered in an online food diary for four days (three weekdays and one weekend day) every 8 weeks throughout the study period. Participants will be sent text messages as reminders about dietary reporting to ensure compliance.

Questionnaires: Participants will complete detailed questionnaires about their health and lifestyle (Appendix B-E) at each visit. Time to pregnancy, early miscarriages, abortions, time to live birth, medication and supplements taken will also be recorded. Psychological well-being will be measured with a Psychological general well-being index (PGWBI) questionnaire (Appendix B). Sleep will be measured using The Horne-Östberg Morningsness-Eveningness Questionnaire (Appendix F) and Pittsburgh Sleep Quality Index (PSQI) (Appendix G). Expectant fathers will be asked to complete questionnaires at baseline and every 8 weeks throughout the trial including questions regarding weight, height, physical activity and diet that will be used as co-variates in later analyses (Appendix H and I).

Fetal ultrasound: Week 12 (visit 3), 19 and 32 with focus on fetal growth and cardiac exam. We will save raw data in addition to standard images to validate a new adaptive technique to optimize image quality. Equipment: CE approved GE Expert Voluson 22 scanner with accompanying probes. The examination in week 12 and 19 will be combined with standard screening ultrasound.

Standard clinical neonatal outcomes will be obtained from the hospital birth records.

Umbilical cord blood will be collected immediately after birth, prior to the delivery of the placenta. Samples will be immediately refrigerated and transported to -80°C within 24 h for later analyses.

Placental tissues are collected from 1) around the base of the umbilical cord on the foetal side, 2) the periphery on the maternal side (full-thickness tissue) 3) the centre of the maternal side, also for storage in RNAlater solution (Invitrogen, Thermofisher scientific, Lithuania) and 4% formaldehyde solution, and 4) the periphery on the maternal side. The samples are put in 1.8 mL cryotubes and snap-frozen immediately in liquid nitrogen, before storage at -80°C for later analyses. The samples in RNAlater solution are stored at 4°C overnight, followed by storage at -80°C for later analyses. The samples in 4% formaldehyde solution are stored under a fume hood at room temperature for 48 hours before histology slide preparation in collaboration with the CMIC Histology Lab at NTNU.

Body composition of the new-born will be measured using bioimpedance (BioScan touch i8-nano, Maltron, UK) 72 h of birth, and after 6-8 weeks.

Cardiac morphology, structure and function of the new-born will be measured by an experienced paediatric cardiologist using a Vivid E95 scanner (GE Vingmed Ultrasound, Norway) and a GE 12s, 6s and/or M5s phased-array transducer within 72 h of birth, and after 6-8 weeks. A full clinical echocardiography including conventional echocardiographic parameters as well as study images with a focus on measurement of systolic and diastolic myocardial function is performed. A corresponding group of neonates (N = 30), from mothers with no known increased risk of GDM and BMI in the normal range (18.6-24.9 kg/m2) will be used for comparison.

#### 7.2 Assessments of Compliance

We will measure compliance to the intervention by questionnaires on time-window for energy intake and using the PAI registrations of physical activity level.

#### 7.3 Predictions

We will use data from the pre-pregnancy period and early pregnancy to assess if we can find early predictors for GDM. In these analyses we will develop artificial intelligence (AI) methods with the goal of improving the understanding and early identification of GDM risk.

### 8 SAFETY MONITORING AND REPORTING

The investigator is responsible for the detection and documentation of events meeting the criteria and definition of an adverse event (AE) or serious adverse event (SAE). Each participant will be instructed to contact the investigator immediately if she manifests with any signs or symptoms they perceive as serious.

The methods for collection of safety data are described below.

#### 8.1 Definitions

##### 8.1.1 Adverse Event (AE)

An AE is any untoward medical occurrence in a participant administered a pharmaceutical product and which does not necessarily have a causal relationship with this treatment.

An adverse event (AE) can therefore be any unfavourable and unintended sign (including an abnormal laboratory finding), symptom, or disease temporally associated with the use of a medicinal (investigational) product, whether or not related to the medicinal (investigational) product.

The term AE is used to include both serious and non-serious AEs.

If an abnormal laboratory value/vital sign are associated with clinical signs and symptoms, the sign/symptom will be reported as an AE and the associated laboratory result/vital sign should be considered additional information that must be collected on the relevant CRF.

##### 8.1.2 Serious Adverse Event (SAE)

Any untoward medical occurrence that:

- Results in death
- Is immediately life-threatening
- Requires in-participant hospitalization or prolongation of existing hospitalization
- Results in persistent or significant disability or incapacity
- Is an important medical event that may jeopardize the subject or may require medical intervention to prevent one of the outcomes listed above

Medical and scientific judgment is to be exercised in deciding on the seriousness of a case. Important medical events may not be immediately life threatening or result in death or hospitalization but may jeopardize the subject or may require intervention to prevent one of the listed outcomes in the definitions above. In such situations, or in doubtful cases, the case should be considered as serious. Hospitalization for administrative reason (for observation or social reasons) is allowed at the investigator’s discretion and will not qualify as serious unless there is an associated adverse event warranting hospitalization.

##### 8.1.3 Suspected Unexpected Serious Adverse Reaction (SUSAR)

Adverse Reaction: all untoward and unintended responses to an investigational medicinal product related to any dose administered.

Unexpected Adverse Reaction: an adverse reaction, the nature or severity of which is not consistent with the applicable product information.

Suspected Unexpected Serious Adverse Reaction: SAE (see section 8.1.2) that is unexpected as defined in section 8.2 and possibly related to the investigational medicinal product(s).

#### 8.2 Expected Adverse Events

We expect no adverse events. However, if it occurs any adverse events, it will be recorded in the CRF.

#### 8.3 Time Period for Reporting AE and SAE

For each participant the standard time period for collecting and recording AE and SAEs will begin at signature of informed consent and will continue until the end of the follow-up period.

During the course of the study all AEs and SAEs will be proactively followed up for each participant; events should be followed up to resolution, unless the event is considered by the investigator to be unlikely to resolve due to the underlying disease. Every effort should be made to obtain a resolution for all events, even if the events continue after discontinuation/study completion.

#### 8.4 Recording of Adverse Events

If the participant has experienced adverse event(s), the investigator will record the following information in the CRF:

- The nature of the event(s) will be described by the investigator in precise standard medical terminology
- The duration of the event will be described in terms of event onset date and event ended data.
- The Causal relationship of the event to the study intervention will be assessed as one of the following:

**Unrelated:**

There is not a temporal relationship to investigational intervention, or there is a reasonable causal relationship between non-investigational intervention, concurrent disease, or circumstance and the AE.

**Unlikely:**

There is a temporal relationship to investigational intervention, but there is not a reasonable causal relationship between the investigational intervention and the AE.

**Possible:**

There is reasonable causal relationship between the investigational intervention and the AE. Dechallenge information is lacking or unclear.

**Probable:**

There is a reasonable causal relationship between the investigational intervention and the AE. The event responds to dechallenge. Rechallenge is not required.

**Definite:**

There is a reasonable causal relationship between the investigational intervention and the AE.

- Action taken
- The outcome of the adverse event – whether the event is resolved or still ongoing.

It is important to distinguish between serious and severe AEs. Severity is a measure of intensity whereas seriousness is defined by the criteria in Section 8.1. An AE of severe intensity is not necessarily considered as serious.

#### 8.5 Reporting Procedure

##### 8.5.1 AEs and SAEs

All adverse events and serious adverse events that should be reported as defined in section 8.1.1 will be recorded in the participants CRF. SAEs must be reported by the investigator to the sponsor, Liaison Committee between the Central Norway Regional Health Authority (RHA) and the Norwegian University of Science and Technology (NTNU), within 24 hours after the site has gained knowledge of the SAE. Every SAE must be documented by the investigator on the SAE pages (to be found in ISF or CRF). The Serious Adverse Event Report Form must be completed and signed. The initial report shall promptly be followed by detailed, written reports if necessary. The initial and follow-up reports shall identify the trial subjects by unique code numbers assigned to the latter.

The sponsor keeps detailed records of all SAEs reported by the investigators and performs an evaluation with respect to seriousness, causality and expectedness.

##### 8.5.2 SUSARs

SUSARs will be reported to the Competent Authority and Ethics Committee according to national regulation. The following timelines should be followed:

The sponsor will ensure that all relevant information about suspected serious unexpected adverse reactions that are fatal or life-threatening is recorded and reported as soon as possible to the Competent Authority and Ethics Committee in any case no later than seven (7) days after knowledge by the sponsor of such a case, and that relevant follow-up information is subsequently communicated within an additional eight (8) days.

All other suspected serious unexpected adverse reactions will be reported to the Competent Authority concerned and to the Ethics Committee concerned as soon as possible but within a maximum of fifteen (15) days of first knowledge by the sponsor.

##### 8.5.3 Clinical Study Report

The adverse events and serious adverse events occurring during the study will be discussed in the safety evaluation part of the Clinical Study Report.

#### 8.6 Procedures in Case of Emergency

The investigator is responsible for assuring that there are procedures and expertise available to cope with emergencies during the study.

### 9 DATA MANAGEMENT AND MONITORING

#### 9.1 Case Report Forms (CRFs)

The designated investigator staff will enter the data required by the protocol into the Case report forms (eCRF). The Principal Investigator is responsible for assuring that data entered into the CRF is complete, accurate, and that entry is performed in a timely manner. The signature of the investigator will attest the accuracy of the data on each CRF. If any assessments are omitted, the reason for such omissions will be noted on the eCRFs. Corrections, with the reason for the corrections will also be recorded. Paper copies of analyses, tests, questionnaires, and logs will be stored (without participants' names, only with study ID numbers).

After database lock, the investigator will receive a CD-ROM or paper copies of the subject data for archiving at the investigational site.

#### 9.2 Confidentiality

The investigator shall arrange for the secure retention of the participant identification and the code list. Participant files shall be kept for the maximum period permitted by each hospital. The study documentation (CRFs, Site File etc.) shall be retained and stored during the study and for 15 years after study closure. All information concerning the study will be stored in a safe place inaccessible to unauthorized personnel.

#### 9.3 Database management

A unique subject number that is assigned when subject signs the Informed Consent Form identifies each subject in the study. Once assigned the subject number cannot be reused for any other subject. The same primary identifier will be used throughout the study. A code connects the participant to the data through a name list, stored in a locker, during and after the trial. Only authorized personnel have access to this list and can find back to the participant.

9.3.1 Use of ultrasound data for other projects: The ultrasound-data collected in this project (raw unprocessed ultrasound data in addition to standard images) are generic and may be used to perform research, test and validate many new algorithms for improving ultrasound images. This is a core research and development interest for the partners in this project. In such, the data collected have very high value for both research, clinical and commercialization purposes, and the partners are planning to use the data for several future projects of image enhancement, also including machine learning algorithms. The benefit of this for patients, is that a limited recruitment of patients at single time instance, yields data that can potentially lead to improved methods many years from now

#### 9.4 Determination of Sample Size and Statistical methods

Power calculation

The primary outcome of this study is glucose tolerance (after a 2 hr oral glucose tolerance testing) in gestational week 28. We consider a difference between the intervention group and the control group of 1.0 mmol/L as the minimally clinical relevant difference, based on findings from the HAPO study[32], and have used the observed standard deviation (SD) in 2-hr plasma glucose from HAPO in our calculations. Sample size computed for a two-sided t-test to determine difference between groups 1.0 mmol/L difference between groups, a SD of 1.3, a statistical power of 0.90 and confidence level of 0.05, yields 37 participants in each group. To allow for an expected exclusion from the study due to not conceiving within the study period (~50%)[33], yielding 74 per group, further drop-out during the study period (10-20%), yielding 93 per group, and to increase statistical power for secondary analyses, we initially estimated that we would need to include 260 participants in the trial. We will, however, terminate inclusion of new participants when we have reached minimum 45 participants in each group who are pregnant in gestational week 12 (to allow for 20% dropout during pregnancy), or at least 200 participants in total (accounting for 20% dropout before pregnancy and the observed 58.5% pregnancy rate in the ongoing trial).

Statistical analyses

The primary analysis will be done according to 'intention to treat' principle, using all obtained data irrespective of participant adherence to the intervention and completeness of outcome measures. We will use linear mixed models (LMMs) to compare primary and secondary continuous outcome measures between groups, with time and group x time interactions as fixed effects, and subject as random factor.[34] Since we expect that there will be no systematical differences between the groups at baseline, means at baseline will be constrained to be equal in the LMMs. We will report estimates for the difference (time x group) with corresponding 95% confidence intervals and p-values in the intervention group compared with the control group. We will check normality of residuals by visual inspection of QQ-plots and perform bootstrapping in cases of non-normal model residuals. For our primary outcome measure, we will consider a p-value $< 0.05$ as statistically significant. For the secondary outcome measures, $p < .01$ will be considered as statistically significant, due to multiple comparisons, and these analyses will be explorative. We will also perform per-protocol analyses; Women who have accumulated an average of 75 PAI per week during the preconception period and who have adhered to TRE at least 4 days per week during preconception, will be included in the per-protocol analyses for the outcomes measured at the end of follow-up. We will report additional results from all women who were included in the trial, from the pre-conception period, irrespective of whether they got pregnant or not during the study period (i.e., for all those who were included).

#### 9.5 Randomization

##### 9.5.1 Allocation- sequence generation

Key elements to specify regarding allocation of interventions are:

- Participants will be stratified according to if they have had GDM in a previous pregnancy (yes/no) and randomized to the intervention group or the control group after baseline testing, using a computer random number generator developed and administered at the Faculty of Medicine, Department of Public Health and general Practice, NTNU, Trondheim, Norway.

##### 9.5.2 Allocation- procedure to randomize a participant

The study will be announced through social media, at hospital and universities homepages, at local stores and public places. An invitation will be sent out to women aged 20-35 years old in Trondheim and surrounding area using the Norwegian population register, where we will use eFORSK to administer sending. The principal investigator and one trial co-ordinator will enroll participants and assign participants to intervention. A computer random number generator developed and administered at the Faculty of Medicine, Department of Public Health and general Practice, NTNU, Trondheim, Norway, have access to the allocation list. Allocation concealment mechanism prevents participants and site personnel from knowing the study group the next participant will be assigned to.

##### 9.5.3 Blinding and emergency unbinding

The study will not be blinded, as it is difficult to blind participants and treatment providers to behavior intervention. Baseline measurements will, however, be done before randomization.

### 10 STUDY MANAGEMENT

#### 10.1 Investigator Delegation Procedure

The principal investigator is responsible for making and updating a “delegation of tasks” listing all the involved co-workers and their role in the project. She will ensure that appropriate training relevant to the study is given to all of these staff, and that any new information of relevance to the performance of this study is forwarded to the staff involved.

#### 10.2 Protocol Adherence

Investigators ascertain they will apply due diligence to avoid protocol deviations.

All significant protocol deviations will be recorded and reported in the Clinical Study Report (CSR).

#### 10.3 Study Amendments

If it is necessary for the study protocol to be amended, the amendment and/or a new version of the study protocol (Amended Protocol) will be notified to and approved by the Competent Authority and the Ethics Committee according to EU and national regulations.

#### 10.4 Audit and Inspections

Authorized representatives of a Competent Authority and Ethics Committee may visit the centre to perform inspections, including source data verification. Likewise, the representatives from sponsor may visit the centre to perform an audit. The purpose of an audit or inspection is to systematically and independently examine all study-related activities and documents to determine whether these activities were conducted, and data were recorded, analysed, and accurately reported according to the protocol, Good Clinical Practice (ICH GCP), and any applicable regulatory requirements. The principal investigator will ensure that the inspectors and auditors will be provided with access to source data/documents.

### 11 ETHICAL AND REGULATORY REQUIREMENTS

The study will be conducted in accordance with ethical principles that have their origin in the Declaration of Helsinki and are consistent with ICH/Good Clinical Practice and applicable regulatory requirements. Registration of participant data will be carried out in accordance with national personal data laws.

#### 11.1 Ethics Committee Approval

The Regional Committee Medical Research Ethics, Norway, and The Human Research Ethics Committee (HREC) have approved the study.

The investigator is responsible for informing the ethics committee of any serious and unexpected adverse events and/or major amendments to the protocol as per national requirements.

#### 11.2 Other Regulatory Approvals

The protocol is submitted and approved by the applicable competent authorities before commencement of the study. The protocol is registered in www.clinicaltrials.gov

#### 11.3 Informed Consent Procedure

The investigator is responsible for giving the participants full and adequate verbal and written information about the nature, purpose, possible risk, and benefit of the study. They will be informed as to the strict confidentiality of their participant data, but that their medical records may be reviewed for trial purposes by authorized individuals other than their treating physician.

It will be emphasized that the participation is voluntary and that the participant is allowed to refuse further participation in the protocol whenever she/he wants. This will not prejudice the participants' subsequent care. Documented informed consent must be obtained for all participants included in the study before they are registered in the study. This will be done in accordance with the national and local regulatory requirements. The investigator is responsible for obtaining signed informed consent.

The signed and dated participant consent forms will be filed in the Investigator Site File binder.

#### 11.4 Subject Identification

The investigator is responsible for keeping a list of all participants (who have received study treatment or undergone any study specific procedure) including participant's date of birth and personal number, full names, and last known addresses.

The participant will be identified in the CRFs by participant number.

### 12 TRIAL SPONSORSHIP AND FINANCING

The trial is funded by the Novo Nordisk Foundation (NNF19SA058975) and by The Liaison Committee for education, research and innovation in Central Norway, and by The Joint Research Committee between St. Olavs hospital and the Faculty of Medicine and Health Sciences, NTNU (FFU).

The ultrasound part of the project is also funded by the Centre for Innovative Ultrasound Solutions (CIUS), a large research and innovation project led by NTNU. CIUS has more than 20 contributors including the Research Council of Norway, Helse Midt-Norge, St. Olavs hospital, NTNU, and GE Healthcare.

### 13 TRIAL INSURANCE

The Principal investigator has insurance coverage for this study through "Pasientskadeerstatningen".

### 14 PUBLICATION POLICY

Upon study completion and finalization of the study report, the results of this study will either be submitted for publication and/or posted in a publicly assessable database of clinical study results.

All personnel who have contributed significantly with the planning and performance of the study (Vancouver convention 1988) may be included in the list of authors.

### 16 LIST OF APPENDICES

A IPAQ

B Bakground information and PGWBI questionnaire (mother), Test 1

C Bakground information (mother), Test 2

D Bakground information (mother), Test 3

E Bakground information (mother), Test 4

F The Horne-Östberg Morningness-Eveningness Questionnaire

G PSQI

H Bakground information (father), Test 1

I Background information (father), Test 2

### APPENDIX A – INTERNATIONAL PHYSICAL ACTIVITY QUESTIONNAIRE (IPAQ)

#### INTERNATIONAL PHYSICAL ACTIVITY QUESTIONNAIRE

We are interested in finding out about the kinds of physical activities that people do as part of their everyday lives. The questions will ask you about the time you spent being physically active in the **last 7 days**. Please answer each question even if you do not consider yourself to be an active person. Please think about the activities you do at work, as part of your house and yard work, to get from place to place, and in your spare time for recreation, exercise or sport.

Think about all the **vigorous** activities that you did in the **last 7 days**. **Vigorous** physical activities refer to activities that take hard physical effort and make you breathe much harder than normal. Think *only* about those physical activities that you did for at least 10 minutes at a time.

1. During the **last 7 days**, on how many days did you do **vigorous** physical activities like heavy lifting, digging, aerobics, or fast bicycling?

   _____ **days per week**

   ☐ No vigorous physical activities → ***Skip to question 3***

2. How much time did you usually spend doing **vigorous** physical activities on one of those days?

   _____ **hours per day**

   _____ **minutes per day**

   ☐ Don't know/Not sure

Think about all the **moderate** activities that you did in the **last 7 days**. **Moderate** activities refer to activities that take moderate physical effort and make you breathe somewhat harder than normal. Think only about those physical activities that you did for at least 10 minutes at a time.

3. During the **last 7 days**, on how many days did you do **moderate** physical activities like carrying light loads, bicycling at a regular pace, or doubles tennis? Do not include walking.

   _____ **days per week**

   ☐ No moderate physical activities → ***Skip to question 5***

**SHORT LAST 7 DAYS SELF-ADMINISTERED** version of the IPAQ. Revised August 2002.

4. How much time did you usually spend doing **moderate** physical activities on one of those days?

_____ **hours per day**

_____ **minutes per day**

☐ Don't know/Not sure

Think about the time you spent **walking** in the **last 7 days**. This includes at work and at home, walking to travel from place to place, and any other walking that you have done solely for recreation, sport, exercise, or leisure.

5. During the **last 7 days**, on how many days did you **walk** for at least 10 minutes at a time?

_____ **days per week**

☐ No walking → ***Skip to question 7***

6. How much time did you usually spend **walking** on one of those days?

_____ **hours per day**

_____ **minutes per day**

☐ Don't know/Not sure

The last question is about the time you spent **sitting** on weekdays during the **last 7 days**. Include time spent at work, at home, while doing course work and during leisure time. This may include time spent sitting at a desk, visiting friends, reading, or sitting or lying down to watch television.

7. During the **last 7 days**, how much time did you spend **sitting** on a **week day**?

_____ **hours per day**

_____ **minutes per day**

☐ Don't know/Not sure

**This is the end of the questionnaire, thank you for participating.**

SHORT LAST 7 DAYS SELF-ADMINISTERED version of the IPAQ. Revised August 2002.

### APPENDIX B – BACKGROUND INFORMATION AND PSYCHOLOGICAL GENERAL WELL-BEING INDEX (PGWBI) QUESTIONNAIRE (MOTHER), TEST 1

BEFORE THE BEGINNING – TEST 1 Initials ☐☐☐☐ ID-number ☐☐☐

Attached is a questionnaire that we would like you to fill out. Your answer is an important contribution to understanding the importance of your overall health and several background variables in relation to other measurements in the study.

Please answer all the questions. Do not spend too long on each question. The first thing that comes to your mind is usually the correct answer.

Your answer must appear clearly. Use black or dark blue pen for filling. If you answer incorrectly, completely fill the square and check the correct box. The form must not be folded.

When you`re done filling out the questionnaires, deliver the form to one of the research workers.

Today`s date *(ddmmyyyy)*: ☐☐·☐☐·☐☐☐☐

**BACKGROUND INFORMATION**

**1) Date of birth:** ☐☐·☐☐·☐☐☐☐

**2) Ethnicity**

☐ Norwegian

☐ Other: ……………………………………………….

**3) Highest educational level**

☐ Compulsory schooling

☐ Upper secondary school

☐ University, less than 4 years

☐ University, 4 years or more

**4) Paid employment?**

(Also include temporary – and short-term employment. Don't consider whether you are off sick or on leave. Paid employment also include work without a fixed salary in a company owned by the family, e.g. farm / business)

☐ Yes, I`m ☐☐☐ % employed

☐ No, I`m a ☐ Student

☐ Disabled

☐ Homemaker

☐ Unemployed act

**5) What kind of work do you do?**

☐ Health/social work ☐ Teaching/research

☐ Office ☐ Industry

☐ Construction and building ☐ Shop assistant

☐ Service trade ☐ Other: ……………………………………………….

**6) Working hours?**

☐ Daytime

☐ Afternoon/evening

☐ Shift work or rotation scheme

☐ No fixed rotation scheme (extra help, stand-in etc.)

**7) Do you work on foot or standing?**

☐ Yes, daily, more than half of the hours worked

☐ Yes, daily, less than half of the hours worked

☐ Periodic, but not daily

☐ Rarely or never

**8) How active are you at your work?**

☐ Sedentary (Sit all day)

☐ Partly sedentary (Walk or stand part of the day)

☐ Active (Walk or stand all day)

☐ Very active

**9) In general, how would you rate your health today?**

☐ Very good

☐ Good

☐ Moderate

☐ Bad

☐ Very bad

**10) "I perceive stress in life, both personally and at work…"**

☐ Very often

☐ Often

☐ Sometimes

☐ Rarely or Never

**11) How important is it for you to exercise and maintain a healthy diet?**
Scale from 1 – 10 (1 = not important at all, and 10 = very important)

| | | | | | | | | | |
|---|---|---|---|---|---|---|---|---|---|
| 1 | 2 | 3 | 4 | 5 | 6 | 7 | 8 | 9 | 10 |

**12) If you consider exercising and maintaining a healthy diet important, do you believe it`s feasible for you to maintain that lifestyle?**
Scale from 1 – 10 (1 = no confidence, and 10 = highly confident)

| | | | | | | | | | |
|---|---|---|---|---|---|---|---|---|---|
| 1 | 2 | 3 | 4 | 5 | 6 | 7 | 8 | 9 | 10 |

**13) Do you smoke or use "snus"?**

☐ Yes, ca. ☐☐ cigarettes daily

☐ Yes, ca. ☐☐ «snus» daily

☐ No

**14) Have you smoked in the past?**

☐ Yes

☐ No (proceed to question 17)

**15) If so, how many years have you smoked in total?**

☐☐ Years

**16) If so, when did you quit smoking?**

☐☐ years ago

**17) How often have you been drinking alcohol (beer, wine or liquor) in the LAST 14 DAYS?**

☐ I have not been drinking, but is not teetotal

☐ 1 - 4 times

☐ 5 – 10 times

☐ More than 10 times

☐ Teetotal, never drink alcohol (proceed to question 19)

**18) If you have been drinking alcohol for the last 14 days, have you ever felt drunk?**

☐ Yes

☐ No

**19) Have there been periods in your life where you have been drinking too much?**

☐ No

☐ Maybe

☐ Yes

**20) Do you use any medications?**

☐ Yes, Specify type……………………………………………………………………………..

☐ No

**21) How is your menstrual cycle?**

☐ Normal cycle length (*less than 35 days between the first day of menstruation to the first day of the next period and / or 10 or more periods per year)*

☐ Cycle length 35 - 42 days (8-0 menstruation/year)

☐ Cycle length 42 days – 6 months (2-7 menstruation/year)

☐ Cycle length over 6 months (0-1 menstruation/year)

**22) Children:**

☐ Yes, I have given birth to ☐ children

☐ No (proceed to question 25)

**23) If so, was it:**

☐ Natural

☐ Infertility treatment

☐ Both natural and infertility treatment

**24) Enter information about your previous birth / births:**

**a) 1. Child:**

**Date of birth:** ☐☐·☐☐·☐☐☐☐ **Birth weight of the child:** ☐☐☐☐gram
*(ddmmyyyy)*

**Ultrasound term:** ☐☐·☐☐·☐☐☐☐ **Your weight gain during pregnancy:** ☐☐kg
*(ddmmyyyy)*

**Complications during pregnancy?**

☐ Severe preeclampsia ☐ Gestational diabetes

☐ Severe fetal growth retardation ☐ Premature birth (before week 34)

**Type of birth:**

☐ Normal vaginal birth without complications

☐ Vacuum or forceps

☐ Breech position

☐ C - section

☐ Other: ...............................................

**Potential complications during labor:**

☐ None

☐ Partly rectal tear

☐ Rectal tear

☐ Do not know

☐ Other: .......................................

**b) 2. Child:**

**Date of birth:** ☐☐.☐☐.☐☐☐☐ *(ddmmyyyy)* **Birth weight of the child:** ☐☐☐☐gram

**Ultrasound term:** ☐☐.☐☐.☐☐☐☐ *(ddmmyyyy)* **Your weight gain during pregnancy:** ☐☐kg

**Complications during pregnancy?**

☐ Severe preeclampsia

☐ Gestational diabetes

☐ Severe fetal growth retardation

☐ Premature birth (before week 34)

**Type of birth:**

☐ Normal vaginal birth without complications

☐ Vacuum or forceps

☐ Breech position

☐ C - section

☐ Other: ...............................................

**Potential complications during labor:**

☐ None

☐ Partly rectal tear

☐ Rectal tear

☐ Do not know

☐ Other: .......................................

c) 3. Child:

**Date of birth:** □□.□□.□□□□ **Birth weight of the child:** □□□□gram
*(ddmmyyyy)*

**Ultrasound term:** □□.□□.□□□□ **Your weight gain during pregnancy:** □□kg
*(ddmmyyyy)*

**Complications during pregnancy?**

- □ Severe preeclampsia
- □ Gestational diabetes
- □ Severe fetal growth retardation
- □ Premature birth (before week 34)

**Type of birth:**

- □ Normal vaginal birth without complications
- □ Vacuum or forceps
- □ Breech position
- □ C - section
- □ Other: ..............................................

**Potential complications during labor:**

- □ None
- □ Partly rectal tear
- □ Rectal tear
- □ Do not know
- □ Other: ....................................

**25) Have you ever been treated for involuntary childlessness?**

- □ Yes
- □ No (proceed to question 27)

**26) Is so, what kind of treatment was it?**

- □ Ovarian surgery (operation on the fallopian tubes, surgery on the uterus)
- □ Another form of surgery
- □ Medicines for endometriosis
- □ Ovulation induction (hormone therapy)
- □ Insemination (injection of semen)
- □ IVF treatment
- □ Other: ....................................................................................................

**27) Have you tried to get pregnant before? (A year back)**

☐ Yes

☐ No (proceed to question 29)

**28) Is so, how long have you been trying to get pregnant?**

☐ ☐ Days

☐ ☐ Months

☐ ☐ Years

**29) Have you experienced spontaneous abortion?**

Yes, I have experienced ☐☐ spontaneous abortions

☐ No

#### WELLNESS QUESTIONS

**30) How have you been feeling in general? (DURING THE PAST WEEK)**

- ☐ In excellent spirits
- ☐ In very good spirits
- ☐ In good spirits mostly
- ☐ I have been up and down in spirits a lot
- ☐ In low spirits mostly
- ☐ In very low spirits

**31) How often were you bothered by any illness, bodily disorder, aches or pains? (DURING THE PAST WEEK)**

- ☐ Every day
- ☐ Almost every day
- ☐ About half of the time
- ☐ Now and then, but less than half the time
- ☐ In low spirits mostly
- ☐ In very low spirits

**32) Did you feel depressed? (DURING THE PAST WEEK)**

- ☐ Yes – to the point that I felt like taking my life
- ☐ Yes – to the point that I did not care about anything
- ☐ Yes – very depressed almost every day
- ☐ Yes – quite depressed almost every day
- ☐ Yes – a little depressed now and then
- ☐ No – never felt depressed at all

**33) Have you been in firm control of your behavior, thoughts, emotions, or feelings? (DURING THE PAST WEEK)**

☐ Yes, definitely so

☐ Yes, for the most part

☐ Generally so

☐ Not too well

☐ No, and I am somewhat disturbed

☐ No, and I am very disturbed

**34) Have you been bothered by nervousness or your "nerves"? (DURING THE PAST WEEK)**

☐ Extremely so - to the point where I could not work or take care of things

☐ Very much so

☐ Quite a bit

☐ Some-enough to bother me

☐ A little

☐ Not at all

**35) How much energy, pop, or vitality did you have or feel? (DURING THE PAST WEEK)**

☐ Very full of energy – lots of pep

☐ Fairly energetic most of the time

☐ My energy level varied quite a bit

☐ Generally low in energy or pep

☐ Very low in energy or pep most of the time

☐ No energy or pep at all – I felt drained, sapped

**36) I felt downhearted and blue DURING THE PAST WEEK.**

☐ None of the time

☐ A little of the time

☐ Some of the time

☐ A good bit of the time

☐ Most of the time

☐ All the time

**37) Were you generally tense-or did you feel any tension? (DURING THE PAST WEEK)**

☐ Yes – extremely tense, most or all the time

☐ Yes – very tense most of the time

☐ Not generally tense, but did feel fairly tense several times

☐ I felt a little tense a few times

☐ My general tension level was quite low

☐ I never felt tense or any tension at all

**38) How happy, satisfied, or pleased have you been with your personal life? (DURING THE PAST WEEK)**

☐ Extremely happy – could not have been more satisfied or pleased

☐ Very happy most of the time

☐ Generally satisfied - pleased

☐ Sometimes fairly happy, sometimes fairly unhappy

☐ Generally dissatisfied, unhappy

☐ Very dissatisfied or unhappy most or all the time

**39) Did you feel healthy enough to carry out the things you like to do or had to do? (DURING THE PAST WEEK)**

☐ Yes – definitely

☐ For the most part

☐ Health problems limited me in some important ways

☐ I was only healthy enough to take care of myself

☐ I needed some help in taking care of myself

☐ I needed someone to help me with most or all of the things I had to do

**40) Have you felt so sad, discouraged, hopeless, or had so many problems that you wondered if anything was worthwhile? (DURING THE PAST WEEK)**

☐ Extremely so-to the point that I have just about given up

☐ Very much so

☐ Quite a bit

☐ Some - enough to bother me

☐ A little bit

☐ Not at all

**41) I woke up feeling fresh and rested DURING THE PAST WEEK**

☐ None of the time

☐ A little of the time

☐ Some of the time

☐ A good bit of the time

☐ Most of the time

☐ All the time

**42) Have you been concerned, worried, or had any fears about your health? (DURING THE PAST WEEK)**

☐ Extremely so

☐ Very much so

☐ Quite a bit

☐ Some, but not a lot

☐ Practically never

☐ Not at all

**43) Have you had any reason to wonder if you were losing your mind, or losing control over the way act, talk, think, feel or of your memory? (DURING THE PAST WEEK)**

☐ Not at all

☐ Only a little

☐ Some – but not enough to be concerned or worried about

☐ Some and I have been a little concerned

☐ Some and I am quite concerned

☐ Yes, very much so and I am very concerned

**44) My daily life was full of things that were interesting to me DURING THE PAST WEEK**

☐ None of the time

☐ A little of the time

☐ Some of the time

☐ A good bit of the time

☐ Most of the time

☐ All the time

**45) Did you feel active, vigorous, or dull, sluggish? (DURING THE PAST WEEK)**

☐ Very active, vigorous every day

☐ Mostly active, vigorous – never really dull, sluggish

☐ Fairly active, vigorous – seldom dull, sluggish

☐ Fairly dull, sluggish - seldom active, vigorous

☐ Mostly dull, sluggish – never really active, vigorous

☐ Very dull, sluggish every day

**46) Have you been anxious, worried, or upset? (DURING THE PAST WEEK)**

☐ Extremely so – to the point of being sick or almost sick

☐ Very much so

☐ Quite a bit

☐ Some – enough to bother me

☐ A little bit

☐ Not at all

**47) I was emotionally stable and sure of myself DURING THE PAST WEEK**

☐ None of the time

☐ A little of the time

☐ Some of the time

☐ A good bit of the time

☐ Most of the time

☐ All of the time

**48) Did you feel relaxed, at ease or high strung, tight, or keyed-up? (DURING THE PAST WEEK)**

☐ Felt relaxed and at ease the whole month

☐ Felt relaxed and at ease most of the time

☐ Generally felt relaxed but at time felt fairly high strung

☐ Generally felt high strung but at time felt fairly relaxed

☐ Felt high strung, tight, or keyed up most of the time

☐ Felt high strung, tight, or keyed up the whole month

**49) I felt cheerful, lighthearted DURING THE PAST WEEK**

☐ None of the time

☐ A little of the time

☐ Some of the time

☐ A good bit of the time

☐ Most of the time

☐ All of the time

**50) I felt tired, worn out, used up, or exhausted DURING THE PAST WEEK**

☐ None of the time

☐ A little of the time

☐ Some of the time

☐ A good bit of the time

☐ Most of the time

☐ All of the time

**51) Have you been under or felt you were under any strain, stress, or pressure? (DURING THE PAST WEEK)**

☐ Yes, almost more than I could bear or stand

☐ Yes, quite a bit of pressure

☐ Yes, some – more than usual

☐ Yes, some – but about usual

☐ Yes, a little

☐ Not at all

#### CASE HISTORY

**52) Do you have any diseases?**

☐ Yes:_____________________________________________________________

_________________________________________________________________

_________________________________________________________________

☐ No

**53) Are there any close relatives who have:**

☐ Diabetes type 1 ☐ High blood pressure

☐ Diabetes type 2 ☐ Overweight/obesity

☐ None of the above

### APPENDIX C – BACKGROUND INFORMATION (MOTHER), TEST 2

BEFORE THE BEGINNING – TEST 2 Initials ☐☐☐☐ ID-number ☐☐☐

Attached is a questionnaire that we would like you to fill out. Your answer is an important contribution to understanding the importance of your overall health and several background variables in relation to other measurements in the study.

Please answer all the questions. Do not spend too long on each question. The first thing that comes to your mind is usually the correct answer.

Your answer must appear clearly. Use black or dark blue pen for filling. If you answer incorrectly, completely fill the square and check the correct box. The form must not be folded.

When you`re done filling out the questionnaires, deliver the form to one of the research workers.

Today`s date *(ddmmyyyy)*: ☐☐·☐☐·☐☐☐☐

#### BACKGROUND INFORMATION

**1) In general, how would you rate your health today?**

☐ Very good

☐ Good

☐ Moderate

☐ Bad

☐ Very bad

**2) "I perceive stress in life, both personally and at work…"**

☐ Very often

☐ Often

☐ Sometimes

☐ Rarely or Never

**3) How important is it for you to exercise and maintain a healthy diet?**
Scale from 1 – 10 (1 = not important at all, and 10 = very important)

| 1 | 2 | 3 | 4 | 5 | 6 | 7 | 8 | 9 | 10 |
|---|---|---|---|---|---|---|---|---|---|

**4) If you consider exercising and maintaining a healthy diet important, do you believe it`s feasible for you to maintain that lifestyle?**
Scale from 1 – 10 (1 = no confidence, and 10 = highly confident)

| 1 | 2 | 3 | 4 | 5 | 6 | 7 | 8 | 9 | 10 |
|---|---|---|---|---|---|---|---|---|---|

**5) Do you smoke or use "snus"?**

☐ Yes, ca. ☐☐ cigarettes daily

☐ Yes, ca. ☐☐ «snus» daily

☐ No

**6) How often have you been drinking alcohol (beer, wine or liquor) in the LAST 14 DAYS?**

☐ I have not been drinking, but is not teetotal (Proceed to question 8)

☐ 1 - 4 time

☐ 5 – 10 times

☐ More than 10 times

☐ Teetotal, never drink alcohol (Proceed to question 8)

**7) If you have been drinking alcohol for the last 14 days, have you ever felt drunk?**

☐ Yes

☐ No

**8) Have you started or stopped on any medications since last time?**

☐ Yes, I have stopped. Specify type…………………………………………………………………………..

☐ Yes, I have started. Specify type…………………………………………………………………………..

☐ No, I haven't started / stopped on any medication since last time

#### WELLNESS QUESTIONS

**9) How have you been feeling in general? (DURING THE PAST WEEK)**

☐ In excellent spirits

☐ In very good spirits

☐ In good spirits mostly

☐ I have been up and down in spirits a lot

☐ In low spirits mostly

☐ In very low spirits

**10) How often were you bothered by any illness, bodily disorder, aches or pains? (DURING THE PAST WEEK)**

☐ Every day

☐ Almost every day

☐ About half of the time

☐ Now and then, but less than half the time

☐ In low spirits mostly

☐ In very low spirits

**11) Did you feel depressed? (DURING THE PAST WEEK)**

☐ Yes – to the point that I felt like taking my life

☐ Yes – to the point that I did not care about anything

☐ Yes – very depressed almost every day

☐ Yes – quite depressed almost every day

☐ Yes – a little depressed now and then

☐ No – never felt depressed at all

**12) Have you been in firm control of your behavior, thoughts, emotions, or feelings? (DURING THE PAST WEEK)**

☐ Yes, definitely so

☐ Yes, for the most part

☐ Generally so

☐ Not too well

☐ No, and I am somewhat disturbed

☐ No, and I am very disturbed

**13) Have you been bothered by nervousness or your "nerves"? (DURING THE PAST WEEK)**

☐ Extremely so - to the point where I could not work or take care of things

☐ Very much so

☐ Quite a bit

☐ Some-enough to bother me

☐ A little

☐ Not at all

**14) How much energy, pop, or vitality did you have or feel? (DURING THE PAST WEEK)**

☐ Very full of energy – lots of pep

☐ Fairly energetic most of the time

☐ My energy level varied quite a bit

☐ Generally low in energy or pep

☐ Very low in energy or pep most of the time

☐ No energy or pep at all – I felt drained, sapped

**15) I felt downhearted and blue DURING THE PAST WEEK.**

☐ None of the time

☐ A little of the time

☐ Some of the time

☐ A good bit of the time

☐ Most of the time

☐ All of the time

**16) Were you generally tense-or did you feel any tension? (DURING THE PAST WEEK)**

☐ Yes – extremely tense, most or all of the time

☐ Yes – very tense most of the time

☐ Not generally tense, but did feel fairly tense several times

☐ I felt a little tense a few times

☐ My general tension level was quite low

☐ I never felt tense or any tension at all

**17) How happy, satisfied, or pleased have you been with your personal life? (DURING THE PAST WEEK)**

☐ Extremely happy – could not have been more satisfied or pleased

☐ Very happy most of the time

☐ Generally satisfied - pleased

☐ Sometimes fairly happy, sometimes fairly unhappy

☐ Generally dissatisfied, unhappy

☐ Very dissatisfied or unhappy most or all the time

**18) Did you feel healthy enough to carry out the things you like to do or had to do? (DURING THE PAST WEEK)**

☐ Yes – definitely so

☐ For the most part

☐ Health problems limited me in some important ways

☐ I was only healthy enough to take care of myself

☐ I needed some help in taking care of myself

☐ I needed someone to help me with most or all of the things I had to do

**19) Have you felt so sad, discouraged, hopeless, or had'so many problems that you wondered if anything was worthwhile? (DURING THE PAST WEEK)**

☐ Extremely so-to the point that I have just about given up

☐ Very much so

☐ Quite a bit

☐ Some - enough to bother me

☐ A little bit

☐ Not at all

**20) I woke up feeling fresh and rested DURING THE PAST WEEK**

☐ None of the time

☐ A little of the time

☐ Some of the time

☐ A good bit of the time

☐ Most of the time

☐ All of the time

**21) Have you been concerned, worried, or had any fears about your health? (DURING THE PAST WEEK)**

☐ Extremely so

☐ Very much so

☐ Quite a bit

☐ Some, but not a lot

☐ Practically never

☐ Not at all

**22) Have you had any reason to wonder if you were losing your mind, or losing control over the way act, talk, think, feel or of your memory? (DURING THE PAST WEEK)**

☐ Not at all

☐ Only a little

☐ Some – but not enough to be concerned or worried about

☐ Some and I have been a little concerned

☐ Some and I am quite concerned

☐ Yes, very much so and I am very concerned

**23) My daily life was full of things that were interesting to me DURING THE PAST WEEK**

☐ None of the time

☐ A little of the time

☐ Some of the time

☐ A good bit of the time

☐ Most of the time

☐ All of the time

**24) Did you feel active, vigorous, or dull, sluggish? (DURING THE PAST WEEK)**

☐ Very active, vigorous every day

☐ Mostly active, vigorous – never really dull, sluggish

☐ Fairly active, vigorous – seldom dull, sluggish

☐ Fairly dull, sluggish - seldom active, vigorous

☐ Mostly dull, sluggish – never really active, vigorous

☐ Very dull, sluggish every day

**25) Have you been anxious, worried, or upset? (DURING THE PAST WEEK)**

☐ Extremely so – to the point of being sick or almost sick

☐ Very much so

☐ Quite a bit

☐ Some – enough to bother me

☐ A little bit

☐ Not at all

**26) I was emotionally stable and sure of myself DURING THE PAST WEEK**

☐ None of the time

☐ A little of the time

☐ Some of the time

☐ A good bit of the time

☐ Most of the time

☐ All of the time

**27) Did you feel relaxed, at ease or high strung, tight, or keyed-up? (DURING THE PAST WEEK)**

☐ Felt relaxed and at ease the whole month

☐ Felt relaxed and at ease most of the time

☐ Generally felt relaxed but at time felt fairly high strung

☐ Generally felt high strung but at time felt fairly relaxed

☐ Felt high strung, tight, or keyed up most of the time

☐ Felt high strung, tight, or keyed up the whole month

**28) I felt cheerful, lighthearted DURING THE PAST WEEK**

☐ None of the time

☐ A little of the time

☐ Some of the time

☐ A good bit of the time

☐ Most of the time

☐ All of the time

**29) I felt tired, worn out, used up, or exhausted DURING THE PAST WEEK**

☐ None of the time

☐ A little of the time

☐ Some of the time

☐ A good bit of the time

☐ Most of the time

☐ All of the time

**30) Have you been under or felt you were under any strain, stress, or pressure? (DURING THE PAST WEEK)**

☐ Yes, almost more than I could bear or stand

☐ Yes, quite a bit of pressure

☐ Yes, some – more than usual

☐ Yes, some – but about usual

☐ Yes, a little

☐ Not at all

### APPENDIX D – BACKGROUND INFORMATION (MOTHER), TEST 3

BEFORE THE BEGINNING – TEST 3 Initials □□□□ ID-number □□□

Attached is a questionnaire that we would like you to fill out. Your answer is an important contribution to understanding the importance of your overall health in relation to your pregnancy and gestational diabetes.

Please answer all the questions. Do not spend too long on each question. The first thing that comes to your mind is usually the correct answer.

Your answer must appear clearly. Use black or dark blue pen for filling. If you answer incorrectly, completely fill the square and check the correct box. The form must not be folded.

When you`re done filling out the questionnaires, deliver the form to one of the research workers.

Today`s date *(ddmmyyyy)*: □□·□□·□□□□

#### BACKGROUND INFORMATION

**1) In general, how would you rate your health today?**

□ Very good

□ Good

□ Moderate

□ Bad

□ Very bad

**2) "I perceive stress in life, both personally and at work…"**

□ Very often

□ Often

□ Sometimes

□ Rarely or Never

**3) How important is it for you to exercise and maintain a healthy diet?**
Scale from 1 – 10 (1 = not important at all, and 10 = very important)

| 1 | 2 | 3 | 4 | 5 | 6 | 7 | 8 | 9 | 10 |
|---|---|---|---|---|---|---|---|---|---|

**4) If you consider exercising and maintaining a healthy diet important, do you believe it`s feasible for you to maintain that lifestyle?**
Scale from 1 – 10 (1 = no confidence, and 10 = highly confident)

| 1 | 2 | 3 | 4 | 5 | 6 | 7 | 8 | 9 | 10 |
|---|---|---|---|---|---|---|---|---|---|

**5) Do you smoke or use "snus"?**

☐ Yes, ca. ☐☐ cigarettes daily

☐ Yes, ca. ☐☐ «snus» daily

☐ No

**6) Have you started or stopped on any medications since last time?**

☐ Yes, I have stopped. Specify type………………………………………………………………………

☐ Yes, I have started. Specify type………………………………………………………………………

☐ No, I haven't started / stopped on any medication since last time

**7) a) Term:** ☐☐.☐☐.☐☐☐☐ **b) Gestational week <u>NOW</u>:** ☐☐

**8) Did you have any trouble getting pregnant?**

☐ Yes

☐ No

**9) How long did it take from your decision to try to get pregnant, until the pregnancy occurred?**

*(write 0 months if the pregnancy occurred at the first attempt)* ☐☐ months

**10) How were your menstrual cycle during the period before the pregnancy occurred?**

☐ Normal cycle length (*less than 35 days between the first day of menstruation to the first day of the next period and / or 10 or more periods per year)*

☐ Cycle length 35 - 42 days (8-0 menstruation/year)

☐ Cycle length 42 days – 6 months (2-7 menstruation/year)

☐ Cycle length over 6 months (0-1 menstruation/year)

**11) Have you had any bleeding in this pregnancy?**

☐ Yes

☐ No (proceed to question 13)

**12) If you have had bleedings during this pregnancy, please state the date when the bleeding started, how many days it lasted and how much you bled:**

| **Date** *(ddmmyyyy)* | **Duration of bleeding** (number of days) | **Amount of blood** |
|---|---|---|
| **Date 1. bleeding** | | |
| ☐☐.☐☐.☐☐☐☐ | ☐☐ | ☐ Spotting<br>☐ More than spotting |
| **Date 2. bleeding** | | |
| ☐☐.☐☐.☐☐☐☐ | ☐☐ | ☐ Spotting<br>☐ More than spotting |
| **Date 3. bleeding** | | |
| ☐☐.☐☐.☐☐☐☐ | ☐☐ | ☐ Spotting<br>☐ More than spotting |

**13) Have you had any nausea during this pregnancy?**

☐ Yes

☐ No (proceed to question 15)

**14) If you have had pregnancy sickness during this pregnancy, please state the date when the nausea started and how long it lasted (if you are still sick, leave it blank):**

Nausea started (dd.mm.yyyy) ☐☐.☐☐.☐☐☐☐

Nausea lasted until (dd.mm.yyyy) ☐☐.☐☐.☐☐☐☐

**15) Have you been off sick during this pregnancy?**

☐ Yes

☐ No (proceed to question 17)

**16) If you have been off sick during this pregnancy, please specify below:**

a) Reported sick, from (ddmmyyyy): ☐☐.☐☐.☐☐☐☐

until (ddmmyyyy): ☐☐.☐☐.☐☐☐☐

b) Sickness absence percentage: ☐☐☐ %

c) Are you actively on sick leave? ☐ Yes ☐ No

d) Reason for sick leave:________________________________________

____________________________________________________

____________________________________________________

e) Comments:____________________________________________

____________________________________________________

____________________________________________________

**17) Have you maintained the same level of physical activity as before your pregnancy?**

☐ I was more active before pregnancy

☐ I am as active as before the pregnancy

☐ I am more active now than before pregnancy

#### WELLNESS QUESTIONS

**18) How have you been feeling in general? (DURING THE PAST WEEK)**

☐ In excellent spirits

☐ In very good spirits

☐ In good spirits mostly

☐ I have been up and down in spirits a lot

☐ In low spirits mostly

☐ In very low spirits

**19) How often were you bothered by any illness, bodily disorder, aches or pains? (DURING THE PAST WEEK)**

☐ Every day

☐ Almost every day

☐ About half of the time

☐ Now and then, but less than half the time

☐ In low spirits mostly

☐ In very low spirits

**20) Did you feel depressed? (DURING THE PAST WEEK)**

☐ Yes – to the point that I felt like taking my life

☐ Yes – to the point that I did not care about anything

☐ Yes – very depressed almost every day

☐ Yes – quite depressed almost every day

☐ Yes – a little depressed now and then

☐ No – never felt depressed at all

**21) Have you been in firm control of your behavior, thoughts, emotions, or feelings? (DURING THE PAST WEEK)**

☐ Yes, definitely so

☐ Yes, for the most part

☐ Generally so

☐ Not too well

☐ No, and I am somewhat disturbed

☐ No, and I am very disturbed

**22) Have you been bothered by nervousness or your "nerves"? (DURING THE PAST WEEK)**

☐ Extremely so - to the point where I could not work or take care of things

☐ Very much so

☐ Quite a bit

☐ Some-enough to bother me

☐ A little

☐ Not at all

**23) How much energy, pop, or vitality did you have or feel? (DURING THE PAST WEEK)**

☐ Very full of energy – lots of pep

☐ Fairly energetic most of the time

☐ My energy level varied quite a bit

☐ Generally low in energy or pep

☐ Very low in energy or pep most of the time

☐ No energy or pep at all – I felt drained, sapped

**24) I felt downhearted and blue DURING THE PAST WEEK.**

☐ None of the time

☐ A little of the time

☐ Some of the time

☐ A good bit of the time

☐ Most of the time

☐ All of the time

**25) Were you generally tense-or did you feel any tension? (DURING THE PAST WEEK)**

☐ Yes – extremely tense, most or all of the time

☐ Yes – very tense most of the time

☐ Not generally tense, but did feel fairly tense several times

☐ I felt a little tense a few times

☐ My general tension level was quite low

☐ I never felt tense or any tension at all

**26) How happy, satisfied, or pleased have you been with your personal life? (DURING THE PAST WEEK)**

☐ Extremely happy – could not have been more satisfied or pleased

☐ Very happy most of the time

☐ Generally satisfied - pleased

☐ Sometimes fairly happy, sometimes fairly unhappy

☐ Generally dissatisfied, unhappy

☐ Very dissatisfied or unhappy most or all the time

**27) Did you feel healthy enough to carry out the things you like to do or had to do? (DURING THE PAST WEEK)**

☐ Yes – definitely so

☐ For the most part

☐ Health problems limited me in some important ways

☐ I was only healthy enough to take care of myself

☐ I needed some help in taking care of myself

☐ I needed someone to help me with most or all of the things I had to do

**28) Have you felt so sad, discouraged, hopeless, or had so many problems that you wondered if anything was worthwhile? (DURING THE PAST WEEK)**

- [ ] Extremely so-to the point that I have just about given up
- [ ] Very much so
- [ ] Quite a bit
- [ ] Some - enough to bother me
- [ ] A little bit
- [ ] Not at all

**29) I woke up feeling fresh and rested DURING THE PAST WEEK**

- [ ] None of the time
- [ ] A little of the time
- [ ] Some of the time
- [ ] A good bit of the time
- [ ] Most of the time
- [ ] All of the time

**30) Have you been concerned, worried, or had any fears about your health? (DURING THE PAST WEEK)**

- [ ] Extremely so
- [ ] Very much so
- [ ] Quite a bit
- [ ] Some, but not a lot
- [ ] Practically never
- [ ] Not at all

**31) Have you had any reason to wonder if you were losing your mind, or losing control over the way act, talk, think, feel or of your memory? (DURING THE PAST WEEK)**

☐ Not at all

☐ Only a little

☐ Some – but not enough to be concerned or worried about

☐ Some and I have been a little concerned

☐ Some and I am quite concerned

☐ Yes, very much so and I am very concerned

**32) My daily life was full of things that were interesting to me DURING THE PAST WEEK**

☐ None of the time

☐ A little of the time

☐ Some of the time

☐ A good bit of the time

☐ Most of the time

☐ All of the time

**33) Did you feel active, vigorous, or dull, sluggish? (DURING THE PAST WEEK)**

☐ Very active, vigorous every day

☐ Mostly active, vigorous – never really dull, sluggish

☐ Fairly active, vigorous – seldom dull, sluggish

☐ Fairly dull, sluggish - seldom active, vigorous

☐ Mostly dull, sluggish – never really active, vigorous

☐ Very dull, sluggish every day

**34) Have you been anxious, worried, or upset? (DURING THE PAST WEEK)**

☐ Extremely so – to the point of being sick or almost sick

☐ Very much so

☐ Quite a bit

☐ Some – enough to bother me

☐ A little bit

☐ Not at all

**35) I was emotionally stable and sure of myself DURING THE PAST WEEK**

☐ None of the time

☐ A little of the time

☐ Some of the time

☐ A good bit of the time

☐ Most of the time

☐ All of the time

**36) Did you feel relaxed, at ease or high strung, tight, or keyed-up? (DURING THE PAST WEEK)**

☐ Felt relaxed and at ease the whole month

☐ Felt relaxed and at ease most of the time

☐ Generally felt relaxed but at time felt fairly high strung

☐ Generally felt high strung but at time felt fairly relaxed

☐ Felt high strung, tight, or keyed up most of the time

☐ Felt high strung, tight, or keyed up the whole month

**37) I felt cheerful, lighthearted DURING THE PAST WEEK**

☐ None of the time

☐ A little of the time

☐ Some of the time

☐ A good bit of the time

☐ Most of the time

☐ All of the time

**38) I felt tired, worn out, used up, or exhausted DURING THE PAST WEEK**

☐ None of the time

☐ A little of the time

☐ Some of the time

☐ A good bit of the time

☐ Most of the time

☐ All of the time

**39) Have you been under or felt you were under any strain, stress, or pressure? (DURING THE PAST WEEK)**

☐ Yes, almost more than I could bear or stand

☐ Yes, quite a bit of pressure

☐ Yes, some – more than usual

☐ Yes, some – but about usual

☐ Yes, a little

☐ Not at all

### APPENDIX E – BACKGROUND INFORMATION (MOTHER), TEST 4

BEFORE THE BEGINNING – TEST 4 Initials ☐☐☐☐ ID-number ☐☐☐

Attached is a questionnaire that we would like you to fill out. Your answer is an important contribution to understanding the importance of your overall health in relation to your pregnancy and gestational diabetes.

Please answer all the questions. Do not spend too long on each question. The first thing that comes to your mind is usually the correct answer.

Your answer must appear clearly. Use black or dark blue pen for filling. If you answer incorrectly, completely fill the square and check the correct box. The form must not be folded.

When you`re done filling out the questionnaires, deliver the form to one of the research workers.

Today`s date *(ddmmyyyy)*: ☐☐·☐☐·☐☐☐☐

#### BACKGROUND INFORMATION

**1) In general, how would you rate your health today?**

☐ Very good

☐ Good

☐ Moderate

☐ Bad

☐ Very bad

**2) "I perceive stress in life, both personally and at work…"**

☐ Very often

☐ Often

☐ Sometimes

☐ Rarely or Never

**3) How important is it for you to exercise and maintain a healthy diet?**
Scale from 1 – 10 (1 = not important at all, and 10 = very important)

| 1 | 2 | 3 | 4 | 5 | 6 | 7 | 8 | 9 | 10 |
|---|---|---|---|---|---|---|---|---|---|

**4) If you consider exercising and maintaining a healthy diet important, do you believe it`s feasible for you to maintain that lifestyle?**
Scale from 1 – 10 (1 = no confidence, and 10 = highly confident)

| 1 | 2 | 3 | 4 | 5 | 6 | 7 | 8 | 9 | 10 |
|---|---|---|---|---|---|---|---|---|---|

**5) Do you smoke or use "snus"?**

☐ Yes, ca. ☐☐ cigarettes daily

☐ Yes, ca. ☐☐ «snus» daily

☐ No

**6) Have you started or stopped on any medications since last time?**

☐ Yes, I have stopped. Specify type.......................................................................................................

☐ Yes, I have started. Specify type.........................................................................................................

☐ No, I haven't started / stopped on any medication since last time

**7) Have you had any bleeding in this pregnancy?**

☐ Yes

☐ No (proceed to question 9)

**8) If you have had bleedings during this pregnancy, please state the date when the bleeding started, how many days it lasted and how much you bled:**

| **Date** *(ddmmyyyy)* | **Duration of bleeding** (number of days) | **Amount of blood** |
|---|---|---|
| **Date 1. bleeding** ☐☐·☐☐·☐☐☐☐ | ☐☐ | ☐ Spotting<br>☐ More than spotting |
| **Date 2. bleeding** ☐☐·☐☐·☐☐☐☐ | ☐☐ | ☐ Spotting<br>☐ More than spotting |
| **Date 3. bleeding** ☐☐·☐☐·☐☐☐☐ | ☐☐ | ☐ Spotting<br>☐ More than spotting |

**9) Have you had any nausea during this pregnancy?**

☐ Yes

☐ No (proceed to question 11)

**10) If you have had pregnancy sickness during this pregnancy, please state the date when the nausea started and how long it lasted (if you are still sick, leave it blank):**

Nausea started (dd.mm.yyyy) ☐☐.☐☐.☐☐☐☐

Nausea lasted until (dd.mm.yyyy) ☐☐.☐☐.☐☐☐☐

**11) Have you been off sick during this pregnancy?**

☐ Yes

☐ No (proceed to question 13)

**12) If you have been off sick during this pregnancy, please specify below:**

a) Reported sick, from (ddmmyyyy): ☐☐.☐☐.☐☐☐☐

until (ddmmyyyy): ☐☐.☐☐.☐☐☐☐

b) Sickness absence percentage: ☐☐☐ %

c) Are you actively on sick leave? ☐ Yes ☐ No

d) Reason for sick leave:________________________________________

________________________________________________________

________________________________________________________

e) Comments:________________________________________________

________________________________________________________

________________________________________________________

**13) Have you maintained the same level of physical activity as before your pregnancy?**

☐ I was more active before pregnancy

☐ I am as active as before the pregnancy

☐ I am more active now than before pregnancy

#### WELLNESS QUESTIONS

**14) How have you been feeling in general? (DURING THE PAST WEEK)**

☐ In excellent spirits

☐ In very good spirits

☐ In good spirits mostly

☐ I have been up and down in spirits a lot

☐ In low spirits mostly

☐ In very low spirits

**15) How often were you bothered by any illness, bodily disorder, aches or pains? (DURING THE PAST WEEK)**

☐ Every day

☐ Almost every day

☐ About half of the time

☐ Now and then, but less than half the time

☐ In low spirits mostly

☐ In very low spirits

**16) Did you feel depressed? (DURING THE PAST WEEK)**

☐ Yes – to the point that I felt like taking my life

☐ Yes – to the point that I did not care about anything

☐ Yes – very depressed almost every day

☐ Yes – quite depressed almost every day

☐ Yes – a little depressed now and then

☐ No – never felt depressed at all

**17) Have you been in firm control of your behavior, thoughts, emotions, or feelings? (DURING THE PAST WEEK)**

☐ Yes, definitely so

☐ Yes, for the most part

☐ Generally so

☐ Not too well

☐ No, and I am somewhat disturbed

☐ No, and I am very disturbed

**18) Have you been bothered by nervousness or your "nerves"? (DURING THE PAST WEEK)**

☐ Extremely so - to the point where I could not work or take care of things

☐ Very much so

☐ Quite a bit

☐ Some-enough to bother me

☐ A little

☐ Not at all

**19) How much energy, pop, or vitality did you have or feel? (DURING THE PAST WEEK)**

☐ Very full of energy – lots of pep

☐ Fairly energetic most of the time

☐ My energy level varied quite a bit

☐ Generally low in energy or pep

☐ Very low in energy or pep most of the time

☐ No energy or pep at all – I felt drained, sapped

**20) I felt downhearted and blue DURING THE PAST WEEK.**

☐ None of the time

☐ A little of the time

☐ Some of the time

☐ A good bit of the time

☐ Most of the time

☐ All of the time

**21) Were you generally tense-or did you feel any tension? (DURING THE PAST WEEK)**

☐ Yes – extremely tense, most or all of the time

☐ Yes – very tense most of the time

☐ Not generally tense, but did feel fairly tense several times

☐ I felt a little tense a few times

☐ My general tension level was quite low

☐ I never felt tense or any tension at all

**22) How happy, satisfied, or pleased have you been with your personal life? (DURING THE PAST WEEK)**

☐ Extremely happy – could not have been more satisfied or pleased

☐ Very happy most of the time

☐ Generally satisfied - pleased

☐ Sometimes fairly happy, sometimes fairly unhappy

☐ Generally dissatisfied, unhappy

☐ Very dissatisfied or unhappy most or all the time

**23) Did you feel healthy enough to carry out the things you like to do or had to do? (DURING THE PAST WEEK)**

☐ Yes – definitely so

☐ For the most part

☐ Health problems limited me in some important ways

☐ I was only healthy enough to take care of myself

☐ I needed some help in taking care of myself

☐ I needed someone to help me with most or all of the things I had to do

**24) Have you felt so sad, discouraged, hopeless, or had so many problems that you wondered if anything was worthwhile? (DURING THE PAST WEEK)**

☐ Extremely so-to the point that I have just about given up

☐ Very much so

☐ Quite a bit

☐ Some - enough to bother me

☐ A little bit

☐ Not at all

**25) I woke up feeling fresh and rested DURING THE PAST WEEK**

☐ None of the time

☐ A little of the time

☐ Some of the time

☐ A good bit of the time

☐ Most of the time

☐ All of the time

**26) Have you been concerned, worried, or had any fears about your health? (DURING THE PAST WEEK)**

☐ Extremely so

☐ Very much so

☐ Quite a bit

☐ Some, but not a lot

☐ Practically never

☐ Not at all

**27) Have you had any reason to wonder if you were losing your mind, or losing control over the way act, talk, think, feel or of your memory? (DURING THE PAST WEEK)**

☐ Not at all

☐ Only a little

☐ Some – but not enough to be concerned or worried about

☐ Some and I have been a little concerned

☐ Some and I am quite concerned

☐ Yes, very much so and I am very concerned

**28) My daily life was full of things that were interesting to me DURING THE PAST WEEK**

- ☐ None of the time
- ☐ A little of the time
- ☐ Some of the time
- ☐ A good bit of the time
- ☐ Most of the time
- ☐ All of the time

**29) Did you feel active, vigorous, or dull, sluggish? (DURING THE PAST WEEK)**

- ☐ Very active, vigorous every day
- ☐ Mostly active, vigorous – never really dull, sluggish
- ☐ Fairly active, vigorous – seldom dull, sluggish
- ☐ Fairly dull, sluggish - seldom active, vigorous
- ☐ Mostly dull, sluggish – never really active, vigorous
- ☐ Very dull, sluggish every day

**30) Have you been anxious, worried, or upset? (DURING THE PAST WEEK)**

- ☐ Extremely so – to the point of being sick or almost sick
- ☐ Very much so
- ☐ Quite a bit
- ☐ Some – enough to bother me
- ☐ A little bit
- ☐ Not at all

**31) I was emotionally stable and sure of myself DURING THE PAST WEEK**

☐ None of the time

☐ A little of the time

☐ Some of the time

☐ A good bit of the time

☐ Most of the time

☐ All of the time

**32) Did you feel relaxed, at ease or high strung, tight, or keyed-up? (DURING THE PAST WEEK)**

☐ Felt relaxed and at ease the whole month

☐ Felt relaxed and at ease most of the time

☐ Generally felt relaxed but at time felt fairly high strung

☐ Generally felt high strung but at time felt fairly relaxed

☐ Felt high strung, tight, or keyed up most of the time

☐ Felt high strung, tight, or keyed up the whole month

**33) I felt cheerful, lighthearted DURING THE PAST WEEK**

☐ None of the time

☐ A little of the time

☐ Some of the time

☐ A good bit of the time

☐ Most of the time

☐ All of the time

**34) I felt tired, worn out, used up, or exhausted DURING THE PAST WEEK**

☐ None of the time

☐ A little of the time

☐ Some of the time

☐ A good bit of the time

☐ Most of the time

☐ All of the time

**35) Have you been under or felt you were under any strain, stress, or pressure? (DURING THE PAST WEEK)**

☐ Yes, almost more than I could bear or stand

☐ Yes, quite a bit of pressure

☐ Yes, some – more than usual

☐ Yes, some – but about usual

☐ Yes, a little

☐ Not at all

### APPENDIX F - THE HORNE-ÖSTBERG MORNINGNESS-EVENINGNESS QUESTIONNAIRE

**MORNINGNESS-EVENINGNESS QUESTIONNAIRE**
**Self-Assessment Version (MEQ-SA)[1]**

Name: ______________________________ Date: ________________________

For each question, please select the answer that best describes you by circling the point value that best indicates how you have felt in recent weeks.

1. *Approximately* what time would you get up if you were entirely free to plan your day?

   [5] 5:00 AM–6:30 AM *(05:00–06:30 h)*
   [4] 6:30 AM–7:45 AM *(06:30–07:45 h)*
   [3] 7:45 AM–9:45 AM *(07:45–09:45 h)*
   [2] 9:45 AM–11:00 AM *(09:45–11:00 h)*
   [1] 11:00 AM–12 noon *(11:00–12:00 h)*

2. *Approximately* what time would you go to bed if you were entirely free to plan your evening?

   [5] 8:00 PM–9:00 PM *(20:00–21:00 h)*
   [4] 9:00 PM–10:15 PM *(21:00–22:15 h)*
   [3] 10:15 PM–12:30 AM *(22:15–00:30 h)*
   [2] 12:30 AM–1:45 AM *(00:30–01:45 h)*
   [1] 1:45 AM–3:00 AM *(01:45–03:00 h)*

3. If you usually have to get up at a specific time in the morning, how much do you depend on an alarm clock?

   [4] Not at all
   [3] Slightly
   [2] Somewhat
   [1] Very much

---

[1]Some stem questions and item choices have been rephrased from the original instrument (Horne and Östberg, 1976) to conform with spoken American English. Discrete item choices have been substituted for continuous graphic scales. Prepared by Terman M, Rifkin JB, Jacobs J, White TM (2001), New York State Psychiatric Institute, 1051 Riverside Drive, Unit 50, New York, NY, 10032. January 2008 version. Supported by NIH Grant MH42931. *See also:* automated version (AutoMEQ) at www.cet.org.

Horne JA and Östberg O. A self-assessment questionnaire to determine morningness-eveningness in human circadian rhythms. International Journal of Chronobiology, 1976: 4, 97-100.

4. How easy do you find it to get up in the morning (when you are not awakened unexpectedly)?

    [1] Very difficult
    [2] Somewhat difficult
    [3] Fairly easy
    [4] Very easy

5. How alert do you feel during the first half hour after you wake up in the morning?

    [1] Not at all alert
    [2] Slightly alert
    [3] Fairly alert
    [4] Very alert

6. How hungry do you feel during the first half hour after you wake up?

    [1] Not at all hungry
    [2] Slightly hungry
    [3] Fairly hungry
    [4] Very hungry

7. During the first half hour after you wake up in the morning, how do you feel?

    [1] Very tired
    [2] Fairly tired
    [3] Fairly refreshed
    [4] Very refreshed

8. If you had no commitments the next day, what time would you go to bed compared t your usual bedtime?

    [4] Seldom or never later
    [3] Less that 1 hour later
    [2] 1-2 hours later
    [1] More than 2 hours later

9. You have decided to do physical exercise. A friend suggests that you do this for one hour twice a week, and the best time for him is between 7-8 AM *(07-08 h)*. Bearing in mind nothing but your own internal "clock," how do you think you would perform?

   [4] Would be in good form
   [3] Would be in reasonable form
   [2] Would find it difficult
   [1] Would find it very difficult

10. At *approximately* what time in the evening do you feel tired, and, as a result, in need of sleep?

   [5] 8:00 PM–9:00 PM *(20:00–21:00 h)*
   [4] 9:00 PM–10:15 PM *(21:00–22:15 h)*
   [3] 10:15 PM–12:45 AM *(22:15–00:45 h)*
   [2] 12:45 AM–2:00 AM *(00:45–02:00 h)*
   [1] 2:00 AM–3:00 AM *(02:00–03:00 h)*

11. You want to be at your peak performance for a test that you know is going to be mentally exhausting and will last two hours. You are entirely free to plan your day. Considering only your "internal clock," which one of the four testing times would you choose?

   [6] 8 AM–10 AM *(08–10 h)*
   [4] 11 AM–1 PM *(11–13 h)*
   [2] 3 PM–5 PM *(15–17 h)*
   [0] 7 PM–9 PM *(19–21 h)*

12. If you got into bed at 11 PM *(23 h)*, how tired would you be?

   [0] Not at all tired
   [2] A little tired
   [3] Fairly tired
   [5] Very tired

13. For some reason you have gone to bed several hours later than usual, but there is no need to get up at any particular time the next morning. Which one of the following are you most likely to do?

[4] Will wake up at usual time, but will not fall back asleep
[3] Will wake up at usual time and will doze thereafter
[2] Will wake up at usual time, but will fall asleep again
[1] Will not wake up until later than usual

14. One night you have to remain awake between 4-6 AM *(04-06 h)* in order to carry out a night watch. You have no time commitments the next day. Which one of the alternatives would suit you best?

[1] Would not go to bed until the watch is over
[2] Would take a nap before and sleep after
[3] Would take a good sleep before and nap after
[4] Would sleep only before the watch

15. You have two hours of hard physical work. You are entirely free to plan your day. Considering only your internal "clock," which of the following times would you choose?

[4] 8 AM–10 AM *(08–10 h)*
[3] 11 AM–1 PM *(11–13 h)*
[2] 3 PM–5 PM *(15–17 h)*
[1] 7 PM–9 PM *(19–21 h)*

16. You have decided to do physical exercise. A friend suggests that you do this for one hour twice a week. The best time for her is between 10-11 PM *(22-23 h)*. Bearing in mind only your internal "clock," how well do you think you would perform?

[1] Would be in good form
[2] Would be in reasonable form
[3] Would find it difficult
[4] Would find it very difficult

17. Suppose you can choose your own work hours. Assume that you work a five-hour day (including breaks), your job is interesting, and you are paid based on your performance. At *approximately* what time would you choose to begin?

    [5] 5 hours starting between 4–8 AM *(05–08 h)*
    [4] 5 hours starting between 8–9 AM *(08–09 h)*
    [3] 5 hours starting between 9 AM–2 PM *(09–14 h)*
    [2] 5 hours starting between 2–5 PM *(14–17 h)*
    [1] 5 hours starting between 5 PM–4 AM *(17–04 h)*

18. At *approximately* what time of day do you usually feel your best?

    [5] 5–8 AM *(05–08 h)*
    [4] 8–10 AM *(08–10 h)*
    [3] 10 AM–5 PM *(10–17 h)*
    [2] 5–10 PM *(17–22 h)*
    [1] 10 PM–5 AM *(22–05 h)*

19. One hears about "morning types" and "evening types." Which one of these types do you consider yourself to be?

    [6] Definitely a morning type
    [4] Rather more a morning type than an evening type
    [2] Rather more an evening type than a morning type
    [1] Definitely an evening type

____ **Total points for all 19 questions**

### APPENDIX G – PITTSBURGH SLEEP QUALITY INDEX (PSQI)

#### The Pittsburgh Sleep Quality Index (PSQI)

**Name:** ______________________ **Date:** ______________

**Instructions:** The following questions relate to your usual sleep habits during the past month only. Your answers should indicate the most accurate reply for the majority of days and nights in the past month. Please answer all questions.
During the past month,

1. When have you usually gone to bed? ______________

2. How long (in minutes) has it taken you to fall asleep each night? ______________

3. When have you usually gotten up in the morning? ______________

4. How many hours of actual sleep do you get at night? (This may be different than the number of hours you spend in bed) ______________

Please check the appropriate blank below.

| 5. During the past month, how often have you had trouble sleeping because you... | Not during the past month (0) | Less than once a week (1) | Once or twice a week (2) | Three or More times a week (3) |
|---|---|---|---|---|
| a. Cannot get to sleep within 30 minutes | a. _____ | _____ | _____ | _____ |
| b. Wake up in the middle of the night or early morning | b. _____ | _____ | _____ | _____ |
| c. Have to get up to use the bathroom | c. _____ | _____ | _____ | _____ |
| d. Cannot breathe comfortable | d. _____ | _____ | _____ | _____ |
| e. Cough or snore loudly | e. _____ | _____ | _____ | _____ |
| f. Feel too cold | f. _____ | _____ | _____ | _____ |
| g. Feel too hot | g. _____ | _____ | _____ | _____ |
| h. Have bad dreams | h. _____ | _____ | _____ | _____ |
| i. Have pain | i. _____ | _____ | _____ | _____ |
| j. Other reason(s), please describe, including how often you have had trouble sleeping because of this reason(s): | j. _____ | _____ | _____ | _____ |
| 6. During the past month, how often have you taken medicine (prescribed or "over the counter") to help you sleep? | 6. _____ | _____ | _____ | _____ |
| 7. During the past month, how often have you had trouble staying awake while driving, eating meals, or engaging in social activity? | 7. _____ | _____ | _____ | _____ |
| 8. During the past month, how much of a problem has it been for you to keep up enthusiasm to get things done? | 8. _____ | _____ | _____ | _____ |

| | Very good (0) | Fairly good (1) | Fairly bad (2) | Very bad (3) |
|---|---|---|---|---|
| 9. During the past month, how would you rate your sleep quality overall? | 9. _____ | _____ | _____ | _____ |

Physician Determined Global PSQI Score: ________

### APPENDIX H – BACKGROUND INFORMATION (FATHER), TEST 1

BEFORE THE BEGINNING – TEST 1 Initials ☐☐☐☐ ID-number ☐☐☐

Attached is a questionnaire that we would like you to fill out. Your answer is an important contribution to understanding the importance of your overall health and several background variables in relation to other measurements in the study.

Please answer all the questions. Do not spend too long on each question. The first thing that comes to your mind is usually the correct answer.

Your answer must appear clearly. Use black or dark blue pen for filling. If you answer incorrectly, completely fill the square and check the correct box. The form must not be folded.

When you`re done filling out the questionnaires, deliver the form to one of the research workers.

Today`s date *(ddmmyyyy)*: ☐☐·☐☐·☐☐☐☐

#### BACKGROUND INFORMATION

**1) Date of birth:** ☐☐·☐☐·☐☐☐☐

**2) Weight:** ☐☐☐kg

**3) Height:** ☐☐☐cm

**4) Ethnicity**

☐ Norwegian

☐ Other: ……………………………………………….

**5) Highest educational level**

☐ Compulsory schooling

☐ Upper secondary school

☐ University, less than 4 years

☐ University, 4 years or more

**6) Paid employment?**

(Also include temporary – and short-term employment. Don't take into account whether you are off sick or on leave. Paid employment also include work without a fixed salary in a company owned by the family, e.g. farm / business)

☐ Yes, I`m ☐☐☐ % employed

☐ No, I`m a ☐ Student

☐ Disabled

☐ Homemaker

☐ Unemployed

**7) What kind work do you do?**

☐ Health/social work ☐ Teaching/research

☐ Office ☐ Industry

☐ Construction and building ☐ Shop assistant

☐ Service trade ☐ Other: ………………………………………………

**8) Working hours?**

☐ Daytime

☐ Afternoon/evening

☐ Shift work or rotation scheme

☐ No fixed rotation scheme (extra help, stand-in etc.)

**9) Do you work on foot or standing?**

☐ Yes, daily, more than half of the hours worked

☐ Yes, daily, less than half of the hours worked

☐ Periodic, but not daily

☐ Rarely or never

**10) How active are you at your work?**

☐ Sedentary (Sit all day)

☐ Partly sedentary (Walk or stand part of the day)

☐ Active (Walk or stand all day)

☐ Very active

**11) In general, how would you rate your health today?**

☐ Very good

☐ Good

☐ Moderate

☐ Bad

☐ Very bad

**12) "I perceive stress in life, both personally and at work…"**

☐ Very often

☐ Often

☐ Sometimes

☐ Rarely or Never

**13) How would you say your diet is? Based on the LAST 14 DAYS.**

☐ Very healthy

☐ Healthy

☐ Neither healthy nor unhealthy

☐ Unhealthy

☐ Very unhealthy

**14) Do you meet the recommended amount of physical activity? Adult aged 18-64 should do at least of 150 minutes of moderate-intensity aerobic physical activity throughout the week or do at least 75 minutes of vigorous-intensity aerobic physical activity throughout the week or an equivalent combination of moderate- and vigorous-intensity activity.**

☐ Yes

☐ No

**15) Do you smoke or use "snus"? Based on the LAST 14 DAYS.**

☐ Yes, ca. ☐☐ cigarettes daily

☐ Yes, ca. ☐☐ «snus» daily

☐ No

**16) Have you smoked in the past?**

☐ Yes

☐ No (Proceed to question 19)

**17) If so, how many years have you smoked in total?**

☐☐ Years

**18) If so, when did you quit smoking?**

☐☐ years ago

**19) How often have you been drinking alcohol (beer, wine or liquor) in the LAST 14 DAYS?**

☐ I have not been drinking, but is not teetotal (Proceed to question 19)

☐ 1 - 4 time

☐ 5 – 10 times

☐ More than 10 times

☐ Teetotal, never drink alcohol (Proceed to question 19)

**20) If you have been drinking alcohol for the LAST 14 DAYS, have you ever felt drunk?**

☐ Yes

☐ No

**21) Have there been periods in your life when you have been drinking too much?**

☐ No

☐ Maybe

☐ Yes

**22) Do you use any medications?**

☐ Yes, Specify type……………………………………………………………………………..

☐ No

#### CASE HISTORY

**23) Do you have any diseases?**

☐ Yes:____________________________________________________________

________________________________________________________________

________________________________________________________________

☐ No

**24) Are there any close relatives who have:**

☐ Diabetes type 1 ☐ High blood pressure

☐ Diabetes type 2 ☐ Overweight/obesity

☐ None of the above

### APPENDIX I – BACKGROUND INFORMATION (FATHER), TEST 2

BEFORE THE BEGINNING – TEST 2 Initials ☐☐☐☐ ID-number ☐☐☐

Attached is a questionnaire that we would like you to fill out. Your answer is an important contribution to understanding the importance of your overall health and several background variables in relation to other measurements in the study.

Please answer all the questions. Do not spend too long on each question. The first thing that comes to your mind is usually the correct answer.

Your answer must appear clearly. Use black or dark blue pen for filling. If you answer incorrectly, completely fill the square and check the correct box. The form must not be folded.

When you`re done filling out the questionnaires, deliver the form to one of the research workers.

Today`s date *(ddmmyyyy)*: ☐☐.☐☐.☐☐☐☐

#### BACKGROUND INFORMATION

**1) Weight:** ☐☐☐kg

**2) In general, how would you rate your health today?**

☐ Very good

☐ Good

☐ Moderate

☐ Bad

☐ Very bad

**3) "I perceive stress in life, both personally and at work…"**

☐ Very often

☐ Often

☐ Sometimes

☐ Rarely or Never

**4) How would you say your diet is? Based on the LAST 14 DAYS.**

☐ Very healthy

☐ Healthy

☐ Neither healthy nor unhealthy

☐ Unhealthy

☐ Very unhealthy

**5) Do you meet the recommended amount of physical activity? Adult aged 18-64 should do at least of 150 minutes of moderate-intensity aerobic physical activity throughout the week or do at least 75 minutes of vigorous-intensity aerobic physical activity throughout the week or an equivalent combination of moderate- and vigorous-intensity activity.**

☐ Yes

☐ No

**6) Do you smoke or use "snus"? Based on the LAST 14 DAYS.**

☐ Yes, ca. ☐☐ cigarettes daily

☐ Yes, ca. ☐☐ «snus» daily

☐ No

**7) How often have you been drinking alcohol (beer, wine or liquor) in the LAST 14 DAYS?**

☐ I have not been drinking, but is not teetotal (Proceed to question 9)

☐ 1 - 4 time

☐ 5 – 10 times

☐ More than 10 times

☐ Teetotal, never drink alcohol (Proceed to question 9)

**8) If you have been drinking alcohol for the LAST 14 DAYS, have you ever felt drunk?**

☐ Yes

☐ No

**9) Have you started or stopped on any medications since last time?**

☐ Yes, I have stopped. Specify type………………………………………………………………………………..

☐ Yes, I have started. Specify type………………………………………………………………………………..

☐ No, I haven't started / stopped on any medication since last time
